# δ subvariants of SARS-COV-2 in Israel, Qatar and Bahrain: Optimal vaccination as an effective strategy to block viral evolution and control the pandemic

**DOI:** 10.1101/2021.11.01.21265445

**Authors:** Xiang-Jiao Yang

## Abstract

δ variant has rapidly become the predominant pandemic driver and yielded four subvariants (δ1, δ2, δ3 and δ4). Among them, δ1 has been mainly responsible for the latest COVID-19 waves in India, Southeast Asia, Europe and the USA. A relevant question is how δ subvariants may have driven the pandemic in the rest of the world. In both Israel and Qatar, mRNA-based vaccination has been rolled out competitively, but the outcomes are quite different in terms of controlling the recent waves resulting from δ variant. This raises the question whether δ subvariants have acted differently in Israel and Qatar. In both countries, δ variant was first identified in April 2021 and δ1 subvariant constituted ∼50% δ genomes from April to May 2021. But the situation started to diverge in June 2021: In Israel, δ1 variant was encoded by 92.0% δ genomes, whereas this fraction was only 43.9% in Qatar. Moreover, a δ1 sublineage encoding spike T791I was identified in Israel but not Qatar. This sublineage accounted for 31.8% δ genomes sequenced in June 2021 and declined to 13.3% in October 2021. In August 2021, δ1 also became dominant in Qatar and a major sublineage encoding spike D1259H emerged. This sublineage has evolved further and acquired additional spike substitutions, including K97E, S255F, I693S, I712S, I1104L, E1258D and/or V1177I, in Qatar and other countries, such as Czech Republic, France and Mexico. Monthly distribution of the above sublineages suggests that the one from Qatar is much more of concern than that from Israel. Different from what was in Israel and Qatar, δ2 subvariant has also been important in Bahrain, whereas a δ2 sublineage encoding spike V1264L and A1736V of NSP3 was dominant in June 2021, but was gradually taken over by δ1 subvariant. These results suggest that δ1 and δ2 subvariants continue their evolution in different countries. The recent successful pandemic control in Israel, Qatar and Bahrain supports that δ1 and δ2 subvariants are still sensitive to timed vaccination, thereby urging the use of optimal immunity as a strategy to block SARS-COV-2 evolution and control the pandemic.

## INTRODUCTION

Coronavirus disease 2019 (COVID-19) has caused the pandemic, led to tragic loss of life, overloaded health care systems and crippled the economy around the world. Genomic surveillance of SARS-CoV-2 has firmly established that its evolution is the driving force of this pandemic [1]. In addition to many variants of interests [1], the virus has yielded four variants of concern as designated by the WHO: α (B.1.1.7) [2], β (B.1.351) [3], γ (P.1) [4] and δ (B.1.617.2) [5]. I reasoned that the evolutionary trajectory of SARS-COV-2 would yield important insights into potential candidates for future variants of concern, which would in turn help us take pre-emptive strikes against such candidates. To dissect the evolutionary trajectory, I have leveraged the >4 million SARS-COV-2 genomes in the GISAID (global initiative on sharing avian influenza data) sequence database [6]. As a scientist who had focused mainly on mouse development, genetic disease and cancer before the pandemic [7-10], I have been so impressed and deeply moved by the coordinated global efforts in generating and maintaining this invaluable GISAID resource when such an unprecedented challenge arrived in a totally unexpected manner. When I found this database during the initial lockdown in March 2020, I asked whether my expertise and knowledge about genetic and cancer mutations, protein structure and function, and post-translational protein modifications [11] could be helpful for annotating SARS-COV-2 mutations. Because of the GISAID database [6] and two highly useful bioinformatic tools developed by scientists in Europe [12-14], I have recently found that δ variant had evolved and yielded four subvariants (δ1, δ2, δ3 and δ4) in India before spreading to other countries [15]. Among the subvariants, δ1 has emerged as the major pandemic driver and δ2 plays a much less important role, whereas δ3 and δ4 have gradually faded away [15]. δ1 subvariant is also a major pandemic driver in Europe [15], the USA [16], Indonesia, Singapore and Malaysia [17], raising the question whether it is also true in the rest of the world.

In parallel, I have followed how effective vaccination has been against SARS-COV-2 and its variants in different countries around the world. While I was so thrilled to witness that after Israel rolled out its vaccination program, COVID-19 cases dropped sharply in the spring of 2021, it was puzzling that δ variant then caused a surging wave in the summer. In comparison, the latest wave due to δ variant is much less severe in Qatar, another pioneer in rolling out mRNA-based vaccines. This difference raises the possibility that composition of δ subvariants may have contributed to the different outcomes in Israel and Qatar. In both countries, δ1 subvariant constituted ∼50% δ genomes from April to May 2021. But the situation started to diverge in June 2021, with δ1 variant being much more dominant in Israel than it was in Qatar. Moreover, a δ1 sublineage encoding spike T791I was identified in Israel but not Qatar. Thus, composition of δ subvariants in June 2021 may be one factor behind the different pandemic outcomes in Israel and Qatar. Another difference is the pace at which vaccination was rolled out: Israel had achieved full vaccination in >50% of its population by the end of March 2021, but Qatar only reached a similar level in June 2021 (Our World in Data, https://ourworldindata.org/; accessed on October 29, 2021). Related to this, two recent studies have uncovered that waning immunity has contributed significantly to the latest surging wave of COVID-19 cases in Israel [18,19].

In August 2021, δ1 also became dominant in Qatar and a major sublineage encoding D1259H at the cytoplasmic tail of spike protein emerged. Unexpectedly, monthly distribution of δ genomes suggests that this sublineage is more of concern than that from Israel. For comparison, I have analyzed SARS-COV-2 genomes identified in Bahrain, another Middle East country that has also rolled out a vaccination program competitively. Different from what was observed in Israel and Qatar, both δ1 and δ2 subvariants have been important in Bahrain. In the country, a δ2 sublineage encoding spike V1264L, just five residues away from D1259H [17], was dominant in June 2021 but has been gradually taken over by δ1 subvariant since then. The recent successful pandemic control in all three countries suggest that these different sublineages are all sensitive to vaccination, reiterating that optimal immunity is an effective strategy to block evolution of δ subvariants and control the pandemic. In comparison, genomic information is relatively much more limited from other countries in the Middle East. Thus, to avoid fighting in the dark, it is important to use genomic surveillance as the guiding light to combat this constantly evolving virus. Both genomic surveillance and optimal vaccination should help us end the prolonged pandemic as swiftly as possible.

## RESULTS AND DISCUSSION

### SARS-COV-2 evolution causes multiple waves of COVID-19 cases in Israel

Since identification of the first COVD-19 case on February 21, 2020, there have been five waves in the country (Fig. 1A). Compared to the latest three waves, the first two are relatively minor. Among the five waves, the most recent one started at the beginning of June 2021 and is the most powerful (Fig. 1A). It should be noted, however, that despite being the most severe in terms of the confirmed daily case number, this wave ranked only the third in terms of mortality (Our World in Data, accessed on October 30, 2021). Still, it is puzzling that the case number is so high with the latest wave because vaccination started at January 2021 and 2-dose vaccination had reached 50.5% and 58.6% of the population by March 16 and June 04, 2021, respectively (Fig. 1A). Importantly, the country started to administrate booster doses at the beginning of August 2021 and has recently reached >44% of the population (Fig. 1A) [18,19]. As a result, the surging wave has been brought under control (Fig. 1A) [18,19]. This heroic effort was detailed in two important studies that have been published or posted online recently [18,19]. One explanation is why the surging wave occurred when 2-dose vaccination had reached such a high level is that apparent immunity from vaccination had waned by the summer [18,19]. For this, there may be two reasons that are not mutually exclusive: 1) vaccination-induced immunity *per se* declines as time passes, and 2) the virus evolves and yields variants that are more resistant to vaccination-induced immunity than the original strain.

**Figure 1.**
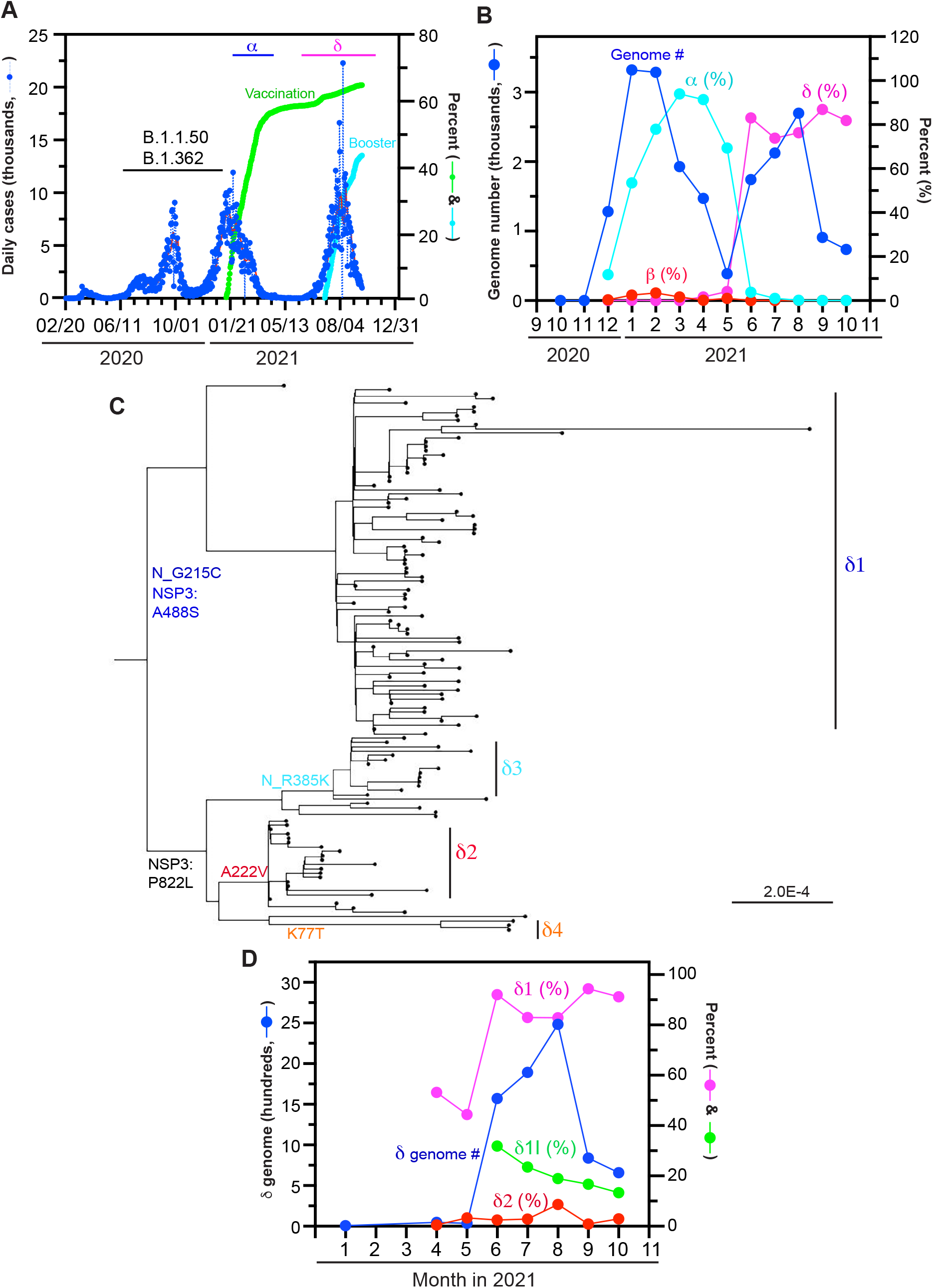
SARS-COV-2 evolution causes multiple waves of COVID-19 cases in Israel. (**A**) Epidemiological curve and vaccination progress in the country. The curve is composed of five waves, peaked on April 04, 2020; July 25, 2020; September 28, 2020; January 17, 2021 and September 14, 2021. While α variant was mainly responsible for the second half of the fourth wave, δ variant acted as the predominant driver of the fifth wave. For preparation of this panel, the Our World in Data website was accessed on October 18, 2021. (**B**) Monthly distribution of SARS-COV-2 variant genomes detected in Israel. **(C)** Phylogenetic analysis of 140 δ genomes identified in Israel by June 15, 2021. The analysis was carried out with the package RAxML-NG, to generate 20 maximum likelihood trees and one bestTree for presentation via FigTree. The strain names and GISAID accession numbers of the genomes are provided in Fig. S1. The genomes were downloaded from the GISAID SARS-COV-2 genome sequence database on August 22, 2021. (**D**) Monthly distribution of δ subvariant genomes identified in Israel. For preparation of panels B and D, the GASAID database was accessed on October 29, 2021.

To investigate the second possibility, I analyzed SARS-COV-2 genomes identified and sequenced in Israel. For this, I first quantified variant genomes deposited into the GISAID database by the country. This analysis revealed that by December 2020, multiple lineages, such as B.1.1, B.1.150 and B.1.362, were responsible for the first half of the fourth wave (Fig. 1A). In December 2020, α variant was detected (Fig. 1B). Its genome number rapidly rose and made it a major pandemic driver in January 2021 (Fig. 1B). In March and April 2021, it was almost the sole pandemic driver in the country (Fig. 1B). But its genome number declined in May and June 2021, at a speed even faster than its rapid rise in January and February (Fig. 1B). This decline coincided with the progress of vaccination (Fig. 1A). Mysteriously, this decline also coincided with the rise of the δ variant genome number (Fig. 1B). This variant soon became a dominant driver in June (Fig. 1B). However, the wave only started to surge in July (Fig. 1A), suggesting that there was a latent period in June 2021.

As described in several recent studies [15-17], δ variant has evolved and yielded different subvariants. To understand how such subvariants may have contributed to the latest surging wave In Israel, I carried out phylogenetic analysis of the 140 δ genomes that were identified there by June 15, 2021. The analysis was carried out through the package RAxML-NG [14] to generate 20 maximum likelihood trees and one bestTree for presentation via FigTree. As shown in Figs 1C & S1, the genomes form four distinct clusters corresponding to the four δ subvariants, very similar to what was described in three related studies about δ subvariants in India, Southeast Asia, Europe and the USA [15-17]. From the phylogenetic tree, it is also clear that δ1 subvariant had been dominant, with the other three being relatively minor, in Israel by June 15, 2021. In support of this, δ1 subvariant has been the dominant subvariant since then (Fig. 1D). Since June 2021, this subvariant has accounted for almost 90% of all genomes identified in the country (Fig. 1D). The δ2 subvariant fraction also rose slightly and reached ∼8.6% in August 2021, but it declined afterwards (Fig. 1D). Therefore, since June 2021, δ1 has been the dominant subvariant while δ2 subvariant has only played a minor role.

### Mutation profiling uncovers δ1I as a new δ1 sublineage

To examine the 140 δ genomes from Israel in more details, I carried out mutation profiling of these genomes, by utilizing Coronapp, an effective Web-based COVID-19 genome annotator [12,13]. Shown in Fig. 2A-B are substitutions in NSP3 and spike proteins. Three δ1-specific NSP3 substitutions, A488S, P1228L and P1469S [15-17], are present in ∼50% genomes, whereas the remaining fraction carries P822L (Fig. 2A), an NSP3 substitution specific to δ2, δ3 and δ4 [15-17]. In agreement with this, G215C and R385K of nucleocapsid protein are encoded by 90 (64%) and 17 (12%) genomes, respectively (data not shown). The δ2-specific spike substitution A222V is encoded by 22 (∼16%) genomes, whereas the δ4-specific spike substitution K77T is found with only three genomes (Fig. 2B). Such subvariant composition is good agreement with phylogenetic analysis (Fig. 1C), further supporting that δ1 had become dominant by June 15, 2021.

**Figure 2.**
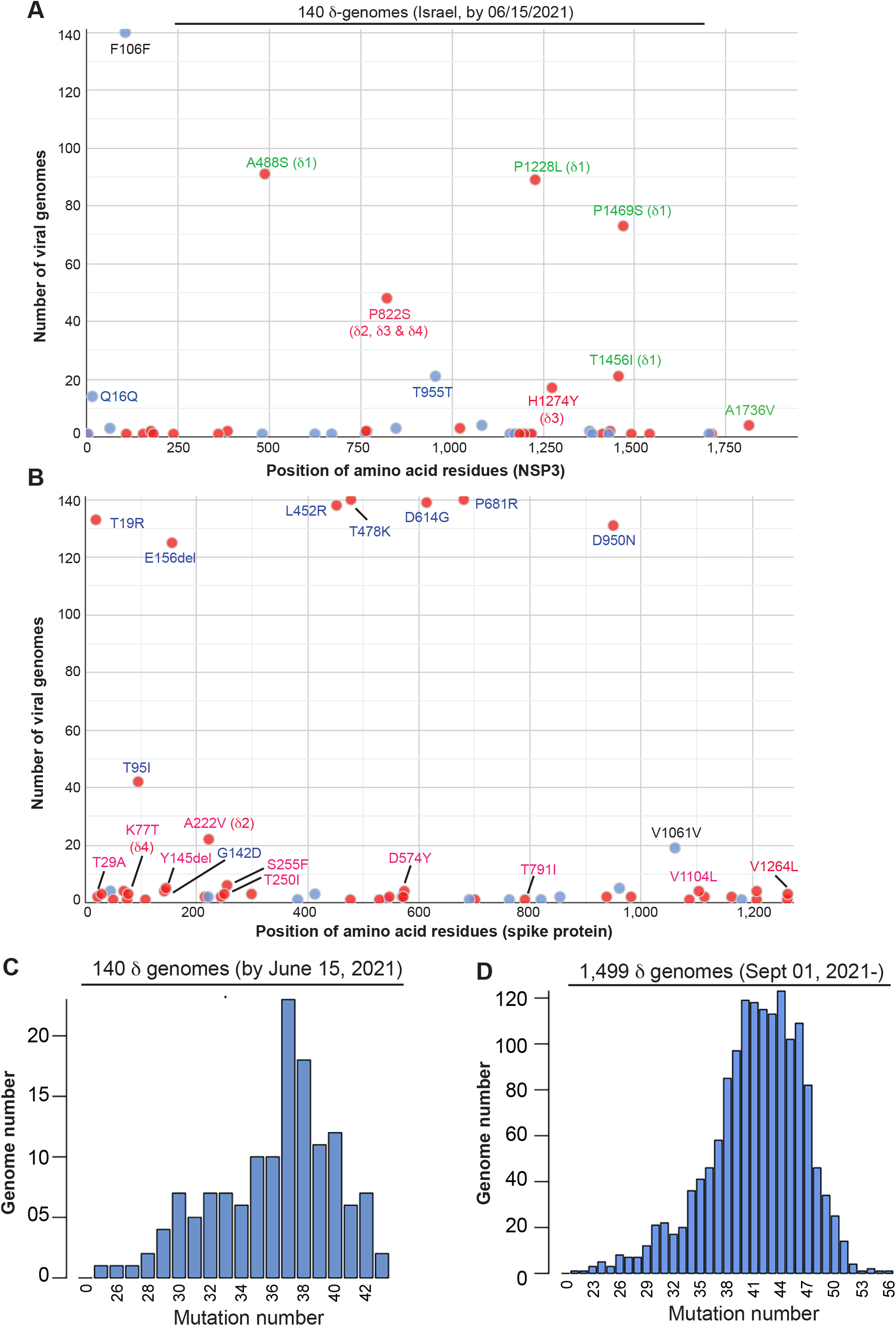
Mutation profiling of δ genomes identified in Israel. (**A-C**) Mutation profile of the 140 δ genomes identified in Israel by June 15, 2021. The genomes were downloaded from the GISAID database on August 22, 2021 for mutation profiling via Coronapp [12,13]. Shown in (A) and (B) are substitutions in NSP3 and spike proteins, respectively, whereas panel C displays the mutation loads of the genomes (i.e., numbers of mutations that the genomes carry). (**D**) Mutation load distribution of 1,499 δ genomes identified from September to October 2021. The genomes were downloaded from the GISAID database on October 29, 2021. See the Materials and Methods section about the markers used to identify the subvariants.

Based on the mutation load (i.e., mutation number per genome), there appears two populations, with the first peaked at 33 mutations per genome and the second population at 37 mutations per genome (Fig. 2C). Based on the previous studies [15-17], the first peak corresponds to δ2, δ3 and δ4 while the second peak is due to δ1. Notably, one δ1 genome encodes spike T791I and three δ2 genomes carry spike V1264L (Fig. 2B). As described below, these four genomes are related to two δ1 and δ2 sublineages identified in July and August 2021. Moreover, some genomes encode spike T29A, T250I and Q613H (Fig. 3B), substitutions corresponding to a new δ1 sublineage that originated in Morocco but has been identified in Europe [15]. In addition, one sublineage encodes I850L (Fig. 3B). Moreover, there are some δ1 genomes encoding S255F (Fig. 2B) and two genomes encoding spike A222V (data not shown). Thus, there are genomes corresponding to new δ1 sublineage.

**Figure 3.**
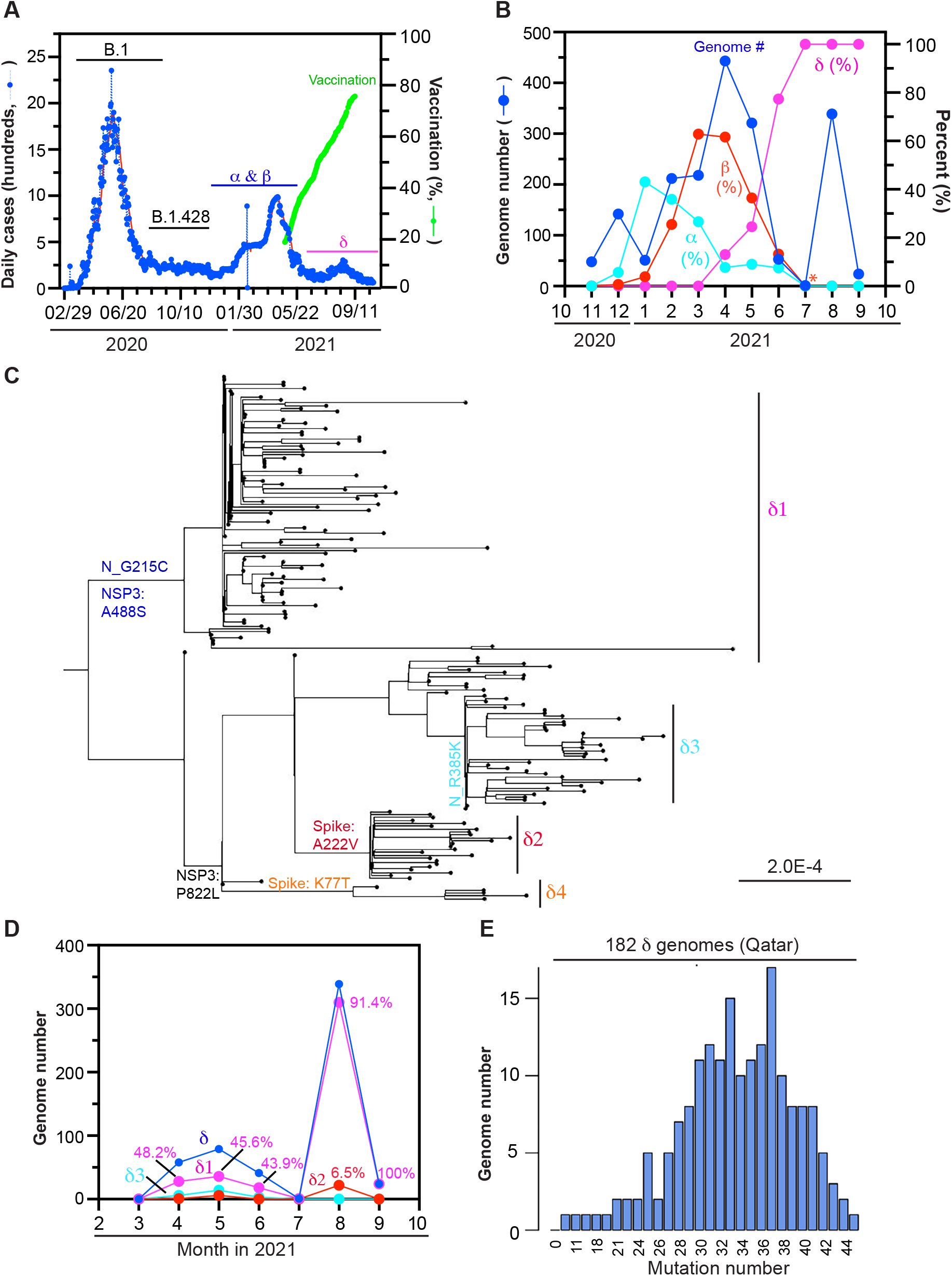
SARS-COV-2 variants as pandemic drivers in Qatar. (**A**) Epidemiological curve and vaccination progress in the country. There have been three waves, peaked on June 03, 2020; April 15, 2021 and August 20, 2021. While α and β variants contributed significantly to the second wave, δ variant is the sole driver of the third wave. For preparation of this panel, the Our World in Data website was accessed on October 18, 2021. (**B**) Monthly distribution of SARS-COV-2 variant genomes detected in Qatar. **(C)** Phylogenetic analysis of the 182 δ genomes identified in the country. The analysis was carried out through the package RAxML-NG as in Fig. 1C. The strain names and GISAID accession numbers of the genomes are provided in Fig. S2. 178 of them were identified by June 30, 2021 and 4 genomes were identified from July-August 2021. The genomes were downloaded from the GISAID database on October 17, 2021. (**D**) Monthly distribution of δ subvariant genomes detected in Qatar. For preparation of panels B and D, the GASAID database was accessed on October 26, 2021. (**E**) Mutation load distribution of the 182 δ genomes. The genomes were subject to mutation profiling via Coronapp [12,13].

To understand how δ variant would evolve in Israel after June 15, 2021, I carried out mutation profiling of 1,499 δ genomes identified there from September to October, by using Coronapp [12,13]. As shown in Fig. 2D, the average mutation load of the major peak is 42 mutations per genome, which is five mutations more than the average load of the second population identified by June 15, 2021 (Fig. 2C), suggesting that additional mutations have been gained since June 16, 2021. As shown in Fig. S3A, the predominant majority of the genomes encodes δ1 subvariant, with only a few for P822L. This P822L population of genomes is due to δ2 because there are a similar number of genomes encoding spike A222V (Fig. 3B). By contrast, no genomes encode spike K77T (Fig. S3B) or nucleocapsid R385K (Fig. S3C), so δ3 and δ4 had faded away in Israel by September 2021. Therefore, δ1 has become predominant while δ2 has only played a minor role.

One outstanding feature is that a significant subpopulation of the genomes encodes an extra spike substitution, T791I. The corresponding new lineage is referred to as δ1I, where the letter stands for I791. There were 500 δ1I genomes identified in June 2021, but this fraction (31.8% of all δ genomes uncovered during the month) decreased gradually to 13.3% in October 2021 (Fig. 1D). δ1I was first identified in Texas, USA but has spread to other countries around the world. There are over 3,310 such genomes in the GISAID database (accessed on October 30, 2021) and ∼20% of them (578 genomes, mainly from Europe) encode an extra nucleocapsid substitution, Q9L. This substitution alters the N-terminal tail of nucleocapsid and the tail is known immune epitope, so Q9L may confer immune evasion. No δ1I genomes from Israel encode this extra substitution.

Beside T791I, there are some genomes encoding I850L or V1264L (Fig. S3B). The latter is associated with δ2 variant. V1264L confers a dileucine motif in the cytoplasmic tail of spike protein, so a variant carrying such a substitution may exhibit added evolutionary advantage [17]. δ2L, a δ2 sublineage with V1264L, has been identified in the USA [17]. Such genomes had been identified in Israel by June 15, 2021 (Fig. 2B), so the sublineage might have been imported before then. This sublineage may explain why the δ2 genome number stably increased from June to August 2021 (Fig. 1D). There are a few genomes encoding three spike substitutions, T29A, T250I and Q613H (Fig. S3B). A related δ1 sublineage has been identified in Europe [15]. Moreover, there are a few δ1 genomes encoding spike A222V (data not shown). Some of these genomes also carry spike Y145H and are related to a new lineage known as AY.4.2 [1] or δ1V [15,17,20]. Therefore, since June 2021, δ1 subvariant and the sublineage δ1I have been the major pandemic drivers in Israel.

### δ subvariants in Qatar by June 2021

As shown in Fig. 3A, there have been three waves of COVID-19 cases in Qatar, peaked on June 03, 2020; April 15, 2021 and August 20, 2021. While both α and β variants contributed significantly to the second wave, δ variant was the sole driver of the third wave (Fig. 3A-B). To examine δ subvariants from Qatar, I carried out phylogenetic analysis of 182 δ genomes identified in the country, by using the package RAxML-NG [14]. 178 of the genomes had been identified by June 30, 2021, with the remaining 4 from July or August 2021. As shown in Fig. 3C, the genomes form four clusters, with the δ1 cluster roughly equal to the sum of the δ2, δ3 and δ4 clusters. This is different from the situation in Israel, where by June 15, 2021, the δ1 cluster had been much larger than the sum of the δ2, δ3 and δ4 clusters (Fig. 1C). Different from that in Israel (Fig. 1C), the δ3 cluster larger than the δ2 cluster (Figs 3C & S2). This is significant as δ2 appears to be more virulent than δ3 [15-17]. The mutation load distribution of the 182 δ genomes is shown in Fig. 3E. Clearly, the first population is much larger than the counterpart of the genomes from Israel (Fig. 2C). Mutation profiling of the 182 genomes (Fig. 4) confirms the findings made from phylogenetic analysis (Figs 3C & S2). Notably, neither spike T791I nor V1264L was identified in the genomes (Fig. 4B). Notable, the δ3 genomes encode an extra nucleocapsid substitution, L139F (Fig. 4C). Thus, there are several differences between Israel and Qatar in terms of δ subvariant genomes identified by June 2021.

**Figure 4.**
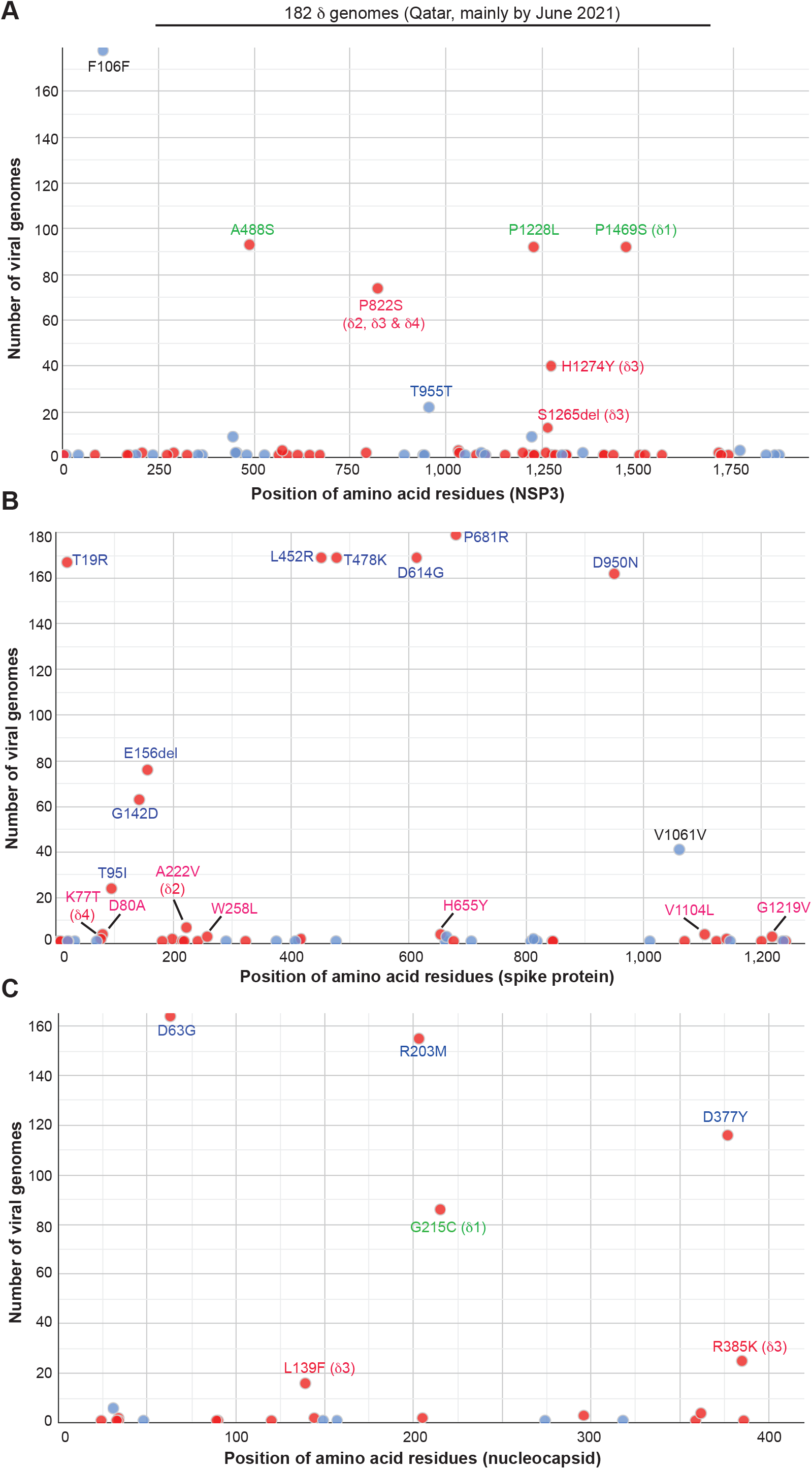
Mutation profiling of the 182 δ genomes identified in Qatar. The genomes were downloaded from the GISAID database for mutation profiling as in Fig. 4E. Shown here in (A), (B) and (C) are substitutions in NSP3, spike and nucleocapsid proteins, respectively.

### Emergence of spike D1259H-encoding δ1 sublineages in Qatar and other countries

To understand how δ variant would evolve in Qatar after June 2021, I carried out mutation profiling of δ genomes identified there afterwards, by using Coronapp [12,13]. There is one genome available for July 2021, so only 363 genomes identified from August and September 2021 were analyzed. Unexpectedly, about 50% of them encode an extra spike substitution, D1259H (Fig. 5A). This δ1 sublineage is referred to as δ1H, where the letter H stands for H1259. Intriguingly, this is located at the cytoplasmic tail of spike protein and only five residues away from V1264L, a potential driver mutation of a virulent δ1 sublineage behind the recent surging waves of COVID-19 cases in Indonesia, Singapore and Malaysia [17]. There are a few genomes encoding T29A, T255I and Q613H, three spike substitutions that are present in a δ1 sublineage already identified in Europe [15]. On such genome also encodes spike H69L and V70F, two substitutions in a loop that is often deleted in other variants. Although the genome number is still low (Fig. 5A), it may be important to watch out δ1H genomes encoding these extra spike substitutions (i.e., T29A, H69L, V70F, T255I and/or Q613H).

**Figure 5.**
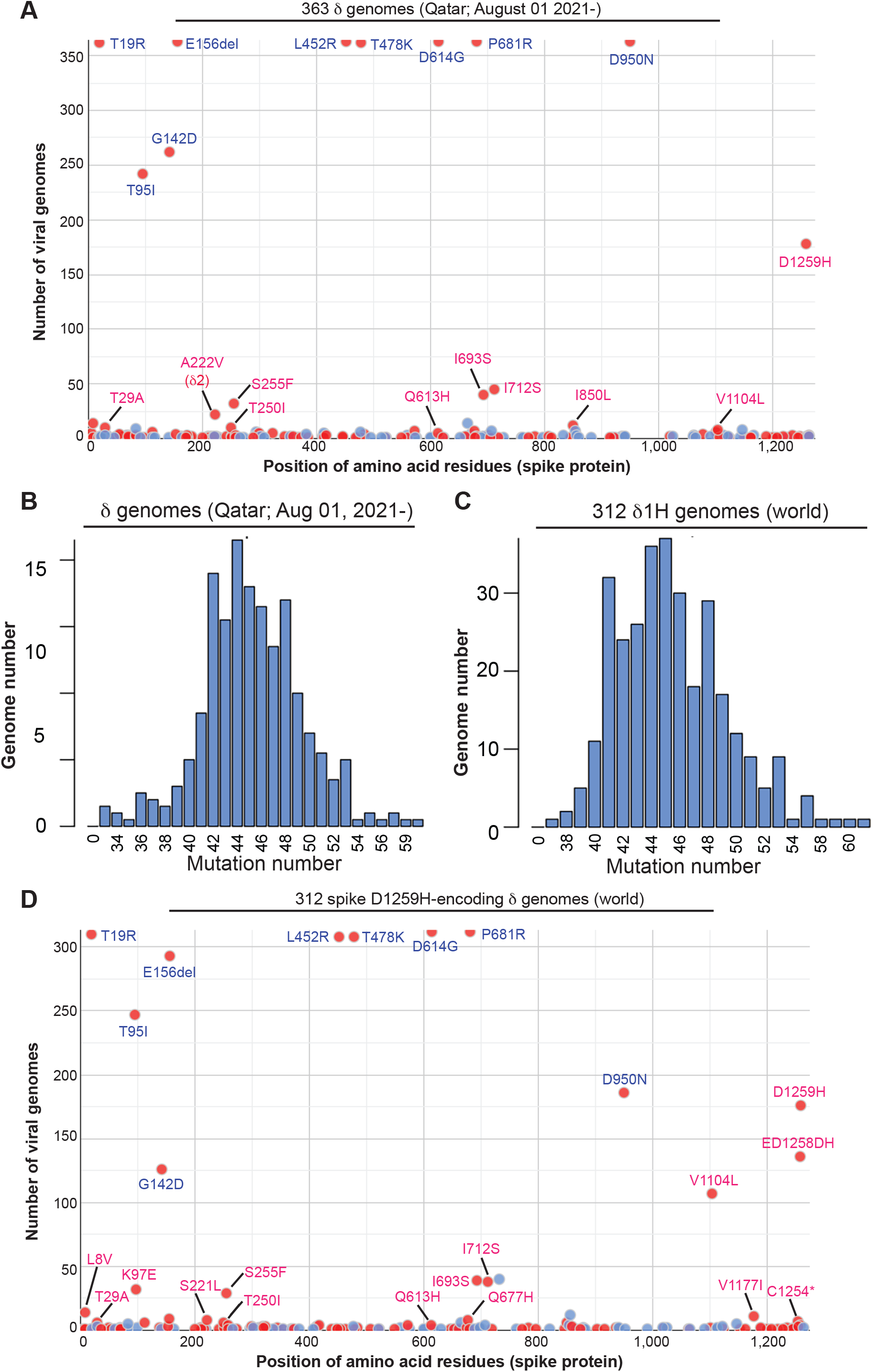
Mutation profiling of δ genomes identified in Qatar and around the world. (**A-B**) Mutation profiling of 363 δ genomes identified in Qatar from August to September 2021. The genomes were downloaded from the GISAID database on October 26, 2021 for mutation profiling via Cronapp. Shown here in (A) and (B) are spike substitutions and the mutation load distribution, respectively. (**C-D**) Mutation profiling of spike D1259H-encoding δ genomes identified in different countries around the world. The genomes were downloaded for mutation profiling as in panel A. Shown here in (C) and (D) are spike substitutions and the mutation load distribution, respectively.

Among the 363 genomes, 339 were identified in August 2021 and 24 were September. Strikingly, almost all 24 genomes encode D1259H along with an extra NSP15 substitution, D132Y (data not shown). In addition, several other spike substitutions, S255F, I693S, I712S and V1104L, are associated with subpopulations of δ1H genomes (Fig. 5A). Among them, S255F is present in almost all 24 genomes from September. S255 is at the C-terminal end of a loop composed of residues 244-246, which are often deleted in other variants, so S255F may confer a similar evolutionary advantage as deletion of the loop itself. I693S and I712S are associated with each other in a subpopulation of about 40 genomes (Fig. 5A). Moreover, they are present in 6 genomes that also encode S255F. Among the 6 genomes, 5 of them were identified in September 2021. Alarmingly, these 6 genomes possess an average mutation load of 50 mutations per genome. In comparison, the average mutation load for the 363 genomes is 44-45 mutations per genome (Fig. 5B). This is still relatively high compared to other δ genomes identified around the same time [15-17,20].

To determine the origin of δ1H, I asked whether there are similar genomes identified in other countries. Related to this, there are 312 such genomes in the GISAID database. The initial genome was identified in Czech Republic in March 2021. In addition to Qatar, 128 are from Mexico and the rest are mainly from Europe. Mutation analysis of these 312 genomes showed an average of 45 mutations per genome (Fig. 5C), which is very similar to the value corresponding to those from Qatar (Fig. 5B). Unexpectedly, there is a large subpopulation (∼130 genomes) encoding an extra spike substitution E1258D, adjacent to D1259H (Fig. 5D). Over 100 genomes in this subpopulation carry another spike substitution, V1104L (Fig. 5D). This is a recurrent substitution and has been found in other variants. V1104 forms a hydrophobic core with I1114 and I1115, so V1104L may improve the interaction. About 30 of them also encodes K97E and a third of these genomes carry an additional spike substitution, V1177I (Fig. 5D). V1177I is adjacent to V1176F, a signature substitution of γ variant [20], and may confer evolutionary advantage. Thus, δ1 subvariant has evolved and yielded δ1H, which has evolved further and generated additional sublineages.

### A δ2 sublineage and δ1 subvariant as major pandemic drivers in Bahrain

Distinct δ subvariant profiles in Israel and Qatar led me to carry out a survey of the situation in other Middle East countries. In the GISAID database (accessed from October 27 to 30, 2021), few δ genomes are available from Iran, Iraq, Saudi Arabia, United Arabia Emirates and Yemen. There are 167 δ genomes from Jordan and 165 of them belong to δ1. Strikingly, 104 encodes spike S255F, indicating that this S255F-encoding δ1 sublineage is the pandemic driver in the country. There are 80 and 159 δ genomes from Lebanon and Oman, respectively, but mutation profiling yielded typical δ subvariant composition observed in India. By contrast, there are many more δ genomes (321) available from Bahrain. Moreover, these genomes show a distinct feature (see below). In addition, like Israel and Qatar, a vaccination program has been rolled out competitively. Thus, I analyzed these genomes from Bahrain in details.

As shown in Fig. 6A, there have been multiple waves of COVID-19 cases in Bahrain, with the latest peaked on May 29, 2021 and being the worst. The initial portion of the wave was due to α variant, whereas the later part is because of δ variant (Fig. 1A-B). Different from Israel and Qatar, Bahrain rolled out its initial 2-dose vaccination program relying heavily on the inactivated virus vaccine from Sinopharm. However, mRNA-based vaccine was also adopted as boosters. The pace of vaccination is more similar to that in Qatar than it was in Israel. By the end of June 2021, its population vaccination rate reached 56.5%. Coincident with the progress of vaccination, the wave dropped sharply after its peak on May 29, 2021 and reached the bottom at the beginning of July 2021 (Fig. 6A). Since then, there have been no major new waves of cases, which may in part due to the booster program that started in the country in June 2021.

**Figure 6.**
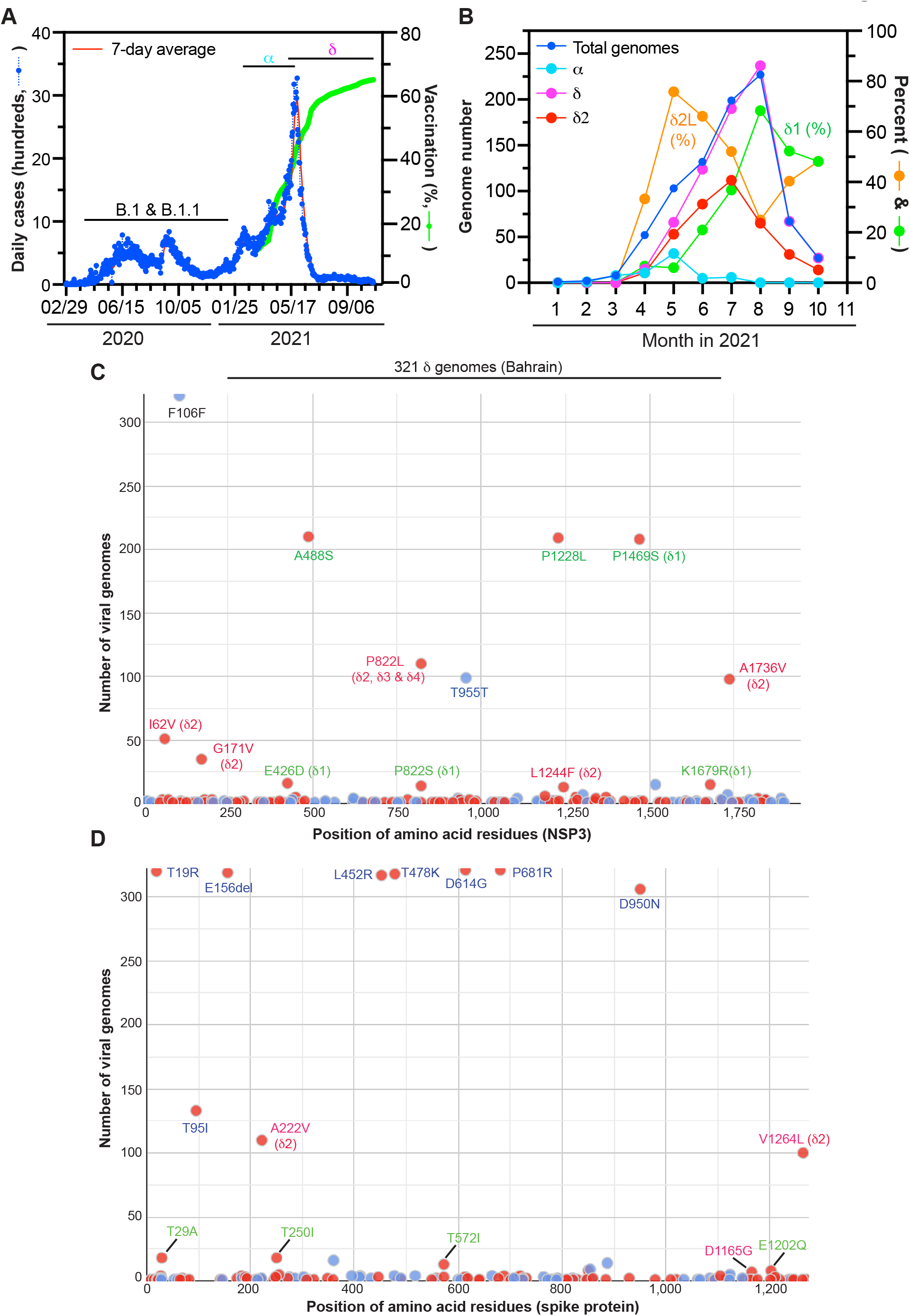
SARS-COV-2 evolution drives the pandemic in Bahrain. (**A**) Epidemiological curve and vaccination progress in the country. Both α and δ variants are responsible for the latest wave. For preparation of this panel, the Our World in Data website was accessed on October 29, 2021. (**B**) Monthly distribution of SARS-COV-2 variant genomes detected in Bahrain. For preparation of the panel, the GASAID database was accessed on October 29, 2021. (**C-D**) Mutation profiling of 321 δ genomes identified in the country. The genomes were downloaded from the GISAID database on October 27, 2021 for mutation profiling via Cronapp. Shown here in (A) and (B) are substitutions in NSP3 and spike proteins, respectively. Note that spike T95I is associated with δ1 subvariant and spike G142D is not encoded by the genomes.

To determine how δ subvariants may have contributed to the surging wave of COVID-19 cases and affected effectiveness of the vaccination program in Bahrain, I carried out mutation profiling of 321 δ genomes identified in the country. As shown in Fig. 6C, 70% of the genomes encodes δ1 subvariant and 30% carries P822L of NSP3. P822L is specific to δ2, δ3 and δ4 [15-17]. A similar number of genomes encode spike A222V (Fig. 6D), so the P822L-encoding genomes are due to δ2 subvariant. Strikingly, an extra NSP3 substitution, A1736V, and an extra spike substitution, V1264L, are present in about 100 genomes (Fig. 6C-D). Importantly, these two substitutions are encoded in the same set of δ2 genomes, suggesting that these genomes correspond to a new variant, referred to δ2L, where the letter L denotes L1264. A similar δ2 sublineage has been described previously [16]. One difference is that the NSP3 substitution is absent in the one described previously [16]. In the GISAID database (accessed on Oct 27, 2021), there are 490 δ2L genomes that also encode A1736V of NSP3.

As shown in Fig. 6B, δ2L was predominant in Bahrain during May 2021 when the COVID-19 wave was surging. Thus, this sublineage was the main pandemic driver in the country. It was still dominant in June and July 2021, but δ1 subvariant started to rise in June and became dominant in August 2021. The apparent decline in September and October might be related to insufficient genomes for a convincing conclusion (Fig. 6B). Thus, δ2L was a major pandemic driver in May and June 2021, but δ1 subvariant has gradually gained dominance since then. These results also suggest that despite its extra spike and NSP3 substitutions (Fig. 6C-D), δ2L is still less virulent than δ1 subvariant. This conclusion is in agreement with what was observed with δ2L in the USA [17].

### Function finetuning by spike substitutions in new variants from Israel, Qatar and Bahrain

As shown in Fig. 7A, spike protein is composed of multiple domains: N-terminal domain, receptor-binding domain, fusion peptide, fusion peptide proximal region, heptad-repeat regions 1 and 2, transmembrane motif and C-terminal cytoplasmic tail [21]. I693S is close to the furin-cleave site (Fig. 7A). Structurally I693 forms hydrophobic interaction with V656, so I693S may disrupt this interaction and affects the local region around the furin cleavage site. I712 is part of a hydrophobic core formed with F1075, V1094 and V1096 of S2 domain, so I712S may destabilize this core. Thus, I693S and I712S in δ1H2 from Qatar (Figs 5C & 7F) may synergize with each other.

**Figure 7.**
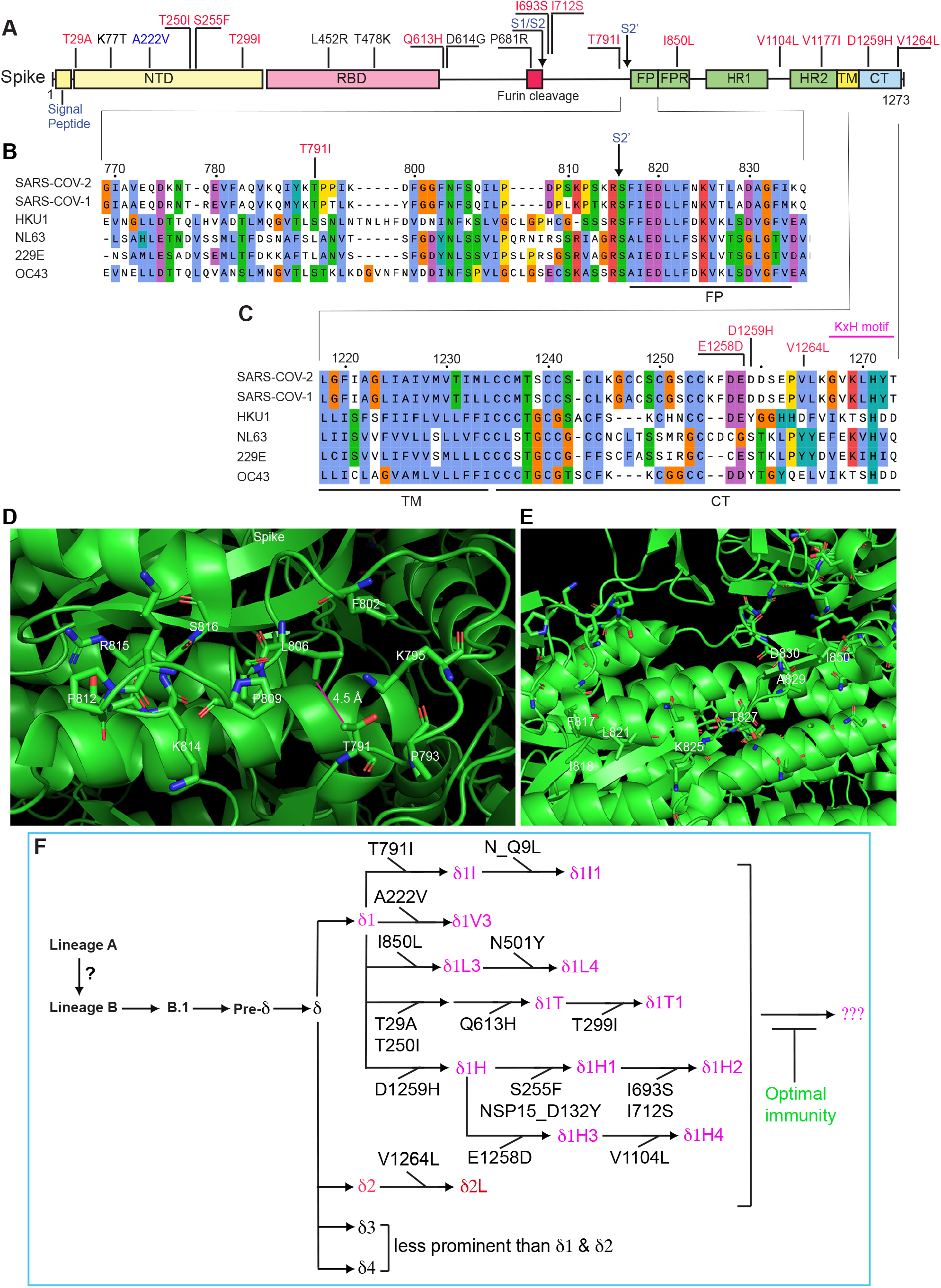
Mechanistic impact of spike substitutions and a model about SARS-COV-2 evolution. (**A**) Domain organization of spike protein. Some key substitutions are shown in black and new substitutions to be described herein are in red. NTD, N-terminal domain; RBD, receptor-binding domain; S1/S2, boundary of S1 and S2 domains after furin cleavage between residues R685 and S686; FP, fusion peptide; FPR, fusion peptide proximal region; HR1 and HR2, heptad-repeat regions 1 and 2, respectively; S2’, cleavage site between residues R815 and S816 in S2 domain; TM, transmembrane motif; CT, C-terminal cytoplasmic tail. The domain organization was adapted from a published study [21]. (**B-C**) Sequence alignment of spike proteins from SARS-COV-2 and five other coronaviruses. The fusion peptide and its N-terminal region are shown in panel B, whereas the cytoplasmic tail is displayed along with a portion of the transmembrane domain in panel C. (**D**) Structural details on spike T791I. T791 is close to the S2’ cleavage site between R815 and S816 (A). The side chain of T791 is 4.5 Å away from L806, so T791I may confer hydrophobic interaction between L806 and I791. (**E**) Structural details on spike I850L. The side chain of I850 is 4.8 Å away from that of A829, so I850L may improve hydrophobic interaction between them. A829 is part of the fusion peptide (A-B), so I850L may finetune the cell fusion ability. Panels C-D are based on PyMol presentation of 6XRA from the PDB database. (**F**) Cartoon showing the relationship of δ and its subvariants. Four subvariants (δ1, δ2, δ3 and δ4) were initially identified in India. δ1 acquires spike T791I and yields δ1I. When it was identified in Israel in June 2021, it was a major δ1 sublineage but its ratio has declined since then (Fig. 1D). In Europe, δ1I acquires Q9L of nucleocapsid protein and gives rise to δ1I1, but this new lineage has not been identified in Israel. δ2L is a major variant in Bahrain but appears to be less competitive than δ1 (Fig. 7B). δ1H emerged suddenly as a key variant in Qatar in July 2021 and its sublineage δ1H1 accounts almost all 24 genomes identified in Septemeber 2021. Moreover, some of the genomes also encode spike I693S and I712S. The resulting lineage carries four spike substitutions and one NSP15 substitution compared to δ1 itself. Thus, δ1H and its sublineages may be the most dangerous variants that have been identified in the Middle East. Despite these different δ subvariants in in Israel, Qatar and Bahrain, the recent successful pandemic control in the countries suggests that optimal immunity through 2-dose vaccination and a subsequent booster constitutes an effective strategy. This continuous branching model is adapted from what has been presented [15-17,20,34].

A recent study showed that fusion peptide proximal region is critical for virus-cell fusion [21]. T791 is close the S2’ cleavage site between R815 and S816 (Fig. 7B). The side chain of T791 is 4.5 Å away from L806, so T791I may confer hydrophobic interaction between L806 and I791 (Fig. 7D). However, it remains unclear whether δ1I is more virulent than δ1 itself. This sublineage does not appear so at least in Israel (Fig. 1D), where vaccination had reached to a high level in June 2021 (Fig. 1A). It remains a possibility that δ1I is more virulent than δ1 itself with vaccination-naïve population.

There are some genomes encoding I850L (Fig. S3B). This is recurrent and has been identified in many other variants, suggesting its importance. I850 is located within the fusion peptide proximal region (Fig. 7A-B). The side chain is 4.8 Å away from A829 of fusion peptide, so I850L may improve the hydrophobic interaction between L850 and A829 (Fig. 7E). Thus, the I850L-encoding sublineage (referred to as δ1L3) may also be more virulent than δ1 itself. Of relevance, this sublineage accounts for a significant portion of δ genomes identified in Turkey, where δ1L3 has acquired spike N501Y and yielded a new sublineage (Fig. 7F). N501Y is shared by α, β and γ variants of concern) [2-4], but absent in δ variant itself and δ [5] (Fig. 6A). Moreover, N501Y is known to improve interaction of the original SARS-COV-2 spike protein with the ACE2 receptor [22]. Thus, it will be important to assess whether this substitution also confers advantage to δ1 subvariant.

V1264 is located at the C-terminal cytoplasmic tail (Fig. 7C). This tail is important for cytoplasmic trafficking [23,24]. Moreover, the cytoplasmic tail facilitates the expression of spike protein on cell surface and promotes syncytia formation between infected cells [25]. A described previously [17], V1264L confers an acidic dileucine motif to spike protein because residues 1157-1262 are acidic and residue 1265 is leucine (Fig. 7C). Such a motif may affect endocytosis of many membrane-associated receptors of the resulting protein [26-28] and mediates clathrin- and AP2-dependent endocytosis of the HIV-1 envelope protein [29]. Like V1264L, both E1258D and D1259H are located at the cytoplasmic tail (Fig. 7C). Both affect an diacidic motif and may also affect spike protein trafficking [30]. Thus, E1258D, D1259H and V1264L may finetune the function of spike protein by altering its cytoplasmic tail.

Shown in Fig. 7F is a cartoon about the relationship of δ and its subvariants. Four subvariants (δ1, δ2, δ3 and δ4) were initially identified in India [15]. δ1 has acquired spike T791I and yielded δ1I, which was initially identified in April 2021 in Texas, USA. In Europe, δ1I acquires Q9L of nucleocapsid protein and gives rise to δ1I1, but this new lineage has not been identified in Israel. δ1 has evolved and yielded δ1H by acquiring spike D1259H (Fig. 7F). Moreover, this sublineage has evolved further and generated additional lineages (Fig. 7F). Based on what observed in Qatar and other countries (Fig. 5), δ1H and its sublineages are alarming. They are perhaps much more alarming than δ1I identified in Israel and δ1L uncovered in Bahrain. Among the three countries, the δ subvariant composition in June 2021 was perhaps the worst in Israel and the most favorable in Bahrain (Figs 1D, 3D & 6B). Notably, SARS-COV-2 genomic information is much more limited from other countries in the Middle East. Thus, it is important to enhance genomic surveillance and leverage it as the guiding light to combat this constantly evolving virus.

### Multiple factors contribute to the different pandemic outcomes in Israel and Qatar

δ1 subvariant became predominant in Israel but not Qatar in June 2021. Another difference is that starting in June 2021, a δ1 sublineage (δ1I) encoding spike T791I was identified in Israel but not in Qatar. The δ1I fraction was 31.8% of all δ genomes uncovered in June 2021 (Fig. 1D). In addition to these two differences, vaccination was rolled out more slowly in Qatar (Fig. 1A & 4A), so by the time δ variant invaded both countries from April to June 2021, the population immunity level in Qatar might have not waned as it had in Israel. If so, this is a matter of luck as no one would have predicted such a coincidence. The fourth difference is that only Pfizer/BioNtech vaccine has been administrated in Israel [18,19], whereas Moderna vaccine has also been deployed in Qatar [31]. This vaccine confers immunity that does not decline as quickly as the Pfizer/BioNtech one [32]. The fifth difference is that the eventual vaccinated population has reached to a much higher level in Qatar (Figs 1A & 4A). The last possibility is whether there has been behavioral difference in the two populations in Israel and Qatar when public health measures were administrated. Therefore, multiple factors may have contributed to the different pandemic outcomes in the two countries. Importantly, the booster program in Israel has successfully controlled the recent surging wave [18,19], thereby proving an effective remedy to control δ1 and its sublineages. Of relevance, boosters might have also kept the pandemic control effective in Bahrain since July 2021. Therefore, achieving optimal immunity is an effective strategy to block δ evolution and end the pandemic as swiftly as possible (Fig. 7F).

## Supporting information

Acknowledgement table S1 for the GISAID SARS-COV-2 genomes used in this study

Acknowledgement table S2 for the GISAID SARS-COV-2 genomes used in this study

Acknowledgement table S3 for the GISAID SARS-COV-2 genomes used in this study

## Data Availability

All data produced in the present study are available upon reasonable request to the authors

## ACKNOWLEDGEMENT

I gratefully acknowledge the GISAID for diligent and tireless maintenance SARS-COV-2 genomes and numerous investigators for the valuable genome sequences used in this work (see the supplementary section for details). I am also grateful to Professor Federico M. Giorgi at University of Bologna, Italy, for developing the Coronapp COVID-19 genome annotator and generously allowing me timely access to the server. This work was supported by funds from Canadian Institutes of Health Research (CIHR), Natural Sciences and Engineering Research Council of Canada (NSERC) and Compute Canada (to X.J.Y.).

## MATERIALS AND METHODS

### SARS-COV-2 genome sequences, mutational profiling and phylogenetic analysis

The genomes were downloaded the GISAID database (https://www.gisaid.org/) as specified in figure legends. CoVsurver (https://www.gisaid.org/epiflu-applications/covsurver-mutations-app/) was used to analyze mutations on representative SARS-COV-2 genomes. Fasta files containing specific groups of genomes were downloaded from the GISAID database. During downloading, each empty space in the Fasta file headers was replaced by an underscore because such a space makes the files incompatible for subsequent mutational profiling, sequence alignment and phylogenetic analysis, as described with details in two other studies [15,33]. The Fasta headers were shortened and modified further as described [15,33]. The cleaned Fasta files were used for mutational profiling via Coronapp (http://giorgilab.unibo.it/coronannotator/), a web-based mutation annotation application [12,13]. Two cleaned Fasta files were also uploaded onto SnapGene (version 5.3.2) for multisequence alignment via the MAFFT tool, and RAxML-NG version 0.9.0 [14] was used for subsequent phylogenetic analysis as described [15].

### Defining different variant genomes using various markers

α, β, γ, δ and other variant genomes were downloaded from the GISAID database as defined by the server. δ subvariant genomes were defined as described [15]. Briefly, nucleocapsid substitutions G215C and R385K were used as markers for δ1 or δ3 genomes, respectively. Spike substitutions A222V and K77T were used as markers for δ2 or δ4 genomes, respectively. In Europe, there are many δ1V genomes that also encode spike A222V, so the NSP3 substitution P822L was used together with spike A222V to identify δ2 genomes. As discussed previously [15], there are several limitations with these markers. But the limitations should not affect the overall conclusions.

### PyMol structural modeling

The PyMol molecular graphics system (version 2.4.2, https://pymol.org/2/) from Schrödinger, Inc. was used for downloading structure files from the PDB database for further analysis and image generation. Structural images were cropped via Adobe Photoshop for further presentation through Illustrator.

### Pandemic and vaccination data

Pandemic and vaccination data were downloaded from the Our World in Data website (https://ourworldindata.org/explorers/coronavirus-data-explorer?zoomToSelection=true&time=2020-03-01..latest&facet=none&pickerSort=desc&pickerMetric=total_cases&Metric=Confirmed+cases&Interval=Cumulative&Relative+to+Population=false&Align+outbreaks=false&country=∼OWID_WRL) as a .csv file for further processing via Excel and figure generation through Prism 9.0 and Adobe Illustrator.

## SUPPLEMENTAL INFORMATION

This section includes 1) three supplementary figures with detailed information on the two phylogenetic trees and 2) four acknowledgement tables for the GISAID genomes used in this work.

## SUPPLEMENTAL FIGURE LEGENDS

**Figure S1.**
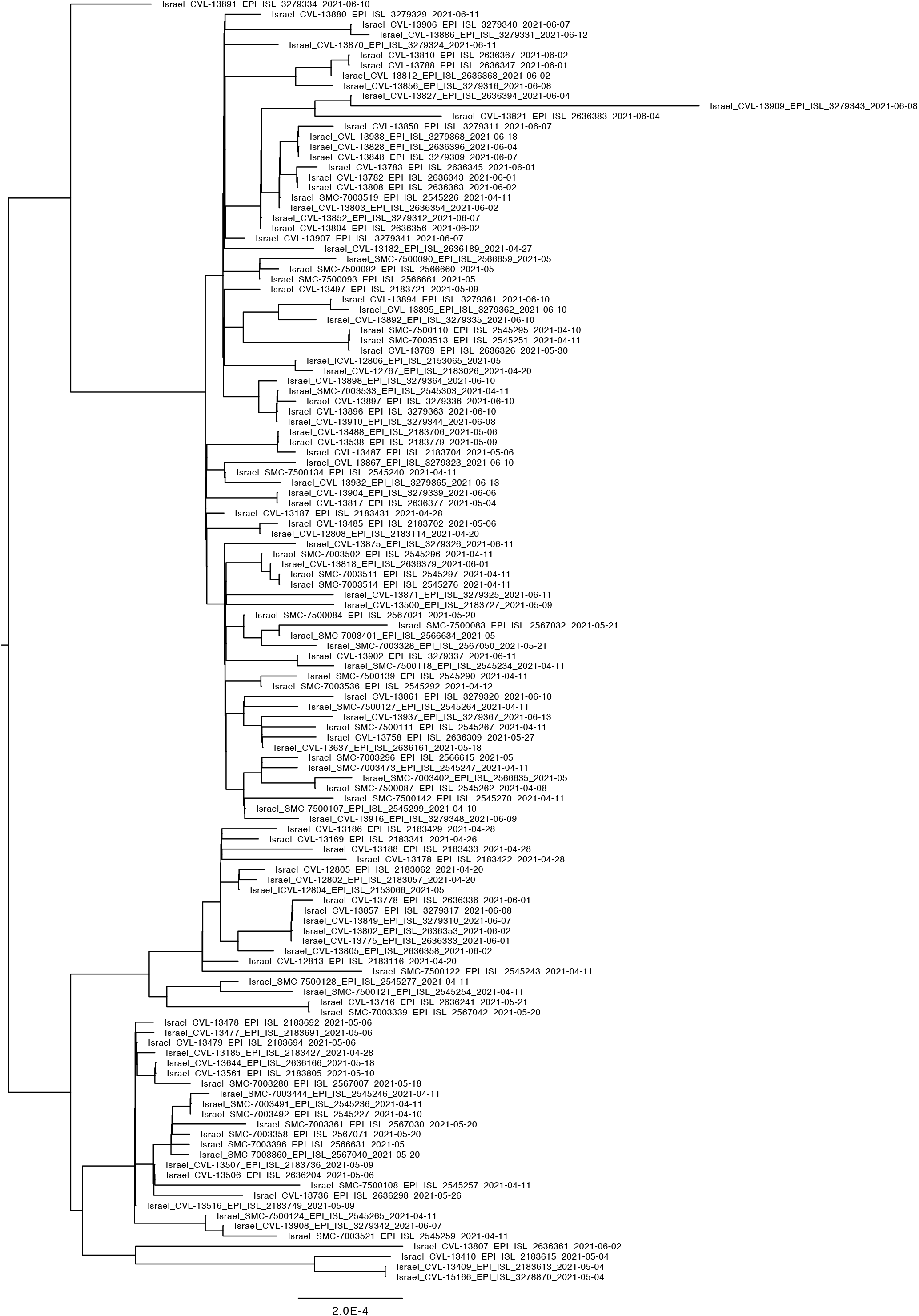
Phylogenetic analysis of 140 δ genomes identified in Israel by June 15, 2021. The analysis was carried out via the package RAxML-NG, to generate 20 maximum likelihood trees and one bestTree for presentation via Figtree. The strain names and GISAID accession numbers of the genomes are detailed here, with an abbreviated version of three shown in Fig. 3D. Note that during RAxML-NG analysis, when identical genomes were identified, only one such genome was left for analysis and the rest were automatically removed, so not all 140 genomes are shown on the tree presented here. An abbreviated version of this tree is shown in Fig. 1C.

**Figure S2.**
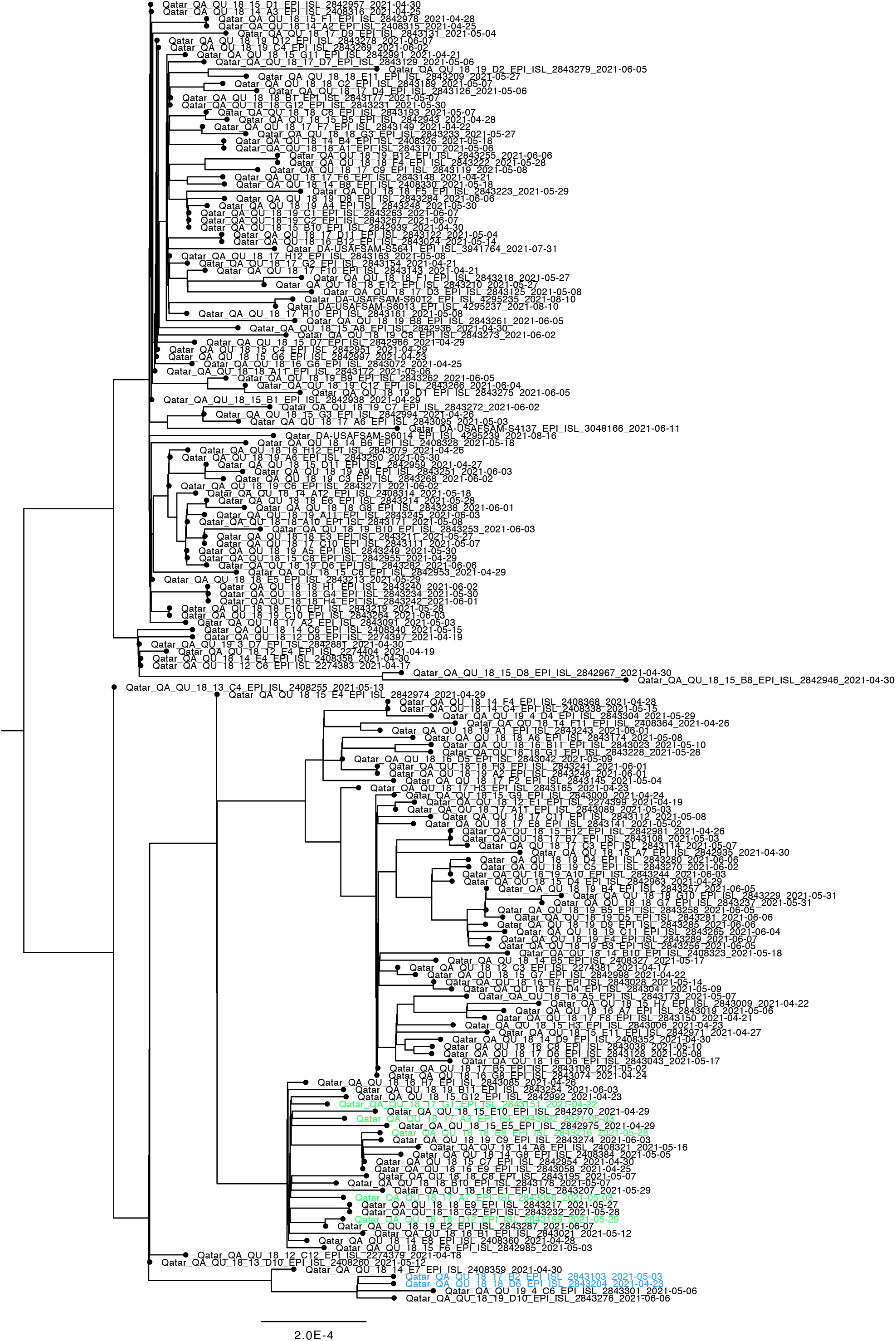
Phylogenetic analysis of 182 δ genomes identified in Qatar. The analysis was carried out as in Fig. 4C, with the strain names and GISAID accession numbers of the genomes detailed here. Note that during RAxML-NG analysis, when identical genomes were identified, only one such genome was left for analysis and the rest were automatically removed, so not all 182 are shown on the tree presented here.

**Figure S3.**
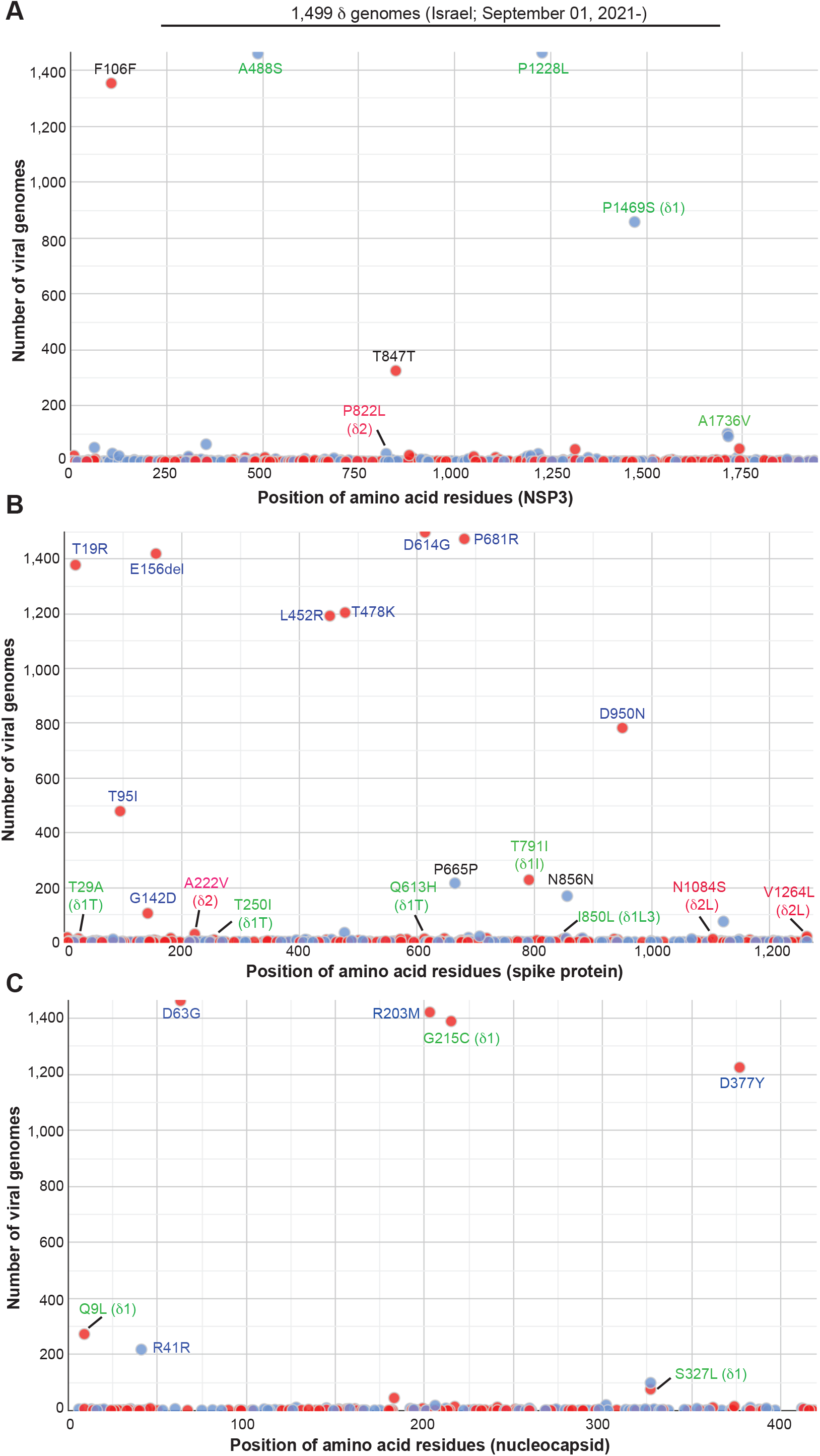
Mutation profiling of the 1,499 δ genomes identified in Israel from September to October 2021. The genomes were downloaded from the GISAID database for mutation profiling as in Fig. 2D. Shown here in (A), (B) and (C) are substitutions in NSP3, spike and nucleocapsid proteins, respectively.

## REFERENCES

1. Rambaut, A., E.C. Holmes, A. O’Toole, V. Hill, J.T. McCrone, C. Ruis, L. du Plessis, and O.G. Pybus. (2020). A dynamic nomenclature proposal for SARS-CoV-2 lineages to assist genomic epidemiology. Nat Microbiol 5, 1403–1407.

2. Volz, E., S. Mishra, M. Chand, J.C. Barrett, R. Johnson, L. Geidelberg, W.R. Hinsley, D.J. Laydon, G. Dabrera, A. O’Toole, R. Amato, M. Ragonnet-Cronin, I. Harrison, B. Jackson, C.V. Ariani, O. Boyd, N.J. Loman, J.T. McCrone, S. Goncalves, D. Jorgensen, R. Myers, V. Hill, D.K. Jackson, K. Gaythorpe, N. Groves, J. Sillitoe, D.P. Kwiatkowski, C.-G.U. consortium, S. Flaxman, O. Ratmann, S. Bhatt, S. Hopkins, A. Gandy, A. Rambaut, and N.M. Ferguson. (2021). Assessing transmissibility of SARS-CoV-2 lineage B.1.1.7 in England. Nature, epub.

3. Tegally, H., E. Wilkinson, M. Giovanetti, A. Iranzadeh, V. Fonseca, J. Giandhari, D. Doolabh, S. Pillay, E.J. San, N. Msomi, K. Mlisana, A. von Gottberg, S. Walaza, M. Allam, A. Ismail, T. Mohale, A.J. Glass, S. Engelbrecht, G. Van Zyl, W. Preiser, F. Petruccione, A. Sigal, D. Hardie, G. Marais, N.Y. Hsiao, S. Korsman, M.A. Davies, L. Tyers, I. Mudau, D. York, C. Maslo, D. Goedhals, S. Abrahams, O. Laguda-Akingba, A. Alisoltani-Dehkordi, A. Godzik, C.K. Wibmer, B.T. Sewell, J. Lourenco, L.C.J. Alcantara, S.L. Kosakovsky Pond, S. Weaver, D. Martin, R.J. Lessells, J.N. Bhiman, C. Williamson, and T. de Oliveira. (2021). Detection of a SARS-CoV-2 variant of concern in South Africa. Nature 592, 438–443.

4. Faria, N.R., et al. (2021). Genomics and epidemiology of the P.1 SARS-CoV-2 lineage in Manaus, Brazil. Science 372, 815–821.

5. Mlcochova, P., et al. (2021). SARS-CoV-2 B.1.617.2 Delta variant replication and immune evasion. Nature, epub.

6. Elbe, S. and G. Buckland-Merrett. (2017). Data, disease and diplomacy: GISAID’s innovative contribution to global health. Glob Chall 1, 33–46.

7. You, L., L. Li, K. Yan, J. Zou, J. Belle, A. Nijnik, E. Wang, and X.J. Yang. (2016). BRPF1 is essential for development of fetal hematopoietic stem cells. J Clin Investig 126, 3247–3262.

8. Yan, K., J. Rousseau, R.O. Littlejohn, C. Kiss, A. Lehman, J.R. Rosenfeld, C.T.R. Stumpel, A.P. Stegmann, L. Robak, F. Scaglia, T.T. Nguyen, H. Fu, N.F. Ajeawung, M.V. Camurri, L. Li, A. Gardham, B. Panis, M. Almannai, M.J. Sacoto, B. Baskin, C. Ruivenkamp, D. Study, C. Study, M.T. Cho, T. Potjer, G.W. Santen, M.J. Parker, N. Canham, M. McKinnon, L. Potocki, J. MacKenzie, E.R. Roeder, P.M. Campeau, and X.J. Yang. (2017). Mutations in the chromatin regulator gene BRPF1 causes syndromic intellectual disability and deficient histone acetylation. Am. J. Hum. Genet. 100, 91–104.

9. Li, L., M. Ghorbani, M.W. Hubshman, J. Rousseau, I. Thiffault, R.E. Schnur, C. Breen, R. Oegema, J.M. Weiss, Q. Waisfisz, S. Welner, H. Kingston, E.M. Boon, L. Basel-Salmon, O. Konen, H. Goldberg, L. Bazaq, S. Tzur, X. Bi, M. Bruccoleri, K. McWalter, M. Cho, M. Scarano, S.S. Brooks, S.S. Hughes, K.L.I. van Gassen, J.M. van Hagen, T.K. Pandita, P.B. Agrawal, P.M. Campeau, and X.J. Yang. (2020). Lysine acetyltransferase 8 is involved in cerebral development and syndromic intellectual disability. J. Clin. Investig. 130, 1431–1445.

10. Yan, K., J. Rousseau, K. Machol, L.A. Cross, K.E. Agre, C.F. Gibson, A. Goverde, K.L. Engleman, H. Verdin, E. Baere, L. Potocki, D. Zhou, M. Cadieux-Dion, G.A. Bellus, M.D. Wagner, R.J. Hale, N. Esber, A.F. Riley, B.D. Solomon, M.T. Cho, H. Porter, B.C. Lanpher, A.M. Lewis, J. Savatt, I. Thiffault, B. Callewaert, P.M. Campeau, and X.J. Yang. (2020). Deficient histone H3 propionylation by BRPF1-KAT6 complexes in neurodevelopmental disorders and cancer. Sci. Adv. 6, eaax0021.

11. Yang, X.J. (2005). Multisite protein modification and intramolecular signaling. Oncogene 24, 1653–1662.

12. Mercatelli, D., L. Triboli, E. Fornasari, F. Ray, and F.M. Giorgi. (2021). Coronapp: A web application to annotate and monitor SARS-CoV-2 mutations. J Med Virol 93, 3238–3245.

13. Mercatelli, D. and F.M. Giorgi. (2020). Geographic and Genomic Distribution of SARS-CoV-2 Mutations. Front Microbiol 11, 1800.

14. Kozlov, A.M., D. Darriba, T. Flouri, B. Morel, and A. Stamatakis. (2019). RAxML-NG: a fast, scalable and user-friendly tool for maximum likelihood phylogenetic inference. Bioinformatics 35, 4453–4455.

15. Yang, X.J. (2021). SARS-COV-2 delta variant drives the pandemic in India and Europe through two subvariants. medRxiv, https://www.medrxiv.org/content/10.1101/2021.10.16.21265096v1.

16. Yang, X.J. (2021). SARS-COV-2 delta variant drives the pandemic in the USA through two subvariants. Research Square, https://www.researchsquare.com/article/rs-986605/v1.

17. Yang, X.J. (2021). Delta-1 variant of SARS-COV-2 acquires spike V1264L and drives the pandemic in Indonesia, Singapore and Malaysia. Research Square, https://www.researchsquare.com/article/rs-999390/v1.

18. Bar-On, Y.M., Y. Goldberg, M. Mandel, O. Bodenheimer, L. Freedman, S. Alroy-Preis, N. Ash, A. Huppert, and R. Milo. (2021). Protection Across Age Groups of BNT162b2 Vaccine Booster against Covid-19. medRxiv, https://www.medrxiv.org/content/10.1101/2021.10.07.21264626v1.

19. Bar-On, Y.M., Y. Goldberg, M. Mandel, O. Bodenheimer, L. Freedman, N. Kalkstein, B. Mizrahi, S. Alroy-Preis, N. Ash, R. Milo, and A. Huppert. (2021). Protection of BNT162b2 Vaccine Booster against Covid-19 in Israel. N Engl J Med 385, 1393–1400.

20. Yang, X.J. (2021). Delta-1 variant of SARS-COV-2 acquires spike V1176F and yields a highly mutated subvariant in Europe. bioRxiv, https://www.biorxiv.org/content/10.1101/2021.10.16.463825v2.

21. Cai, Y., J. Zhang, T. Xiao, C.L. Lavine, S. Rawson, H. Peng, H. Zhu, K. Anand, P. Tong, A. Gautam, S. Lu, S.M. Sterling, R.M. Walsh, Jr., S. Rits-Volloch, J. Lu, D.R. Wesemann, W. Yang, M.S. Seaman, and B. Chen. (2021). Structural basis for enhanced infectivity and immune evasion of SARS-CoV-2 variants. Science 373, 642–648.

22. Starr, T.N., A.J. Greaney, S.K. Hilton, D. Ellis, K.H.D. Crawford, A.S. Dingens, M.J. Navarro, J.E. Bowen, M.A. Tortorici, A.C. Walls, N.P. King, D. Veesler, and J.D. Bloom. (2020). Deep Mutational Scanning of SARS-CoV-2 Receptor Binding Domain Reveals Constraints on Folding and ACE2 Binding. Cell 182, 1295–1310 e20.

23. Sadasivan, J., M. Singh, and J.D. Sarma. (2017). Cytoplasmic tail of coronavirus spike protein has intracellular targeting signals. J Biosci 42, 231–244.

24. Jennings, B.C., S. Kornfeld, and B. Doray. (2021). A weak COPI binding motif in the cytoplasmic tail of SARS-CoV-2 spike glycoprotein is necessary for its cleavage, glycosylation, and localization. FEBS Lett 595, 1758–1767.

25. Cattin-Ortola, J., L.G. Welch, S.L. Maslen, G. Papa, L.C. James, and S. Munro. (2021). Sequences in the cytoplasmic tail of SARS-CoV-2 Spike facilitate expression at the cell surface and syncytia formation. Nat Commun 12, 5333.

26. Doray, B., J.M. Knisely, L. Wartman, G. Bu, and S. Kornfeld. (2008). Identification of acidic dileucine signals in LRP9 that interact with both GGAs and AP-1/AP-2. Traffic 9, 1551–62.

27. Kelly, B.T., A.J. McCoy, K. Spate, S.E. Miller, P.R. Evans, S. Honing, and D.J. Owen. (2008). A structural explanation for the binding of endocytic dileucine motifs by the AP2 complex. Nature 456, 976–979.

28. Tikkanen, R., S. Obermuller, K. Denzer, R. Pungitore, H.J. Geuze, K. von Figura, and S. Honing. (2000). The dileucine motif within the tail of MPR46 is required for sorting of the receptor in endosomes. Traffic 1, 631–40.

29. Byland, R., P.J. Vance, J.A. Hoxie, and M. Marsh. (2007). A conserved dileucine motif mediates clathrin and AP-2-dependent endocytosis of the HIV-1 envelope protein. Mol Biol Cell 18, 414–25.

30. Lin, Y.C., B.M. Chen, W.C. Lu, C.I. Su, Z.M. Prijovich, W.C. Chung, P.Y. Wu, K.C. Chen, I.C. Lee, T.Y. Juan, and S.R. Roffler. (2013). The B7-1 cytoplasmic tail enhances intracellular transport and mammalian cell surface display of chimeric proteins in the absence of a linear ER export motif. PLoS One 8, e75084.

31. Chemaitelly, H., P. Tang, M.R. Hasan, S. AlMukdad, H.M. Yassine, F.M. Benslimane, H.A. Al Khatib, P. Coyle, H.H. Ayoub, Z. Al Kanaani, E. Al Kuwari, A. Jeremijenko, A.H. Kaleeckal, A.N. Latif, R.M. Shaik, H.F. Abdul Rahim, G.K. Nasrallah, M.G. Al Kuwari, H.E. Al Romaihi, A.A. Butt, M.H. Al-Thani, A. Al Khal, R. Bertollini, and L.J. Abu-Raddad. (2021). Waning of BNT162b2 Vaccine Protection against SARS-CoV-2 Infection in Qatar. N Engl J Med.

32. Self, W.H., et al. (2021). Comparative Effectiveness of Moderna, Pfizer-BioNTech, and Janssen (Johnson & Johnson) Vaccines in Preventing COVID-19 Hospitalizations Among Adults Without Immunocompromising Conditions - United States, March-August 2021. MMWR Morb Mortal Wkly Rep 70, 1337–1343.

33. Yang, X.J. (2021). SARS-COV-2 C.1.2 variant is highly mutated but may exhibit reduced affinity for ACE2 receptor. bioRxiv, https://www.biorxiv.org/content/10.1101/2021.10.16.464644v1.

34. Yang, X.J. (2021). SARS-COV-2 gamma variant acquires spike P681H or P681R for improved viral fitness bioRxiv, https://www.biorxiv.org/content/10.1101/2021.10.16.464641v1.

