## Supplementary material for "δ subvariants of SARS-COV-2 in Israel, Qatar and Bahrain: Optimal vaccination as an effective strategy to block viral evolution and control the pandemic": Acknowledgement table S1 for the GISAID SARS-COV-2 genomes used in this study

We gratefully acknowledge the following Authors from the Originating laboratories responsible for obtaining the specimens, as well as the Submitting laboratories where the genome data were generated and shared via GISAID, on which this research is based.

All Submitters of data may be contacted directly via www.gisaid.org

Authors are sorted alphabetically.

| Accession ID | Originating Laboratory | Submitting Laboratory | Authors |
|---|---|---|---|
| EPI_ISL_4169984, EPI_ISL_4169986 | Central Virology Laboratory, Israel Ministry of Health | Israel Institute for Biological Research | Adi Beth-Din; Anat Zvi; Inbar Cohen-Gihon; Nir Paran; Ofir Israeli; Yfat Yahalom Ronen |
| EPI_ISL_2153065, EPI_ISL_2153066, EPI_ISL_5507782, EPI_ISL_5507783, EPI_ISL_5507784, EPI_ISL_5507785, EPI_ISL_5507786, EPI_ISL_5507787, EPI_ISL_5507788, EPI_ISL_5507803, EPI_ISL_5507809, EPI_ISL_5507818, EPI_ISL_5507823, EPI_ISL_5507828, EPI_ISL_5507836, EPI_ISL_5507839, EPI_ISL_5507848, EPI_ISL_5507853, EPI_ISL_5507859, EPI_ISL_5507873, EPI_ISL_5507884, EPI_ISL_5507887, EPI_ISL_5507894, EPI_ISL_5507902, EPI_ISL_5507907, EPI_ISL_5507913, EPI_ISL_5507920, EPI_ISL_5507925, EPI_ISL_5507928, EPI_ISL_5507935, EPI_ISL_5507949, EPI_ISL_5507954, EPI_ISL_5507963, EPI_ISL_5507968, EPI_ISL_5507974, EPI_ISL_5507975, EPI_ISL_5507976, EPI_ISL_5507977, EPI_ISL_5507978, EPI_ISL_5507979, EPI_ISL_5507980, EPI_ISL_5507981, EPI_ISL_5507982, EPI_ISL_5507983, EPI_ISL_5507984, EPI_ISL_5507985, EPI_ISL_5507986, EPI_ISL_5507987, EPI_ISL_5507988, EPI_ISL_5507989, EPI_ISL_5507990, EPI_ISL_5507991, EPI_ISL_5507992, EPI_ISL_5507993, EPI_ISL_5507994, EPI_ISL_5507995, EPI_ISL_5507996, EPI_ISL_5507997, EPI_ISL_5507998, EPI_ISL_5507999, EPI_ISL_5508000, EPI_ISL_5508001, EPI_ISL_5508002, EPI_ISL_5508003, EPI_ISL_5508004, EPI_ISL_5508005, EPI_ISL_5508006, EPI_ISL_5508007, EPI_ISL_5508008, EPI_ISL_5508009, EPI_ISL_5508010, EPI_ISL_5508011, EPI_ISL_5508015, EPI_ISL_5508016, EPI_ISL_5508017, EPI_ISL_5508018, EPI_ISL_5508019, EPI_ISL_5508020, EPI_ISL_5508021, EPI_ISL_5508022, EPI_ISL_5508023, EPI_ISL_5508024, EPI_ISL_5508025, EPI_ISL_5508026, EPI_ISL_5508027, EPI_ISL_5508028, EPI_ISL_5508029, EPI_ISL_5508030, EPI_ISL_5508031, EPI_ISL_5508032, EPI_ISL_5508033, EPI_ISL_5508034, EPI_ISL_5508035, EPI_ISL_5508036, EPI_ISL_5508037, EPI_ISL_5508038, EPI_ISL_5508039, EPI_ISL_5508040, EPI_ISL_5508041, EPI_ISL_5508042, EPI_ISL_5508043, EPI_ISL_5508044, EPI_ISL_5508045, EPI_ISL_5508046, EPI_ISL_5508047, EPI_ISL_5508048, EPI_ISL_5508049, EPI_ISL_5508050, EPI_ISL_5508051, EPI_ISL_5508052, EPI_ISL_5508053, EPI_ISL_5508054, EPI_ISL_5508055, EPI_ISL_5508056, EPI_ISL_5508057, EPI_ISL_5508058, EPI_ISL_5508059, EPI_ISL_5508060, EPI_ISL_5508061, EPI_ISL_5508062, EPI_ISL_5508063, EPI_ISL_5508064, EPI_ISL_5508065, EPI_ISL_5508066, EPI_ISL_5508067, EPI_ISL_5508068, EPI_ISL_5508069, EPI_ISL_5508070, EPI_ISL_5508071, EPI_ISL_5508072, EPI_ISL_5508073, EPI_ISL_5508074, EPI_ISL_5508078, EPI_ISL_5508079, EPI_ISL_5508080, EPI_ISL_5508081, EPI_ISL_5508082, EPI_ISL_5508083, EPI_ISL_5508084, EPI_ISL_5508085, EPI_ISL_5508086, EPI_ISL_5508087, EPI_ISL_5508088, EPI_ISL_5508089, EPI_ISL_5508090, EPI_ISL_5508091, EPI_ISL_5508092, EPI_ISL_5508093, EPI_ISL_5508094, EPI_ISL_5508095, EPI_ISL_5508096, EPI_ISL_5508097, EPI_ISL_5508098, EPI_ISL_5508099, EPI_ISL_5508100, EPI_ISL_5508101, EPI_ISL_5508102, EPI_ISL_5508103, EPI_ISL_5508104, EPI_ISL_5508105, EPI_ISL_5508106, EPI_ISL_5508107, EPI_ISL_5508108, EPI_ISL_5508109, EPI_ISL_5508110, EPI_ISL_5508111, EPI_ISL_5508112, EPI_ISL_5508113, EPI_ISL_5508114, EPI_ISL_5508115, EPI_ISL_5508116, EPI_ISL_5508117, EPI_ISL_5508118, EPI_ISL_5508119, EPI_ISL_5508120, EPI_ISL_5508121, EPI_ISL_5508122, EPI_ISL_5508123, EPI_ISL_5508124, EPI_ISL_5508125, EPI_ISL_5508126, EPI_ISL_5508127, EPI_ISL_5508128, EPI_ISL_5508129, EPI_ISL_5508130, EPI_ISL_5508131, EPI_ISL_5508132, EPI_ISL_5508136, EPI_ISL_5508137, EPI_ISL_5508138, EPI_ISL_5508139, EPI_ISL_5508140, EPI_ISL_5508141, EPI_ISL_5508142, EPI_ISL_5508143, EPI_ISL_5508144, EPI_ISL_5508145, EPI_ISL_5508146, EPI_ISL_5508147, EPI_ISL_5508148, EPI_ISL_5508149, EPI_ISL_5508150, EPI_ISL_5508151, EPI_ISL_5508152, EPI_ISL_5508153, EPI_ISL_5508154, EPI_ISL_5508155, EPI_ISL_5508156, EPI_ISL_5508157, EPI_ISL_5508158, EPI_ISL_5508159, EPI_ISL_5508160, EPI_ISL_5508161, EPI_ISL_5508162, EPI_ISL_5508163, EPI_ISL_5508164, EPI_ISL_5508165, EPI_ISL_5508166, EPI_ISL_5508167, EPI_ISL_5508168, EPI_ISL_5508173, EPI_ISL_5508174, EPI_ISL_5508175, EPI_ISL_5508176, EPI_ISL_5508177, EPI_ISL_5508178, EPI_ISL_5508179, EPI_ISL_5508180, EPI_ISL_5508181, EPI_ISL_5508182, EPI_ISL_5508183, EPI_ISL_5508184, EPI_ISL_5508185, EPI_ISL_5508186, EPI_ISL_5508187, EPI_ISL_5508188, EPI_ISL_5508189, EPI_ISL_5508190, EPI_ISL_5508191, EPI_ISL_5508192, EPI_ISL_5508193, EPI_ISL_5508194, EPI_ISL_5508195, EPI_ISL_5508196, EPI_ISL_5508197, EPI_ISL_5508198, EPI_ISL_5508199, EPI_ISL_5508200, EPI_ISL_5508201, EPI_ISL_5508202, EPI_ISL_5508203, EPI_ISL_5508205, EPI_ISL_5508206, EPI_ISL_5508207, EPI_ISL_5508208, EPI_ISL_5508209, EPI_ISL_5508210, EPI_ISL_5508211, EPI_ISL_5508212, EPI_ISL_5508213, EPI_ISL_5508214, EPI_ISL_5508215, EPI_ISL_5508216, EPI_ISL_5508217, EPI_ISL_5508218, EPI_ISL_5508219, EPI_ISL_5508220, EPI_ISL_5508221, EPI_ISL_5508222, EPI_ISL_5508223, EPI_ISL_5508224, EPI_ISL_5508225, EPI_ISL_5508226, EPI_ISL_5508227, EPI_ISL_5508228, EPI_ISL_5508229, EPI_ISL_5508230, EPI_ISL_5508231, EPI_ISL_5508232, EPI_ISL_5508233, EPI_ISL_5508234, EPI_ISL_5508235, EPI_ISL_5508236, EPI_ISL_5508237, EPI_ISL_5508238, EPI_ISL_5508239, EPI_ISL_5508240, EPI_ISL_5508241, EPI_ISL_5508242, EPI_ISL_5508243, EPI_ISL_5508244, EPI_ISL_5508245, EPI_ISL_5508246, EPI_ISL_5508247, EPI_ISL_5508248, EPI_ISL_5508249, EPI_ISL_5508250, EPI_ISL_5508251, EPI_ISL_5508252, EPI_ISL_5508253, EPI_ISL_5508254, EPI_ISL_5508255, EPI_ISL_5508256, EPI_ISL_5508257, EPI_ISL_5508258, EPI_ISL_5508259, EPI_ISL_5508260, EPI_ISL_5508261, EPI_ISL_5508262, EPI_ISL_5508263, EPI_ISL_5508264, EPI_ISL_5508265, EPI_ISL_5508266, EPI_ISL_5508267, EPI_ISL_5508268, EPI_ISL_5508269, EPI_ISL_5508270, EPI_ISL_5508271, EPI_ISL_5508272, EPI_ISL_5508273, EPI_ISL_5508274, EPI_ISL_5508275, EPI_ISL_5508279, EPI_ISL_5508280, EPI_ISL_5508281, EPI_ISL_5508282, EPI_ISL_5508283, EPI_ISL_5508284, EPI_ISL_5508285, EPI_ISL_5508286, EPI_ISL_5508287, EPI_ISL_5508288, EPI_ISL_5508289, EPI_ISL_5508290, EPI_ISL_5508291, EPI_ISL_5508292, EPI_ISL_5508293, EPI_ISL_5508294, EPI_ISL_5508295, EPI_ISL_5508296, EPI_ISL_5508297, EPI_ISL_5508298, EPI_ISL_5508299, EPI_ISL_5508300, EPI_ISL_5508301, EPI_ISL_5508302, EPI_ISL_5508303, EPI_ISL_5508304, EPI_ISL_5508305, EPI_ISL_5508306, EPI_ISL_5508307, EPI_ISL_5508308, EPI_ISL_5508309, EPI_ISL_5508310, EPI_ISL_5508311, EPI_ISL_5508312, EPI_ISL_5508313, EPI_ISL_5508314, EPI_ISL_5508315, EPI_ISL_5508316, EPI_ISL_5508317, EPI_ISL_5508318, EPI_ISL_5508319, EPI_ISL_5508320, EPI_ISL_5508321, EPI_ISL_5508322, EPI_ISL_5508323, EPI_ISL_5508328, EPI_ISL_5508329, EPI_ISL_5508330, EPI_ISL_5508331, EPI_ISL_5508332, EPI_ISL_5508333, EPI_ISL_5508334, EPI_ISL_5508335, EPI_ISL_5508336, EPI_ISL_5508337, EPI_ISL_5508338, EPI_ISL_5508339, EPI_ISL_5508340, EPI_ISL_5508341, EPI_ISL_5508342, EPI_ISL_5508343, EPI_ISL_5508344, EPI_ISL_5508345, EPI_ISL_5508346, EPI_ISL_5508347, EPI_ISL_5508348, EPI_ISL_5508349, EPI_ISL_5508350, EPI_ISL_5508351, EPI_ISL_5508352, EPI_ISL_5508353, EPI_ISL_5508354, EPI_ISL_5508355, EPI_ISL_5508356, EPI_ISL_5508357, EPI_ISL_5508358, EPI_ISL_5508359, EPI_ISL_5508360, EPI_ISL_5508361, EPI_ISL_5508362, EPI_ISL_5508363, EPI_ISL_5508364, EPI_ISL_5508365, EPI_ISL_5508366, EPI_ISL_5508367, EPI_ISL_5508368, EPI_ISL_5508369, EPI_ISL_5508370, EPI_ISL_5508371, EPI_ISL_5508372, EPI_ISL_5508373, EPI_ISL_5508374, EPI_ISL_5508375, EPI_ISL_5508376, EPI_ISL_5508377, EPI_ISL_5508378, EPI_ISL_5508379, EPI_ISL_5508380, EPI_ISL_5508381, EPI_ISL_5508382, EPI_ISL_5508383, EPI_ISL_5508384, EPI_ISL_5508385, EPI_ISL_5508386, EPI_ISL_5508387, EPI_ISL_5508388, EPI_ISL_5508389, EPI_ISL_5508390, EPI_ISL_5508391, EPI_ISL_5508392, EPI_ISL_5508393, EPI_ISL_5508394, EPI_ISL_5508395, EPI_ISL_5508396, EPI_ISL_5508397, EPI_ISL_5508398, EPI_ISL_5508399, EPI_ISL_5508400, EPI_ISL_5508401, EPI_ISL_5508402, EPI_ISL_5508403, EPI_ISL_5508404, EPI_ISL_5508405, EPI_ISL_5508406, EPI_ISL_5508407, EPI_ISL_5508408, EPI_ISL_5508409, EPI_ISL_5508410, EPI_ISL_5508411, EPI_ISL_5508412, EPI_ISL_5508413, EPI_ISL_5508414, EPI_ISL_5508415, EPI_ISL_5508416, EPI_ISL_5508417, EPI_ISL_5508418, EPI_ISL_5508419, EPI_ISL_5508420, EPI_ISL_5508421, EPI_ISL_5508422, EPI_ISL_5508423, EPI_ISL_5508424, EPI_ISL_5508425, EPI_ISL_5508426, EPI_ISL_5508427, EPI_ISL_5508428, EPI_ISL_5508429, EPI_ISL_5508430, EPI_ISL_5508431, EPI_ISL_5508432, EPI_ISL_5508433, EPI_ISL_5508434, EPI_ISL_5508437, EPI_ISL_5508438, EPI_ISL_5508439, EPI_ISL_5508440, EPI_ISL_5508441, EPI_ISL_5508442, EPI_ISL_5508443, EPI_ISL_5508444, EPI_ISL_5508445, EPI_ISL_5508446, EPI_ISL_5508447, EPI_ISL_5508448, EPI_ISL_5508449, EPI_ISL_5508450, EPI_ISL_5508451, EPI_ISL_5508452, EPI_ISL_5508453, EPI_ISL_5508454, EPI_ISL_5508455, EPI_ISL_5508456, EPI_ISL_5508457, EPI_ISL_5508458, EPI_ISL_5508459, EPI_ISL_5508460, EPI_ISL_5508461, EPI_ISL_5508462, EPI_ISL_5508463, EPI_ISL_5508464, EPI_ISL_5508465, EPI_ISL_5508466, EPI_ISL_5508467, EPI_ISL_5508468, EPI_ISL_5508469, EPI_ISL_5508470, EPI_ISL_5508471, EPI_ISL_5508472, EPI_ISL_5508473, EPI_ISL_5508474, EPI_ISL_5508475, EPI_ISL_5508476, EPI_ISL_5508477, EPI_ISL_5508478, EPI_ISL_5508479, EPI_ISL_5508480, EPI_ISL_5508481, EPI_ISL_5508482, EPI_ISL_5508483, EPI_ISL_5508484, EPI_ISL_5508485, EPI_ISL_5508486, EPI_ISL_5508487, EPI_ISL_5508488, EPI_ISL_5508489, EPI_ISL_5508490, EPI_ISL_5508491, EPI_ISL_5508492, EPI_ISL_5508493, EPI_ISL_5508494, EPI_ISL_5508495, EPI_ISL_5508496, EPI_ISL_5508497, EPI_ISL_5508498, EPI_ISL_5508499, EPI_ISL_5508500, EPI_ISL_5508501, EPI_ISL_5508502, EPI_ISL_5508503, EPI_ISL_5508504, EPI_ISL_5508505, EPI_ISL_5508506, EPI_ISL_5508507, EPI_ISL_5508508, EPI_ISL_5508509, EPI_ISL_5508510, EPI_ISL_5508511, EPI_ISL_5508512, EPI_ISL_5508513, EPI_ISL_5508514, EPI_ISL_5508515, EPI_ISL_5508516, EPI_ISL_5508517, EPI_ISL_5508518, EPI_ISL_5508519, EPI_ISL_5508520, EPI_ISL_5508521, EPI_ISL_5508522, EPI_ISL_5508526, EPI_ISL_5508527, EPI_ISL_5508528, EPI_ISL_5508529, EPI_ISL_5508530, EPI_ISL_5508531, EPI_ISL_5508532, EPI_ISL_5508533, EPI_ISL_5508534, EPI_ISL_5508535, EPI_ISL_5508536, EPI_ISL_5508537, EPI_ISL_5508538, EPI_ISL_5508539, EPI_ISL_5508540, EPI_ISL_5508541, EPI_ISL_5508542, EPI_ISL_5508543, EPI_ISL_5508544, EPI_ISL_5508545, EPI_ISL_5508546, EPI_ISL_5508547, EPI_ISL_5508548, EPI_ISL_5508549, EPI_ISL_5508550, EPI_ISL_5508551, EPI_ISL_5508552, EPI_ISL_5508553, EPI_ISL_5508554, EPI_ISL_5508555, EPI_ISL_5508556, EPI_ISL_5508557, EPI_ISL_5508558, EPI_ISL_5508559, EPI_ISL_5508560, EPI_ISL_5508561, EPI_ISL_5508562, EPI_ISL_5508563, EPI_ISL_5508564, EPI_ISL_5508565, EPI_ISL_5508566, EPI_ISL_5508567, EPI_ISL_5508568, EPI_ISL_5508569, EPI_ISL_5508570, EPI_ISL_5508571, EPI_ISL_5508572, EPI_ISL_5508573, EPI_ISL_5508574, EPI_ISL_5508575, EPI_ISL_5508576, EPI_ISL_5508577, EPI_ISL_5508578, EPI_ISL_5508579, EPI_ISL_5508580, EPI_ISL_5508581, EPI_ISL_5508582, EPI_ISL_5508583, EPI_ISL_5508584, EPI_ISL_5508585, EPI_ISL_5508586, EPI_ISL_5508587, EPI_ISL_5508588, EPI_ISL_5508589, EPI_ISL_5508590, EPI_ISL_5508591, EPI_ISL_5508592, EPI_ISL_5508593, EPI_ISL_5508594, EPI_ISL_5508595, EPI_ISL_5508596, EPI_ISL_5508600, EPI_ISL_5508601, EPI_ISL_5508602, EPI_ISL_5508603, EPI_ISL_5508604, EPI_ISL_5508605, EPI_ISL_5508606, EPI_ISL_5508607, EPI_ISL_5508608, EPI_ISL_5508609, EPI_ISL_5508610, EPI_ISL_5508611, EPI_ISL_5508612, EPI_ISL_5508613, EPI_ISL_5508614, EPI_ISL_5508615, EPI_ISL_5508616, EPI_ISL_5508617, EPI_ISL_5508618, EPI_ISL_5508619, EPI_ISL_5508620, EPI_ISL_5508621, EPI_ISL_5508622, EPI_ISL_5508623, EPI_ISL_5508624, EPI_ISL_5508625, EPI_ISL_5508626, EPI_ISL_5508627, EPI_ISL_5508628, EPI_ISL_5508629, EPI_ISL_5508630, EPI_ISL_5508631, EPI_ISL_5508632, EPI_ISL_5508633, EPI_ISL_5508634, EPI_ISL_5508635, EPI_ISL_5508636, EPI_ISL_5508637, EPI_ISL_5508638, EPI_ISL_5508639, EPI_ISL_5508640, EPI_ISL_5508641, EPI_ISL_5508642, EPI_ISL_5508643, EPI_ISL_5508644, EPI_ISL_5508645, EPI_ISL_5508646, EPI_ISL_5508647, EPI_ISL_5508648, EPI_ISL_5508649, EPI_ISL_5508650, EPI_ISL_5508651, EPI_ISL_5508652, EPI_ISL_5508653, EPI_ISL_5508654, EPI_ISL_5508655, EPI_ISL_5508656, EPI_ISL_5508657, EPI_ISL_5508658, EPI_ISL_5508659, EPI_ISL_5508660, EPI_ISL_5508661, EPI_ISL_5508662, EPI_ISL_5508663, EPI_ISL_5508664, EPI_ISL_5508666, EPI_ISL_5508667, EPI_ISL_5508668, EPI_ISL_5508669, EPI_ISL_5508670, EPI_ISL_5508671, EPI_ISL_5508672, EPI_ISL_5508673, EPI_ISL_5508674, EPI_ISL_5508675, EPI_ISL_5508676, EPI_ISL_5508677, EPI_ISL_5508678, EPI_ISL_5508679, EPI_ISL_5508680, EPI_ISL_5508681, EPI_ISL_5508683, EPI_ISL_5508684, EPI_ISL_5508685, EPI_ISL_5508686, EPI_ISL_5508687, EPI_ISL_5508688, EPI_ISL_5508689, EPI_ISL_5508690, EPI_ISL_5508691, EPI_ISL_5508692, EPI_ISL_5508693, EPI_ISL_5508694, EPI_ISL_5508695, EPI_ISL_5508696, EPI_ISL_5508697, EPI_ISL_5508698, EPI_ISL_5508699, EPI_ISL_5508700, EPI_ISL_5508701, EPI_ISL_5508702, EPI_ISL_5508704, EPI_ISL_5508705, EPI_ISL_5508706, EPI_ISL_5508707, EPI_ISL_5508708, EPI_ISL_5508709, EPI_ISL_5508710, EPI_ISL_5508711, EPI_ISL_5508712, EPI_ISL_5508713, EPI_ISL_5508714, EPI_ISL_5508715, EPI_ISL_5508716, EPI_ISL_5508717, EPI_ISL_5508718, EPI_ISL_5508719, EPI_ISL_5508721, EPI_ISL_5508722, EPI_ISL_5508723, EPI_ISL_5508724, EPI_ISL_5508725, EPI_ISL_5508726, EPI_ISL_5508727, EPI_ISL_5508728, EPI_ISL_5508731, EPI_ISL_5508732, EPI_ISL_5508733, EPI_ISL_5508734, EPI_ISL_5508735, EPI_ISL_5508736, EPI_ISL_5508737, EPI_ISL_5508739, EPI_ISL_5508740, EPI_ISL_5508741, EPI_ISL_5508742, EPI_ISL_5508743, EPI_ISL_5508744, EPI_ISL_5508746, EPI_ISL_5508747, EPI_ISL_5508748, EPI_ISL_5508749, EPI_ISL_5508750, EPI_ISL_5508752, EPI_ISL_5508753, EPI_ISL_5508754, EPI_ISL_5508755, EPI_ISL_5508756, EPI_ISL_5508757, EPI_ISL_5508758, EPI_ISL_5508759, EPI_ISL_5508760, EPI_ISL_5508761, EPI_ISL_5508762, EPI_ISL_5508763, EPI_ISL_5508764, EPI_ISL_5508765, EPI_ISL_5508766, EPI_ISL_5508767, EPI_ISL_5508768, EPI_ISL_5508769, EPI_ISL_5508770, EPI_ISL_5508771, EPI_ISL_5508772, EPI_ISL_5508773, EPI_ISL_5508774, EPI_ISL_5508775, EPI_ISL_5508776, EPI_ISL_5508778, EPI_ISL_5508779, EPI_ISL_5508781, EPI_ISL_5508782, EPI_ISL_5508783, EPI_ISL_5508784, EPI_ISL_5508785, EPI_ISL_5508787, EPI_ISL_5508788, EPI_ISL_5508789, EPI_ISL_5508791, EPI_ISL_5508792, EPI_ISL_5508793, EPI_ISL_5508794, EPI_ISL_5508795, EPI_ISL_5508796, EPI_ISL_5508797, EPI_ISL_5508798, EPI_ISL_5508799, EPI_ISL_5508800, EPI_ISL_5508801, EPI_ISL_5508802, EPI_ISL_5508803, EPI_ISL_5508804, EPI_ISL_5508805, EPI_ISL_5508806, EPI_ISL_5508807, EPI_ISL_5508811, EPI_ISL_5508812, EPI_ISL_5508814, EPI_ISL_5508815, EPI_ISL_5508817, EPI_ISL_5508818, EPI_ISL_5508819, EPI_ISL_5508820, EPI_ISL_5508821, EPI_ISL_5508823, EPI_ISL_5508824, EPI_ISL_5508825, EPI_ISL_5508826, EPI_ISL_5508827, EPI_ISL_5508828, EPI_ISL_5508829, EPI_ISL_5508830, EPI_ISL_5508833, EPI_ISL_5508834, EPI_ISL_5508835, EPI_ISL_5508836, EPI_ISL_5508837, EPI_ISL_5508839, EPI_ISL_5508840, EPI_ISL_5508841, EPI_ISL_5508844, EPI_ISL_5508847, EPI_ISL_5508848, EPI_ISL_5508849, EPI_ISL_5508850, EPI_ISL_5508851, EPI_ISL_5508853, EPI_ISL_5508854, EPI_ISL_5508855, EPI_ISL_5508856, EPI_ISL_5508860, EPI_ISL_5508861, EPI_ISL_5508862, EPI_ISL_5508863, EPI_ISL_5508864, EPI_ISL_5508865, EPI_ISL_5508866, EPI_ISL_5508867, EPI_ISL_5508869, EPI_ISL_5508870, EPI_ISL_5508871, EPI_ISL_5508875, EPI_ISL_5508876, EPI_ISL_5508877, EPI_ISL_5508880, EPI_ISL_5508881, EPI_ISL_5508882, EPI_ISL_5509266, EPI_ISL_5509267, EPI_ISL_5509268, EPI_ISL_5509269, EPI_ISL_5509270, EPI_ISL_5509271, EPI_ISL_5509272, EPI_ISL_5509273, EPI_ISL_5509274, EPI_ISL_5509281, EPI_ISL_5509287, EPI_ISL_5509289, EPI_ISL_5509293, EPI_ISL_5509300, EPI_ISL_5509305, EPI_ISL_5509314, EPI_ISL_5509320, EPI_ISL_5509326, EPI_ISL_5509336, EPI_ISL_5509340, EPI_ISL_5509346, EPI_ISL_5509348, EPI_ISL_5509356, EPI_ISL_5509359, EPI_ISL_5509369, EPI_ISL_5509376, EPI_ISL_5509384, EPI_ISL_5509390, EPI_ISL_5509394, EPI_ISL_5509399, EPI_ISL_5509408, EPI_ISL_5509419, EPI_ISL_5509429, EPI_ISL_5509440, EPI_ISL_5509450, EPI_ISL_5509459, EPI_ISL_5509469, EPI_ISL_5509483, EPI_ISL_5509484, EPI_ISL_5509492, EPI_ISL_5509500, EPI_ISL_5509506, EPI_ISL_5509507, EPI_ISL_5509512, EPI_ISL_5509522, EPI_ISL_5509525, EPI_ISL_5509530, EPI_ISL_5509539, EPI_ISL_5509544, EPI_ISL_5509545, EPI_ISL_5509550, EPI_ISL_5509554, EPI_ISL_5509564, EPI_ISL_5509571, EPI_ISL_5509572, EPI_ISL_5509573, EPI_ISL_5509574, EPI_ISL_5509575, EPI_ISL_5509576, EPI_ISL_5509577, EPI_ISL_5509578, EPI_ISL_5509579, EPI_ISL_5509580, EPI_ISL_5509581, EPI_ISL_5509582, EPI_ISL_5509583, EPI_ISL_5509584, EPI_ISL_5509585, EPI_ISL_5509586, EPI_ISL_5509587, EPI_ISL_5509588, EPI_ISL_5509589, EPI_ISL_5509593, EPI_ISL_5509594, EPI_ISL_5509595, EPI_ISL_5509596, EPI_ISL_5509597, EPI_ISL_5509598, EPI_ISL_5509599, EPI_ISL_5509600, EPI_ISL_5509601, EPI_ISL_5509602, EPI_ISL_5509603, EPI_ISL_5509604, EPI_ISL_5509605, EPI_ISL_5509606, EPI_ISL_5509607, EPI_ISL_5509608, EPI_ISL_5509609, EPI_ISL_5509610, EPI_ISL_5509611, EPI_ISL_5509612, EPI_ISL_5509613, EPI_ISL_5509614, EPI_ISL_5509615, EPI_ISL_5509616, EPI_ISL_5509617, EPI_ISL_5509618, EPI_ISL_5509619, EPI_ISL_5509620, EPI_ISL_5509621, EPI_ISL_5509622, EPI_ISL_5509623, EPI_ISL_5509624, EPI_ISL_5509625, EPI_ISL_5509626, EPI_ISL_5509627, EPI_ISL_5509628, EPI_ISL_5509629, EPI_ISL_5509630, EPI_ISL_5509631, EPI_ISL_5509632, EPI_ISL_5509633, EPI_ISL_5509634, EPI_ISL_5509635, EPI_ISL_5509636, EPI_ISL_5509637, EPI_ISL_5509638, EPI_ISL_5509639, EPI_ISL_5509640, EPI_ISL_5509641, EPI_ISL_5509642, EPI_ISL_5509643, EPI_ISL_5509644, EPI_ISL_5509645, EPI_ISL_5509646, EPI_ISL_5509647, EPI_ISL_5509648, EPI_ISL_5509651, EPI_ISL_5509652, EPI_ISL_5509653, EPI_ISL_5509654, EPI_ISL_5509655, EPI_ISL_5509656, EPI_ISL_5509657, EPI_ISL_5509658, EPI_ISL_5509659, EPI_ISL_5509660, EPI_ISL_5509661, EPI_ISL_5509662, EPI_ISL_5509663, EPI_ISL_5509664, EPI_ISL_5509665, EPI_ISL_5509666, EPI_ISL_5509667, EPI_ISL_5509668, EPI_ISL_5509669, EPI_ISL_5509670, EPI_ISL_5509671, EPI_ISL_5509672, EPI_ISL_5509673, EPI_ISL_5509674, EPI_ISL_5509675, EPI_ISL_5509676, EPI_ISL_5509677, EPI_ISL_5509678, EPI_ISL_5509679, EPI_ISL_5509680, EPI_ISL_5509681, EPI_ISL_5509682, EPI_ISL_5509683, EPI_ISL_5509684, EPI_ISL_5509685, EPI_ISL_5509686, EPI_ISL_5509687, EPI_ISL_5509688, EPI_ISL_5509689, EPI_ISL_5509690, EPI_ISL_5509691, EPI_ISL_5509692, EPI_ISL_5509693, EPI_ISL_5509694, EPI_ISL_5509695, EPI_ISL_5509696, EPI_ISL_5509697, EPI_ISL_5509698, EPI_ISL_5509699, EPI_ISL_5509700, EPI_ISL_5509701, EPI_ISL_5509702, EPI_ISL_5509703, EPI_ISL_5509704, EPI_ISL_5509705, EPI_ISL_5509706, EPI_ISL_5509707, EPI_ISL_5509708, EPI_ISL_5509710, EPI_ISL_5509711, EPI_ISL_5509712, EPI_ISL_5509713, EPI_ISL_5509714, EPI_ISL_5509715, EPI_ISL_5509716, EPI_ISL_5509717, EPI_ISL_5509718, EPI_ISL_5509719, EPI_ISL_5509720, EPI_ISL_5509721, EPI_ISL_5509722, EPI_ISL_5509723, EPI_ISL_5509724, EPI_ISL_5509725, EPI_ISL_5509726, EPI_ISL_5509727, EPI_ISL_5509728, EPI_ISL_5509729, EPI_ISL_5509730, EPI_ISL_5509731, EPI_ISL_5509738, EPI_ISL_5509752, EPI_ISL_5509765, EPI_ISL_5509776, EPI_ISL_5509785, EPI_ISL_5509788, EPI_ISL_5509793, EPI_ISL_5509800, EPI_ISL_5509805, EPI_ISL_5509810, EPI_ISL_5509813, EPI_ISL_5509823, EPI_ISL_5509827, EPI_ISL_5509835, EPI_ISL_5509841, EPI_ISL_5509850, EPI_ISL_5509856, EPI_ISL_5509858, EPI_ISL_5509859, EPI_ISL_5509860, EPI_ISL_5509861, EPI_ISL_5509862, EPI_ISL_5509863, EPI_ISL_5509864, EPI_ISL_5509865, EPI_ISL_5509866, EPI_ISL_5509867, EPI_ISL_5509868, EPI_ISL_5509869, EPI_ISL_5509870, EPI_ISL_5509871, EPI_ISL_5509872, EPI_ISL_5509873, EPI_ISL_5509874, EPI_ISL_5509875, EPI_ISL_5509876, EPI_ISL_5509877, EPI_ISL_5509878, EPI_ISL_5509879, EPI_ISL_5509880, EPI_ISL_5509881, EPI_ISL_5509882, EPI_ISL_5509883, EPI_ISL_5509884, EPI_ISL_5509885, EPI_ISL_5509886, EPI_ISL_5509887, EPI_ISL_5509888, EPI_ISL_5509889, EPI_ISL_5509890, EPI_ISL_5509891, EPI_ISL_5509892, EPI_ISL_5509893, EPI_ISL_5509894, EPI_ISL_5509895, EPI_ISL_5509896, EPI_ISL_5509897, EPI_ISL_5509898, EPI_ISL_5509899, EPI_ISL_5509900, EPI_ISL_5509901, EPI_ISL_5509902, EPI_ISL_5509903, EPI_ISL_5509904, EPI_ISL_5509905, EPI_ISL_5509906, EPI_ISL_5509907, EPI_ISL_5509908, EPI_ISL_5509909, EPI_ISL_5509911, EPI_ISL_5509912, EPI_ISL_5509913, EPI_ISL_5509914, EPI_ISL_5509915, EPI_ISL_5509916, EPI_ISL_5509917, EPI_ISL_5509918, EPI_ISL_5509919, EPI_ISL_5509920, EPI_ISL_5509921, EPI_ISL_5509922, EPI_ISL_5509923, EPI_ISL_5509924, EPI_ISL_5509925, EPI_ISL_5509926, EPI_ISL_5509927, EPI_ISL_5509928, EPI_ISL_5509929, EPI_ISL_5509930, EPI_ISL_5509931, EPI_ISL_5509932, EPI_ISL_5509933, EPI_ISL_5509934, EPI_ISL_5509935, EPI_ISL_5509936, EPI_ISL_5509937, EPI_ISL_5509938, EPI_ISL_5509939, EPI_ISL_5509940, EPI_ISL_5509941, EPI_ISL_5509942, EPI_ISL_5509943, EPI_ISL_5509944, EPI_ISL_5509945, EPI_ISL_5509946, EPI_ISL_5509947, EPI_ISL_5509948, EPI_ISL_5509949, EPI_ISL_5509950, EPI_ISL_5509951, EPI_ISL_5509952, EPI_ISL_5509953, EPI_ISL_5509954, EPI_ISL_5509955, EPI_ISL_5509956, EPI_ISL_5509957, EPI_ISL_5509958, EPI_ISL_5509959, EPI_ISL_5509960, EPI_ISL_5509961, EPI_ISL_5509962, EPI_ISL_5509963, EPI_ISL_5509964, EPI_ISL_5509965, EPI_ISL_5509966, EPI_ISL_5509967, EPI_ISL_5509968, EPI_ISL_5509969, EPI_ISL_5509970, EPI_ISL_5509971, EPI_ISL_5509972, EPI_ISL_5509973, EPI_ISL_5509974, EPI_ISL_5509975, EPI_ISL_5509976, EPI_ISL_5509977, EPI_ISL_5509978, EPI_ISL_5509979, EPI_ISL_5509980, EPI_ISL_5509981, EPI_ISL_5509982, EPI_ISL_5509983, EPI_ISL_5509984, EPI_ISL_5509985, EPI_ISL_5509986, EPI_ISL_5509987, EPI_ISL_5509988, EPI_ISL_5509989, EPI_ISL_5509990, EPI_ISL_5509991, EPI_ISL_5509992, EPI_ISL_5509993, EPI_ISL_5509994, EPI_ISL_5509995, EPI_ISL_5509996, EPI_ISL_5509997, EPI_ISL_5509998, EPI_ISL_5509999, EPI_ISL_5510000, EPI_ISL_5510001, EPI_ISL_5510002, EPI_ISL_5510003, EPI_ISL_5510004, EPI_ISL_5510005, EPI_ISL_5510006, EPI_ISL_5510007, EPI_ISL_5510008, EPI_ISL_5510009, EPI_ISL_5510010, EPI_ISL_5510011, EPI_ISL_5510012, EPI_ISL_5510013, EPI_ISL_5510014, EPI_ISL_5510015, EPI_ISL_5510016, EPI_ISL_5510017, EPI_ISL_5510018, EPI_ISL_5510019, EPI_ISL_5510020, EPI_ISL_5510021, EPI_ISL_5510022, EPI_ISL_5510023, EPI_ISL_5510026, EPI_ISL_5510027, EPI_ISL_5510028, EPI_ISL_5510029, EPI_ISL_5510030, EPI_ISL_5510031, EPI_ISL_5510032, EPI_ISL_5510033, EPI_ISL_5510034, EPI_ISL_5510035, EPI_ISL_5510036, EPI_ISL_5510037, EPI_ISL_5510038, EPI_ISL_5510039, EPI_ISL_5510040, EPI_ISL_5510041, EPI_ISL_5510042, EPI_ISL_5510043, EPI_ISL_5510044, EPI_ISL_5510045, EPI_ISL_5510046, EPI_ISL_5510047, EPI_ISL_5510048, EPI_ISL_5510049, EPI_ISL_5510050, EPI_ISL_5510051, EPI_ISL_5510052, EPI_ISL_5510053, EPI_ISL_5510054, EPI_ISL_5510055, EPI_ISL_5510056, EPI_ISL_5510057, EPI_ISL_5510058, EPI_ISL_5510059, EPI_ISL_5510060, EPI_ISL_5510061, EPI_ISL_5510062, EPI_ISL_5510063, EPI_ISL_5510064, EPI_ISL_5510065, EPI_ISL_5510066, EPI_ISL_5510067, EPI_ISL_5510068, EPI_ISL_5510069, EPI_ISL_5510070, EPI_ISL_5510071, EPI_ISL_5510072, EPI_ISL_5510073, EPI_ISL_5510074, EPI_ISL_5510075, EPI_ISL_5510076, EPI_ISL_5510077, EPI_ISL_5510078, EPI_ISL_5510079, EPI_ISL_5510080, EPI_ISL_5510081, EPI_ISL_5510082, EPI_ISL_5510083, EPI_ISL_5510086, EPI_ISL_5510087, EPI_ISL_5510088, EPI_ISL_5510089, EPI_ISL_5510090, EPI_ISL_5510091, EPI_ISL_5510092, EPI_ISL_5510093, EPI_ISL_5510094, EPI_ISL_5510095, EPI_ISL_5510096, EPI_ISL_5510097, EPI_ISL_5510098, EPI_ISL_5510099, EPI_ISL_5510100, EPI_ISL_5510101, EPI_ISL_5510102, EPI_ISL_5510103, EPI_ISL_5510104, EPI_ISL_5510105, EPI_ISL_5510106, EPI_ISL_5510107, EPI_ISL_5510108, EPI_ISL_5510109, EPI_ISL_5510110, EPI_ISL_5510111, EPI_ISL_5510112, EPI_ISL_5510113, EPI_ISL_5510114, EPI_ISL_5510115, EPI_ISL_5510116, EPI_ISL_5510117, EPI_ISL_5510118, EPI_ISL_5510119, EPI_ISL_5510120, EPI_ISL_5510121, EPI_ISL_5510122, EPI_ISL_5510123, EPI_ISL_5510124, EPI_ISL_5510125, EPI_ISL_5510126, EPI_ISL_5510127, EPI_ISL_5510128, EPI_ISL_5510129, EPI_ISL_5510130, EPI_ISL_5510131, EPI_ISL_5510132, EPI_ISL_5510133, EPI_ISL_5510136, EPI_ISL_5510137, EPI_ISL_5510138, EPI_ISL_5510139, EPI_ISL_5510140, EPI_ISL_5510141, EPI_ISL_5510142, EPI_ISL_5510143, EPI_ISL_5510144, EPI_ISL_5510145, EPI_ISL_5510146, EPI_ISL_5510147, EPI_ISL_5510148, EPI_ISL_5510149, EPI_ISL_5510150, EPI_ISL_5510151, EPI_ISL_5510152, EPI_ISL_5510153, EPI_ISL_5510154, EPI_ISL_5510155, EPI_ISL_5510156, EPI_ISL_5510157, EPI_ISL_5510158, EPI_ISL_5510159, EPI_ISL_5510160, EPI_ISL_5510161, EPI_ISL_5510162, EPI_ISL_5510163, EPI_ISL_5510164, EPI_ISL_5510165, EPI_ISL_5510166, EPI_ISL_5510167, EPI_ISL_5510168, EPI_ISL_5510169, EPI_ISL_5510170, EPI_ISL_5510171, EPI_ISL_5510172, EPI_ISL_5510173, EPI_ISL_5510174, EPI_ISL_5510175, EPI_ISL_5510176, EPI_ISL_5510177, EPI_ISL_5510178, EPI_ISL_5510180, EPI_ISL_5510181, EPI_ISL_5510182, EPI_ISL_5510183, EPI_ISL_5510184, EPI_ISL_5510185, EPI_ISL_5510186, EPI_ISL_5510187, EPI_ISL_5510188, EPI_ISL_5510189, EPI_ISL_5510190, EPI_ISL_5510191, EPI_ISL_5510192, EPI_ISL_5510193, EPI_ISL_5510194, EPI_ISL_5510195, EPI_ISL_5510196, EPI_ISL_5510197, EPI_ISL_5510198, EPI_ISL_5510199, EPI_ISL_5510200, EPI_ISL_5510201, EPI_ISL_5510202, EPI_ISL_5510203, EPI_ISL_5510204, EPI_ISL_5510205, EPI_ISL_5510206, EPI_ISL_5510207, EPI_ISL_5510208, EPI_ISL_5510209, EPI_ISL_5510210, EPI_ISL_5510211, EPI_ISL_5510212, EPI_ISL_5510213, EPI_ISL_5510214, EPI_ISL_5510215, EPI_ISL_5510216, EPI_ISL_5510217, EPI_ISL_5510218, EPI_ISL_5510219, EPI_ISL_5510220, EPI_ISL_5510221, EPI_ISL_5510222, EPI_ISL_5510223, EPI_ISL_5510224, EPI_ISL_5510225, EPI_ISL_5510226, EPI_ISL_5510227, EPI_ISL_5510228, EPI_ISL_5510229, EPI_ISL_5510230, EPI_ISL_5510231, EPI_ISL_5510232, EPI_ISL_5510233, EPI_ISL_5510234, EPI_ISL_5510235, EPI_ISL_5510236, EPI_ISL_5510237, EPI_ISL_5510238, EPI_ISL_5510239, EPI_ISL_5510240, EPI_ISL_5510241, EPI_ISL_5510242, EPI_ISL_5510243, EPI_ISL_5510244, EPI_ISL_5510245, EPI_ISL_5510246, EPI_ISL_5510247, EPI_ISL_5510248, EPI_ISL_5510249, EPI_ISL_5510250, EPI_ISL_5510251, EPI_ISL_5510252, EPI_ISL_5510253, EPI_ISL_5510254, EPI_ISL_5510255, EPI_ISL_5510256, EPI_ISL_5510257, EPI_ISL_5510258, EPI_ISL_5510259, EPI_ISL_5510260, EPI_ISL_5510261, EPI_ISL_5510262, EPI_ISL_5510263, EPI_ISL_5510267, EPI_ISL_5510268, EPI_ISL_5510269, EPI_ISL_5510270, EPI_ISL_5510271, EPI_ISL_5510272, EPI_ISL_5512665, EPI_ISL_5512666, EPI_ISL_5512681, EPI_ISL_5512687, EPI_ISL_5512695, EPI_ISL_5512701, EPI_ISL_5512707, EPI_ISL_5512708, EPI_ISL_5512715, EPI_ISL_5512722, EPI_ISL_5512729, EPI_ISL_5512735, EPI_ISL_5512742, EPI_ISL_5512749, EPI_ISL_5512754, EPI_ISL_5512764, EPI_ISL_5512765, EPI_ISL_5512771, EPI_ISL_5512776, EPI_ISL_5512789, EPI_ISL_5512790, EPI_ISL_5512800, EPI_ISL_5512808, EPI_ISL_5512812, EPI_ISL_5512821, EPI_ISL_5512825, EPI_ISL_5512833, EPI_ISL_5512834, EPI_ISL_5512843, EPI_ISL_5512848, EPI_ISL_5512855, EPI_ISL_5512858, EPI_ISL_5512864, EPI_ISL_5512865, EPI_ISL_5512874, EPI_ISL_5512883, EPI_ISL_5512888, EPI_ISL_5512889, EPI_ISL_5512895, EPI_ISL_5512902, EPI_ISL_5512909, EPI_ISL_5512910, EPI_ISL_5512917, EPI_ISL_5512924, EPI_ISL_5512925, EPI_ISL_5512928, EPI_ISL_5512934, EPI_ISL_5512935, EPI_ISL_5512942, EPI_ISL_5512951, EPI_ISL_5512954, EPI_ISL_5512955, EPI_ISL_5512960, EPI_ISL_5512965, EPI_ISL_5512970, EPI_ISL_5512971, EPI_ISL_5512978, EPI_ISL_5512979, EPI_ISL_5512982, EPI_ISL_5512984, EPI_ISL_5512993, EPI_ISL_5512994, EPI_ISL_5512998, EPI_ISL_5512999, EPI_ISL_5513004, EPI_ISL_5513007, EPI_ISL_5513014, EPI_ISL_5513016, EPI_ISL_5513019, EPI_ISL_5513024, EPI_ISL_5513029, EPI_ISL_5513034, EPI_ISL_5513039, EPI_ISL_5513040, EPI_ISL_5513048, EPI_ISL_5513062, EPI_ISL_5513063, EPI_ISL_5513069, EPI_ISL_5513073, EPI_ISL_5513079, EPI_ISL_5513080, EPI_ISL_5513083, EPI_ISL_5513090, EPI_ISL_5513100, EPI_ISL_5513106, EPI_ISL_5513107, EPI_ISL_5513115, EPI_ISL_5513120, EPI_ISL_5513130, EPI_ISL_5513131, EPI_ISL_5513138, EPI_ISL_5513139, EPI_ISL_5513145, EPI_ISL_5513150, EPI_ISL_5513155, EPI_ISL_5513161, EPI_ISL_5513166, EPI_ISL_5513182, EPI_ISL_5513187, EPI_ISL_5513188, EPI_ISL_5513200, EPI_ISL_5513201, EPI_ISL_5513204, EPI_ISL_5513212, EPI_ISL_5513217, EPI_ISL_5513224, EPI_ISL_5513231, EPI_ISL_5513237, EPI_ISL_5513238, EPI_ISL_5513243, EPI_ISL_5513246, EPI_ISL_5513252, EPI_ISL_5513260, EPI_ISL_5513266, EPI_ISL_5513269, EPI_ISL_5513270, EPI_ISL_5513285, EPI_ISL_5513292, EPI_ISL_5513298, EPI_ISL_5513299, EPI_ISL_5513300, EPI_ISL_5513301, EPI_ISL_5513302, EPI_ISL_5513307, EPI_ISL_5513321, EPI_ISL_5513326, EPI_ISL_5513327, EPI_ISL_5513334, EPI_ISL_5513342, EPI_ISL_5513343, EPI_ISL_5513351, EPI_ISL_5513360, EPI_ISL_5513370, EPI_ISL_5513372, EPI_ISL_5513375, EPI_ISL_5513376, EPI_ISL_5513382, EPI_ISL_5513388, EPI_ISL_5513397, EPI_ISL_5513398, EPI_ISL_5513404, EPI_ISL_5513410, EPI_ISL_5513411, EPI_ISL_5513419, EPI_ISL_5513426, EPI_ISL_5513429, EPI_ISL_5513430, EPI_ISL_5513440, EPI_ISL_5513448, EPI_ISL_5513449, EPI_ISL_5513456, EPI_ISL_5513464, EPI_ISL_5513470, EPI_ISL_5513473, EPI_ISL_5513487, EPI_ISL_5513497, EPI_ISL_5513502, EPI_ISL_5513511, EPI_ISL_5513529, EPI_ISL_5513538, EPI_ISL_5513539, EPI_ISL_5513545, EPI_ISL_5513551, EPI_ISL_5513559, EPI_ISL_5513565, EPI_ISL_5513573, EPI_ISL_5513574, EPI_ISL_5513580, EPI_ISL_5513587, EPI_ISL_5513589, EPI_ISL_5513591, EPI_ISL_5513592, EPI_ISL_5513598, EPI_ISL_5513601, EPI_ISL_5513602, EPI_ISL_5513610, EPI_ISL_5513616, EPI_ISL_5513618, EPI_ISL_5513619, EPI_ISL_5513627, EPI_ISL_5513635, EPI_ISL_5513640, EPI_ISL_5513641, EPI_ISL_5513646, EPI_ISL_5513651, EPI_ISL_5513661, EPI_ISL_5513666, EPI_ISL_5513674, EPI_ISL_5513680, EPI_ISL_5513684, EPI_ISL_5513687, EPI_ISL_5513697, EPI_ISL_5513703, EPI_ISL_5513704, EPI_ISL_5513711, EPI_ISL_5513714, EPI_ISL_5513718, EPI_ISL_5513723, EPI_ISL_5513724, EPI_ISL_5513733, EPI_ISL_5513737, EPI_ISL_5513743, EPI_ISL_5513751, EPI_ISL_5513759, EPI_ISL_5513769, EPI_ISL_5513770, EPI_ISL_5513773, EPI_ISL_5513781, EPI_ISL_5513791, EPI_ISL_5602431, EPI_ISL_5602432, EPI_ISL_5602433, EPI_ISL_5602434, EPI_ISL_5602435, EPI_ISL_5602436, EPI_ISL_5602437, EPI_ISL_5602438, EPI_ISL_5602439, EPI_ISL_5602440, EPI_ISL_5602441, EPI_ISL_5602442, EPI_ISL_5602443, EPI_ISL_5602444, EPI_ISL_5602445, EPI_ISL_5602446, EPI_ISL_5602447, EPI_ISL_5602448, EPI_ISL_5602449, EPI_ISL_5602450, EPI_ISL_5602451, EPI_ISL_5602452, EPI_ISL_5602453, EPI_ISL_5602454, EPI_ISL_5602455, EPI_ISL_5602456, EPI_ISL_5602457, EPI_ISL_5602458, EPI_ISL_5602459, EPI_ISL_5602460, EPI_ISL_5602461, EPI_ISL_5602462, EPI_ISL_5602463, EPI_ISL_5602464, EPI_ISL_5602465, EPI_ISL_5602466, EPI_ISL_5602467, EPI_ISL_5602468, EPI_ISL_5602469, EPI_ISL_5602470, EPI_ISL_5602471, EPI_ISL_5602472, EPI_ISL_5602473, EPI_ISL_5602477, EPI_ISL_5602478, EPI_ISL_5602479, EPI_ISL_5602480, EPI_ISL_5602481, EPI_ISL_5602482, EPI_ISL_5602483, EPI_ISL_5602484, EPI_ISL_5602485, EPI_ISL_5602486, EPI_ISL_5602487, EPI_ISL_5602488, EPI_ISL_5602489, EPI_ISL_5602490, EPI_ISL_5602491, EPI_ISL_5602492, EPI_ISL_5602493, EPI_ISL_5602494, EPI_ISL_5602495, EPI_ISL_5602496, EPI_ISL_5602497, EPI_ISL_5602498, EPI_ISL_5602499, EPI_ISL_5602500, EPI_ISL_5602501, EPI_ISL_5602502, EPI_ISL_5602503, EPI_ISL_5602504, EPI_ISL_5602505, EPI_ISL_5602506, EPI_ISL_5602507, EPI_ISL_5602508, EPI_ISL_5602509, EPI_ISL_5602510, EPI_ISL_5602511, EPI_ISL_5602512, EPI_ISL_5602513, EPI_ISL_5602514, EPI_ISL_5602515, EPI_ISL_5602516, EPI_ISL_5602517, EPI_ISL_5602518, EPI_ISL_5602519, EPI_ISL_5602520, EPI_ISL_5602521, EPI_ISL_5602523, EPI_ISL_5602524, EPI_ISL_5602525, EPI_ISL_5602526, EPI_ISL_5602527, EPI_ISL_5602528, EPI_ISL_5602529, EPI_ISL_5602530, EPI_ISL_5602531, EPI_ISL_5602532, EPI_ISL_5602533, EPI_ISL_5602534, EPI_ISL_5602535, EPI_ISL_5602536, EPI_ISL_5602537, EPI_ISL_5602538, EPI_ISL_5602539, EPI_ISL_5602540, EPI_ISL_5602541, EPI_ISL_5602542, EPI_ISL_5602543, EPI_ISL_5602544, EPI_ISL_5602545, EPI_ISL_5602546, EPI_ISL_5602547, EPI_ISL_5602548, EPI_ISL_5602549, EPI_ISL_5602550, EPI_ISL_5602551, EPI_ISL_5602552, EPI_ISL_5602553, EPI_ISL_5602554, EPI_ISL_5602555, EPI_ISL_5602556, EPI_ISL_5602557, EPI_ISL_5602560, EPI_ISL_5602561, EPI_ISL_5602562, EPI_ISL_5602563, EPI_ISL_5602564, EPI_ISL_5602565, EPI_ISL_5602566, EPI_ISL_5602567, EPI_ISL_5602568, EPI_ISL_5602569, EPI_ISL_5602570, EPI_ISL_5602571, EPI_ISL_5602572, EPI_ISL_5602573, EPI_ISL_5602574, EPI_ISL_5602575, EPI_ISL_5602576, EPI_ISL_5602577, EPI_ISL_5602578, EPI_ISL_5602579, EPI_ISL_5602580, EPI_ISL_5602581, EPI_ISL_5602582, EPI_ISL_5602583, EPI_ISL_5602584, EPI_ISL_5602585, EPI_ISL_5602586, EPI_ISL_5602587, EPI_ISL_5602588, EPI_ISL_5602589, EPI_ISL_5602590, EPI_ISL_5602591, EPI_ISL_5602592, EPI_ISL_5602593, EPI_ISL_5602597, EPI_ISL_5602598, EPI_ISL_5602599, EPI_ISL_5602601, EPI_ISL_5602602, EPI_ISL_5602603, EPI_ISL_5602604, EPI_ISL_5602605, EPI_ISL_5602606, EPI_ISL_5602607, EPI_ISL_5602608, EPI_ISL_5602609, EPI_ISL_5602613, EPI_ISL_5602647, EPI_ISL_5602648, EPI_ISL_5602649, EPI_ISL_5602650, EPI_ISL_5602651, EPI_ISL_5602653, EPI_ISL_5602654, EPI_ISL_5602655, EPI_ISL_5602656, EPI_ISL_5602657, EPI_ISL_5602658, EPI_ISL_5602659, EPI_ISL_5602660, EPI_ISL_5602661, EPI_ISL_5602662, EPI_ISL_5602663, EPI_ISL_5602665, EPI_ISL_5602666, EPI_ISL_5602667, EPI_ISL_5602668, EPI_ISL_5602669, EPI_ISL_5602670, EPI_ISL_5602671, EPI_ISL_5602672, EPI_ISL_5602673, EPI_ISL_5602674, EPI_ISL_5602675, EPI_ISL_5602676, EPI_ISL_5602677, EPI_ISL_5602678, EPI_ISL_5602679, EPI_ISL_5602680, EPI_ISL_5602681, EPI_ISL_5602682, EPI_ISL_5602683, EPI_ISL_5602684, EPI_ISL_5602688, EPI_ISL_5602689, EPI_ISL_5602690, EPI_ISL_5602691, EPI_ISL_5602692, EPI_ISL_5602693, EPI_ISL_5602694, EPI_ISL_5602695, EPI_ISL_5602696, EPI_ISL_5602697, EPI_ISL_5602698, EPI_ISL_5602699, EPI_ISL_5602700, EPI_ISL_5602701, EPI_ISL_5602702, EPI_ISL_5602703, EPI_ISL_5602704, EPI_ISL_5602705, EPI_ISL_5602706, EPI_ISL_5602707, EPI_ISL_5602708, EPI_ISL_5602709, EPI_ISL_5602710, EPI_ISL_5602711, EPI_ISL_5602712, EPI_ISL_5602713, EPI_ISL_5602714, EPI_ISL_5602715, EPI_ISL_5602716, EPI_ISL_5602717, EPI_ISL_5602718, EPI_ISL_5602719, EPI_ISL_5602720, EPI_ISL_5602721, EPI_ISL_5602722, EPI_ISL_5602723, EPI_ISL_5602724, EPI_ISL_5602725, EPI_ISL_5602726, EPI_ISL_5602727, EPI_ISL_5602728, EPI_ISL_5602729, EPI_ISL_5602730, EPI_ISL_5602731, EPI_ISL_5602732, EPI_ISL_5602733, EPI_ISL_5602734, EPI_ISL_5602735, EPI_ISL_5602736, EPI_ISL_5602737, EPI_ISL_5620224, EPI_ISL_5620225, EPI_ISL_5620226, EPI_ISL_5620227, EPI_ISL_5620228, EPI_ISL_5620229, EPI_ISL_5620230, EPI_ISL_5620231, EPI_ISL_5620232, EPI_ISL_5620233, EPI_ISL_5620234, EPI_ISL_5620235, EPI_ISL_5620236, EPI_ISL_5620237, EPI_ISL_5620238, EPI_ISL_5620239, EPI_ISL_5620240, EPI_ISL_5620241, EPI_ISL_5620242, EPI_ISL_5620243, EPI_ISL_5620244, EPI_ISL_5620245, EPI_ISL_5620246, EPI_ISL_5620247, EPI_ISL_5620248, EPI_ISL_5620249, EPI_ISL_5620250, EPI_ISL_5620251, EPI_ISL_5620252, EPI_ISL_5620253, EPI_ISL_5620254, EPI_ISL_5620255, EPI_ISL_5620256, EPI_ISL_5620257, EPI_ISL_5620258, EPI_ISL_5620259, EPI_ISL_5620260, EPI_ISL_5620261, EPI_ISL_5620262, EPI_ISL_5620263, EPI_ISL_5620264, EPI_ISL_5620265, EPI_ISL_5620266, EPI_ISL_5620267, EPI_ISL_5620268, EPI_ISL_5620269, EPI_ISL_5620270, EPI_ISL_5620271, EPI_ISL_5620272, EPI_ISL_5620273, EPI_ISL_5620274, EPI_ISL_5620275, EPI_ISL_5620276, EPI_ISL_5620277, EPI_ISL_5620278, EPI_ISL_5620279, EPI_ISL_5620280, EPI_ISL_5620281, EPI_ISL_5620282, EPI_ISL_5620283, EPI_ISL_5620284, EPI_ISL_5620285, EPI_ISL_5620286, EPI_ISL_5620287, EPI_ISL_5620288, EPI_ISL_5620289, EPI_ISL_5620290, EPI_ISL_5620291, EPI_ISL_5620292, EPI_ISL_5620293, EPI_ISL_5620294, EPI_ISL_5620295, EPI_ISL_5620296, EPI_ISL_5620297, EPI_ISL_5620298, EPI_ISL_5620299, EPI_ISL_5620300, EPI_ISL_5620301, EPI_ISL_5620302, EPI_ISL_5620303, EPI_ISL_5620304, EPI_ISL_5620305, EPI_ISL_5620306, EPI_ISL_5620307, EPI_ISL_5620308, EPI_ISL_5620310, EPI_ISL_5620311, EPI_ISL_5620312, EPI_ISL_5620313, EPI_ISL_5620314, EPI_ISL_5620315, EPI_ISL_5620316, EPI_ISL_5620317, EPI_ISL_5620318, EPI_ISL_5620319, EPI_ISL_5620320, EPI_ISL_5620321, EPI_ISL_5620322, EPI_ISL_5620323, EPI_ISL_5620324, EPI_ISL_5620325, EPI_ISL_5620326, EPI_ISL_5620327, EPI_ISL_5620328, EPI_ISL_5620329, EPI_ISL_5620330, EPI_ISL_5620331, EPI_ISL_5620332, EPI_ISL_5620333, EPI_ISL_5620334, EPI_ISL_5620335, EPI_ISL_5620336, EPI_ISL_5620337, EPI_ISL_5620338, EPI_ISL_5620339, EPI_ISL_5620340, EPI_ISL_5620341, EPI_ISL_5620342, EPI_ISL_5620343, EPI_ISL_5620344, EPI_ISL_5620345, EPI_ISL_5620346, EPI_ISL_5620347, EPI_ISL_5620348, EPI_ISL_5620349, EPI_ISL_5620350, EPI_ISL_5620351, EPI_ISL_5620352, EPI_ISL_5620353, EPI_ISL_5620354, EPI_ISL_5620356, EPI_ISL_5620357, EPI_ISL_5620358, EPI_ISL_5620359, EPI_ISL_5620360, EPI_ISL_5620361, EPI_ISL_5620362, EPI_ISL_5620363, EPI_ISL_5620365, EPI_ISL_5620366, EPI_ISL_5620367, EPI_ISL_5620368, EPI_ISL_5620369, EPI_ISL_5620370, EPI_ISL_5620371, EPI_ISL_5620372, EPI_ISL_5620373, EPI_ISL_5620374, EPI_ISL_5620375, EPI_ISL_5620376, EPI_ISL_5620377, EPI_ISL_5620378, EPI_ISL_5620379, EPI_ISL_5620380, EPI_ISL_5620381, EPI_ISL_5620382, EPI_ISL_5620383, EPI_ISL_5620384, EPI_ISL_5620385, EPI_ISL_5620386, EPI_ISL_5620387, EPI_ISL_5620388, | | | |

EPI_ISL_5620389, EPI_ISL_5620390, EPI_ISL_5620391, EPI_ISL_5620392, EPI_ISL_5620393, EPI_ISL_5620394, EPI_ISL_5620395, EPI_ISL_5620396, EPI_ISL_5620397, EPI_ISL_5620398, EPI_ISL_5620399, EPI_ISL_5620400, EPI_ISL_5620401, EPI_ISL_5620402, EPI_ISL_5620403, EPI_ISL_5620404, EPI_ISL_5620405, EPI_ISL_5620406, EPI_ISL_5620407, EPI_ISL_5620408, EPI_ISL_5620409, EPI_ISL_5620410,
EPI_ISL_5620411, EPI_ISL_5620412, EPI_ISL_5620413, EPI_ISL_5620414, EPI_ISL_5620415, EPI_ISL_5620417, EPI_ISL_5620418, EPI_ISL_5620419, EPI_ISL_5620420, EPI_ISL_5620421, EPI_ISL_5620422, EPI_ISL_5620423, EPI_ISL_5620424, EPI_ISL_5620425, EPI_ISL_5620426, EPI_ISL_5620427, EPI_ISL_5620428, EPI_ISL_5620429, EPI_ISL_5620430, EPI_ISL_5620431, EPI_ISL_5620433, EPI_ISL_5620434,
EPI_ISL_5620435, EPI_ISL_5620436, EPI_ISL_5620437, EPI_ISL_5620438, EPI_ISL_5620439, EPI_ISL_5620440, EPI_ISL_5620441, EPI_ISL_5620442, EPI_ISL_5620445, EPI_ISL_5620446, EPI_ISL_5620447, EPI_ISL_5620448, EPI_ISL_5620449, EPI_ISL_5620450, EPI_ISL_5620451, EPI_ISL_5620452, EPI_ISL_5620453, EPI_ISL_5620454, EPI_ISL_5620455, EPI_ISL_5620456, EPI_ISL_5620457, EPI_ISL_5620458,
EPI_ISL_5620459, EPI_ISL_5620460, EPI_ISL_5620461, EPI_ISL_5620462, EPI_ISL_5620463, EPI_ISL_5620464, EPI_ISL_5620465, EPI_ISL_5620466, EPI_ISL_5620467, EPI_ISL_5620468, EPI_ISL_5620469, EPI_ISL_5620470, EPI_ISL_5620471, EPI_ISL_5620472, EPI_ISL_5620475, EPI_ISL_5620476, EPI_ISL_5620477, EPI_ISL_5620478, EPI_ISL_5620479, EPI_ISL_5620480, EPI_ISL_5620481, EPI_ISL_5620482,
EPI_ISL_5620483, EPI_ISL_5620484, EPI_ISL_5620485, EPI_ISL_5620486, EPI_ISL_5620487, EPI_ISL_5620488, EPI_ISL_5620489, EPI_ISL_5620490, EPI_ISL_5620491, EPI_ISL_5620492, EPI_ISL_5620493, EPI_ISL_5620494, EPI_ISL_5620495, EPI_ISL_5620496, EPI_ISL_5620497, EPI_ISL_5620498, EPI_ISL_5620499, EPI_ISL_5620500, EPI_ISL_5620501, EPI_ISL_5620502, EPI_ISL_5620503, EPI_ISL_5620504,
EPI_ISL_5620505, EPI_ISL_5620506, EPI_ISL_5620507, EPI_ISL_5620508, EPI_ISL_5620509, EPI_ISL_5620510, EPI_ISL_5620511, EPI_ISL_5620512, EPI_ISL_5620513, EPI_ISL_5620514, EPI_ISL_5620515, EPI_ISL_5620516, EPI_ISL_5620517, EPI_ISL_5620519, EPI_ISL_5620520, EPI_ISL_5620521, EPI_ISL_5620522, EPI_ISL_5620523, EPI_ISL_5620524, EPI_ISL_5620525, EPI_ISL_5620526, EPI_ISL_5620527,
EPI_ISL_5620529, EPI_ISL_5620530, EPI_ISL_5620531, EPI_ISL_5620532, EPI_ISL_5620533, EPI_ISL_5620534, EPI_ISL_5620535, EPI_ISL_5620609, EPI_ISL_5620611, EPI_ISL_5620612, EPI_ISL_5620613, EPI_ISL_5620616, EPI_ISL_5620617, EPI_ISL_5620618, EPI_ISL_5620619, EPI_ISL_5620621, EPI_ISL_5620623, EPI_ISL_5620624, EPI_ISL_5620626, EPI_ISL_5620627, EPI_ISL_5620628, EPI_ISL_5620629,
EPI_ISL_5620630, EPI_ISL_5620631, EPI_ISL_5620632, EPI_ISL_5620633, EPI_ISL_5620634, EPI_ISL_5620635, EPI_ISL_5620636, EPI_ISL_5620637, EPI_ISL_5620638, EPI_ISL_5620639, EPI_ISL_5620640, EPI_ISL_5620642, EPI_ISL_5620643, EPI_ISL_5620644, EPI_ISL_5620645, EPI_ISL_5620646, EPI_ISL_5620647, EPI_ISL_5620648, EPI_ISL_5620649, EPI_ISL_5620650, EPI_ISL_5620651, EPI_ISL_5620652,
EPI_ISL_5620653, EPI_ISL_5620654, EPI_ISL_5620655, EPI_ISL_5620656, EPI_ISL_5620657, EPI_ISL_5620658, EPI_ISL_5620659, EPI_ISL_5620660, EPI_ISL_5620661, EPI_ISL_5620666, EPI_ISL_5620667, EPI_ISL_5620668, EPI_ISL_5620669, EPI_ISL_5620670, EPI_ISL_5620671, EPI_ISL_5620673, EPI_ISL_5620674, EPI_ISL_5620675, EPI_ISL_5620676, EPI_ISL_5620677, EPI_ISL_5620678, EPI_ISL_5620679,
EPI_ISL_5620680, EPI_ISL_5620681, EPI_ISL_5620682, EPI_ISL_5620683, EPI_ISL_5620684, EPI_ISL_5620685, EPI_ISL_5620686, EPI_ISL_5620687, EPI_ISL_5620688, EPI_ISL_5620689, EPI_ISL_5620690, EPI_ISL_5620691, EPI_ISL_5620692, EPI_ISL_5620693, EPI_ISL_5620694, EPI_ISL_5620695, EPI_ISL_5620696, EPI_ISL_5620697, EPI_ISL_5620698, EPI_ISL_5620699, EPI_ISL_5620700, EPI_ISL_5620701,
EPI_ISL_5620702, EPI_ISL_5620703, EPI_ISL_5620704, EPI_ISL_5620705, EPI_ISL_5620706, EPI_ISL_5620707, EPI_ISL_5620708, EPI_ISL_5620709, EPI_ISL_5620710, EPI_ISL_5620711, EPI_ISL_5620712, EPI_ISL_5620713, EPI_ISL_5620714, EPI_ISL_5620715, EPI_ISL_5620716, EPI_ISL_5620717, EPI_ISL_5620718, EPI_ISL_5620719, EPI_ISL_5620720, EPI_ISL_5620721, EPI_ISL_5620722, EPI_ISL_5620723,
EPI_ISL_5620724, EPI_ISL_5620725, EPI_ISL_5620726, EPI_ISL_5620727, EPI_ISL_5620728, EPI_ISL_5620729, EPI_ISL_5620730, EPI_ISL_5620731, EPI_ISL_5620732, EPI_ISL_5620733, EPI_ISL_5620734, EPI_ISL_5620738, EPI_ISL_5620739, EPI_ISL_5620740, EPI_ISL_5620741, EPI_ISL_5620742, EPI_ISL_5620743, EPI_ISL_5620744, EPI_ISL_5620745, EPI_ISL_5620746, EPI_ISL_5620747, EPI_ISL_5620748,
EPI_ISL_5620749, EPI_ISL_5620750, EPI_ISL_5620751, EPI_ISL_5620752, EPI_ISL_5620753, EPI_ISL_5620754, EPI_ISL_5620755, EPI_ISL_5620756, EPI_ISL_5620757, EPI_ISL_5620758, EPI_ISL_5620759, EPI_ISL_5620760, EPI_ISL_5620761, EPI_ISL_5620762, EPI_ISL_5620763, EPI_ISL_5620764, EPI_ISL_5620765, EPI_ISL_5620766, EPI_ISL_5620767, EPI_ISL_5620768, EPI_ISL_5620769, EPI_ISL_5620770,
EPI_ISL_5620771, EPI_ISL_5620772, EPI_ISL_5620773, EPI_ISL_5620774, EPI_ISL_5620775, EPI_ISL_5620776, EPI_ISL_5620777, EPI_ISL_5620778, EPI_ISL_5620779, EPI_ISL_5620780, EPI_ISL_5620781, EPI_ISL_5620782, EPI_ISL_5620783, EPI_ISL_5620784, EPI_ISL_5620785, EPI_ISL_5620786, EPI_ISL_5620787, EPI_ISL_5620788, EPI_ISL_5620789, EPI_ISL_5620790, EPI_ISL_5620791, EPI_ISL_5620792,
EPI_ISL_5620793, EPI_ISL_5620794, EPI_ISL_5620795, EPI_ISL_5620796, EPI_ISL_5620797, EPI_ISL_5620798, EPI_ISL_5620799, EPI_ISL_5620800, EPI_ISL_5620801, EPI_ISL_5620802, EPI_ISL_5620803, EPI_ISL_5620804, EPI_ISL_5620805, EPI_ISL_5620806, EPI_ISL_5620807, EPI_ISL_5620808, EPI_ISL_5620809, EPI_ISL_5620810, EPI_ISL_5620814, EPI_ISL_5620815, EPI_ISL_5620816, EPI_ISL_5620817,
EPI_ISL_5620818, EPI_ISL_5620819, EPI_ISL_5620820, EPI_ISL_5620821, EPI_ISL_5620822, EPI_ISL_5620823, EPI_ISL_5620824, EPI_ISL_5620825, EPI_ISL_5620826, EPI_ISL_5620827, EPI_ISL_5620828, EPI_ISL_5620829, EPI_ISL_5620830, EPI_ISL_5620831, EPI_ISL_5620832, EPI_ISL_5620833, EPI_ISL_5620834, EPI_ISL_5620835, EPI_ISL_5620836, EPI_ISL_5620837, EPI_ISL_5620838, EPI_ISL_5620839,
EPI_ISL_5620840, EPI_ISL_5620841, EPI_ISL_5620842, EPI_ISL_5620843, EPI_ISL_5620844, EPI_ISL_5620845, EPI_ISL_5620846, EPI_ISL_5620847, EPI_ISL_5620848, EPI_ISL_5620849, EPI_ISL_5620850, EPI_ISL_5620851, EPI_ISL_5620852, EPI_ISL_5620853, EPI_ISL_5620854, EPI_ISL_5620855, EPI_ISL_5620858, EPI_ISL_5620859, EPI_ISL_5620860, EPI_ISL_5620861, EPI_ISL_5620862, EPI_ISL_5620863,
EPI_ISL_5620864, EPI_ISL_5620865, EPI_ISL_5620866, EPI_ISL_5620867, EPI_ISL_5620868, EPI_ISL_5620869, EPI_ISL_5620870, EPI_ISL_5620871, EPI_ISL_5620872, EPI_ISL_5620873, EPI_ISL_5620874, EPI_ISL_5620875, EPI_ISL_5620876, EPI_ISL_5620877, EPI_ISL_5620878, EPI_ISL_5620882, EPI_ISL_5620883, EPI_ISL_5620884, EPI_ISL_5620885, EPI_ISL_5620886, EPI_ISL_5620887, EPI_ISL_5620888,
EPI_ISL_5620889, EPI_ISL_5620890, EPI_ISL_5620891, EPI_ISL_5620892, EPI_ISL_5620893, EPI_ISL_5620894, EPI_ISL_5620895, EPI_ISL_5620896, EPI_ISL_5620897, EPI_ISL_5620898, EPI_ISL_5620899, EPI_ISL_5620900, EPI_ISL_5620901, EPI_ISL_5620902, EPI_ISL_5620903, EPI_ISL_5620904, EPI_ISL_5620905, EPI_ISL_5620906, EPI_ISL_5620907, EPI_ISL_5620908, EPI_ISL_5620909, EPI_ISL_5620910,
EPI_ISL_5620911, EPI_ISL_5620912, EPI_ISL_5620913, EPI_ISL_5620914, EPI_ISL_5620915, EPI_ISL_5620916, EPI_ISL_5620917, EPI_ISL_5620918, EPI_ISL_5620919, EPI_ISL_5620920, EPI_ISL_5620921, EPI_ISL_5620922, EPI_ISL_5620923, EPI_ISL_5620924, EPI_ISL_5620925, EPI_ISL_5620926, EPI_ISL_5620927, EPI_ISL_5620928, EPI_ISL_5620929, EPI_ISL_5620930, EPI_ISL_5620931, EPI_ISL_5620932,
EPI_ISL_5620933, EPI_ISL_5620934, EPI_ISL_5620935, EPI_ISL_5620936, EPI_ISL_5620937, EPI_ISL_5620938, EPI_ISL_5620939, EPI_ISL_5620940, EPI_ISL_5620941, EPI_ISL_5620942, EPI_ISL_5620943, EPI_ISL_5620944, EPI_ISL_5620945, EPI_ISL_5620946, EPI_ISL_5620947, EPI_ISL_5620948, EPI_ISL_5620953, EPI_ISL_5620954, EPI_ISL_5620955, EPI_ISL_5620956, EPI_ISL_5620957, EPI_ISL_5620958,
EPI_ISL_5620959, EPI_ISL_5620960, EPI_ISL_5620961, EPI_ISL_5620962, EPI_ISL_5620963, EPI_ISL_5620964, EPI_ISL_5620965, EPI_ISL_5620966, EPI_ISL_5620967, EPI_ISL_5620968, EPI_ISL_5620969, EPI_ISL_5620970, EPI_ISL_5620971, EPI_ISL_5620972, EPI_ISL_5620973, EPI_ISL_5620974, EPI_ISL_5620975, EPI_ISL_5620976, EPI_ISL_5620977, EPI_ISL_5620978, EPI_ISL_5620979, EPI_ISL_5620980,
EPI_ISL_5620981, EPI_ISL_5620982, EPI_ISL_5620983, EPI_ISL_5620984, EPI_ISL_5620985, EPI_ISL_5620986, EPI_ISL_5620987, EPI_ISL_5620988, EPI_ISL_5620989, EPI_ISL_5620990, EPI_ISL_5620991, EPI_ISL_5620992, EPI_ISL_5620993, EPI_ISL_5620994, EPI_ISL_5620995, EPI_ISL_5620996, EPI_ISL_5620997, EPI_ISL_5620998, EPI_ISL_5620999, EPI_ISL_5621000, EPI_ISL_5621001, EPI_ISL_5621002,
EPI_ISL_5621003, EPI_ISL_5621004, EPI_ISL_5621005, EPI_ISL_5621006, EPI_ISL_5621007, EPI_ISL_5621008, EPI_ISL_5621009, EPI_ISL_5621010, EPI_ISL_5621011, EPI_ISL_5621012, EPI_ISL_5621013, EPI_ISL_5621014, EPI_ISL_5621015, EPI_ISL_5621016, EPI_ISL_5621017, EPI_ISL_5621018, EPI_ISL_5621019, EPI_ISL_5621020, EPI_ISL_5621021, EPI_ISL_5621023, EPI_ISL_5621024, EPI_ISL_5621025,
EPI_ISL_5621026, EPI_ISL_5621027, EPI_ISL_5621028, EPI_ISL_5621029, EPI_ISL_5621030, EPI_ISL_5621031, EPI_ISL_5621032, EPI_ISL_5621033, EPI_ISL_5621034, EPI_ISL_5621035, EPI_ISL_5621036, EPI_ISL_5621037, EPI_ISL_5621038, EPI_ISL_5621039, EPI_ISL_5621040, EPI_ISL_5621041, EPI_ISL_5621042, EPI_ISL_5621043, EPI_ISL_5621044, EPI_ISL_5621045, EPI_ISL_5621046, EPI_ISL_5621047,
EPI_ISL_5621048, EPI_ISL_5621049, EPI_ISL_5621050, EPI_ISL_5621051, EPI_ISL_5621052, EPI_ISL_5621053, EPI_ISL_5621054, EPI_ISL_5621055, EPI_ISL_5621056, EPI_ISL_5621057, EPI_ISL_5621058, EPI_ISL_5621059, EPI_ISL_5621060, EPI_ISL_5621061, EPI_ISL_5621062, EPI_ISL_5621063, EPI_ISL_5621064, EPI_ISL_5621065, EPI_ISL_5621066, EPI_ISL_5621067, EPI_ISL_5621068, EPI_ISL_5621069,
EPI_ISL_5621070, EPI_ISL_5621071, EPI_ISL_5621072, EPI_ISL_5621073, EPI_ISL_5621074, EPI_ISL_5621075, EPI_ISL_5621076, EPI_ISL_5621077, EPI_ISL_5621078, EPI_ISL_5621079, EPI_ISL_5621080, EPI_ISL_5621081, EPI_ISL_5621082, EPI_ISL_5621083, EPI_ISL_5621084, EPI_ISL_5621085, EPI_ISL_5621086, EPI_ISL_5621087, EPI_ISL_5621088, EPI_ISL_5621089, EPI_ISL_5621090, EPI_ISL_5621091,
EPI_ISL_5621096, EPI_ISL_5621097, EPI_ISL_5621098, EPI_ISL_5621099, EPI_ISL_5621100, EPI_ISL_5621101, EPI_ISL_5621102, EPI_ISL_5621103, EPI_ISL_5621104, EPI_ISL_5621105, EPI_ISL_5621106, EPI_ISL_5621107, EPI_ISL_5621108, EPI_ISL_5621109, EPI_ISL_5621110, EPI_ISL_5621111, EPI_ISL_5621112, EPI_ISL_5621113, EPI_ISL_5621114, EPI_ISL_5621115, EPI_ISL_5621116, EPI_ISL_5621117,
EPI_ISL_5621118, EPI_ISL_5621119, EPI_ISL_5621120, EPI_ISL_5621121, EPI_ISL_5621122, EPI_ISL_5621123, EPI_ISL_5621124, EPI_ISL_5621125, EPI_ISL_5621126, EPI_ISL_5621127, EPI_ISL_5621128, EPI_ISL_5621129, EPI_ISL_5621130, EPI_ISL_5621131, EPI_ISL_5621132, EPI_ISL_5621133, EPI_ISL_5621134, EPI_ISL_5621135, EPI_ISL_5621136, EPI_ISL_5728022, EPI_ISL_5728031, EPI_ISL_5728038,
EPI_ISL_5728039, EPI_ISL_5728040, EPI_ISL_5728043, EPI_ISL_5728044, EPI_ISL_5728051, EPI_ISL_5728052, EPI_ISL_5728060, EPI_ISL_5728061, EPI_ISL_5728062, EPI_ISL_5728071, EPI_ISL_5728072, EPI_ISL_5728077, EPI_ISL_5728078, EPI_ISL_5728079, EPI_ISL_5728086, EPI_ISL_5728087, EPI_ISL_5728094, EPI_ISL_5728095, EPI_ISL_5728096, EPI_ISL_5728099, EPI_ISL_5728100, EPI_ISL_5728103,
EPI_ISL_5728104, EPI_ISL_5728105, EPI_ISL_5728110, EPI_ISL_5728111, EPI_ISL_5728120, EPI_ISL_5728121, EPI_ISL_5728122, EPI_ISL_5728127, EPI_ISL_5728128, EPI_ISL_5728136, EPI_ISL_5728137, EPI_ISL_5728138, EPI_ISL_5728145, EPI_ISL_5728146, EPI_ISL_5728150, EPI_ISL_5728151, EPI_ISL_5728152, EPI_ISL_5728159, EPI_ISL_5728160, EPI_ISL_5728164, EPI_ISL_5728165, EPI_ISL_5728172,
EPI_ISL_5728173, EPI_ISL_5728174, EPI_ISL_5728180, EPI_ISL_5728181, EPI_ISL_5728189, EPI_ISL_5728190, EPI_ISL_5728191, EPI_ISL_5728197, EPI_ISL_5728198, EPI_ISL_5728199, EPI_ISL_5728204, EPI_ISL_5728205, EPI_ISL_5728211, EPI_ISL_5728212, EPI_ISL_5728217, EPI_ISL_5728218, EPI_ISL_5728223, EPI_ISL_5728224, EPI_ISL_5728231, EPI_ISL_5728235, EPI_ISL_5728241, EPI_ISL_5728242,
EPI_ISL_5728248, EPI_ISL_5728249, EPI_ISL_5728250, EPI_ISL_5728258, EPI_ISL_5728259, EPI_ISL_5728269, EPI_ISL_5728270, EPI_ISL_5728271, EPI_ISL_5728277, EPI_ISL_5728278, EPI_ISL_5728281, EPI_ISL_5728282, EPI_ISL_5728283, EPI_ISL_5728287, EPI_ISL_5728288, EPI_ISL_5728289, EPI_ISL_5728298, EPI_ISL_5728299, EPI_ISL_5728305, EPI_ISL_5728306, EPI_ISL_5728307, EPI_ISL_5728314,
EPI_ISL_5728315, EPI_ISL_5728316, EPI_ISL_5728326, EPI_ISL_5728327, EPI_ISL_5728334, EPI_ISL_5728335, EPI_ISL_5728340, EPI_ISL_5728342, EPI_ISL_5728343, EPI_ISL_5728351, EPI_ISL_5728352, EPI_ISL_5728360, EPI_ISL_5728361, EPI_ISL_5728367, EPI_ISL_5728368, EPI_ISL_5728373, EPI_ISL_5728374, EPI_ISL_5728375, EPI_ISL_5728380, EPI_ISL_5728381, EPI_ISL_5728386, EPI_ISL_5728387,
EPI_ISL_5728388, EPI_ISL_5728395, EPI_ISL_5728396, EPI_ISL_5728402, EPI_ISL_5728403, EPI_ISL_5728404, EPI_ISL_5728411, EPI_ISL_5728412, EPI_ISL_5728419, EPI_ISL_5728420, EPI_ISL_5728429, EPI_ISL_5728430, EPI_ISL_5728431, EPI_ISL_5728438, EPI_ISL_5728439, EPI_ISL_5728449, EPI_ISL_5728450, EPI_ISL_5728457, EPI_ISL_5728458, EPI_ISL_5728459, EPI_ISL_5728464, EPI_ISL_5728465,
EPI_ISL_5728471, EPI_ISL_5728472, EPI_ISL_5728473, EPI_ISL_5728480, EPI_ISL_5728483, EPI_ISL_5728484, EPI_ISL_5728485, EPI_ISL_5728492, EPI_ISL_5728493, EPI_ISL_5728494, EPI_ISL_5728504, EPI_ISL_5728505, EPI_ISL_5728507, EPI_ISL_5728508, EPI_ISL_5728513, EPI_ISL_5728514, EPI_ISL_5728515, EPI_ISL_5728523, EPI_ISL_5728524, EPI_ISL_5728527, EPI_ISL_5728528, EPI_ISL_5728535,
EPI_ISL_5728536, EPI_ISL_5728537, EPI_ISL_5728541, EPI_ISL_5728542, EPI_ISL_5728551, EPI_ISL_5728552, EPI_ISL_5728553, EPI_ISL_5728558, EPI_ISL_5728559, EPI_ISL_5728564, EPI_ISL_5728565, EPI_ISL_5728566, EPI_ISL_5728572, EPI_ISL_5728573, EPI_ISL_5728580, EPI_ISL_5728581, EPI_ISL_5728582, EPI_ISL_5728584, EPI_ISL_5728585, EPI_ISL_5728586, EPI_ISL_5728591, EPI_ISL_5728592,
EPI_ISL_5728602, EPI_ISL_5728603, EPI_ISL_5728608, EPI_ISL_5728610, EPI_ISL_5728611, EPI_ISL_5728619, EPI_ISL_5728620, EPI_ISL_5728624, EPI_ISL_5728625, EPI_ISL_5728626, EPI_ISL_5728632, EPI_ISL_5728633, EPI_ISL_5728636, EPI_ISL_5728637, EPI_ISL_5728640, EPI_ISL_5728641, EPI_ISL_5728642, EPI_ISL_5728645, EPI_ISL_5728646, EPI_ISL_5728650, EPI_ISL_5728652, EPI_ISL_5728653,
EPI_ISL_5728658, EPI_ISL_5728659, EPI_ISL_5728660, EPI_ISL_5728667, EPI_ISL_5728668, EPI_ISL_5728673, EPI_ISL_5728674, EPI_ISL_5728675, EPI_ISL_5728681, EPI_ISL_5728682, EPI_ISL_5728689, EPI_ISL_5728690, EPI_ISL_5728691, EPI_ISL_5728694, EPI_ISL_5728695, EPI_ISL_5728701, EPI_ISL_5728702, EPI_ISL_5728703, EPI_ISL_5728710, EPI_ISL_5728711, EPI_ISL_5728712, EPI_ISL_5728718,
EPI_ISL_5728719, EPI_ISL_5728729, EPI_ISL_5728730, EPI_ISL_5728737, EPI_ISL_5728738, EPI_ISL_5728743, EPI_ISL_5728744, EPI_ISL_5728745, EPI_ISL_5728752, EPI_ISL_5728753, EPI_ISL_5728756, EPI_ISL_5728757, EPI_ISL_5728762, EPI_ISL_5728763, EPI_ISL_5728771, EPI_ISL_5728772, EPI_ISL_5728773, EPI_ISL_5728779, EPI_ISL_5728780, EPI_ISL_5728787, EPI_ISL_5728788, EPI_ISL_5728797,
EPI_ISL_5728798, EPI_ISL_5728799, EPI_ISL_5728808, EPI_ISL_5728809, EPI_ISL_5728813, EPI_ISL_5728814, EPI_ISL_5728815, EPI_ISL_5728820, EPI_ISL_5728821, EPI_ISL_5728823, EPI_ISL_5728824, EPI_ISL_5728825, EPI_ISL_5728833, EPI_ISL_5728834, EPI_ISL_5728840, EPI_ISL_5728841, EPI_ISL_5728842, EPI_ISL_5728849, EPI_ISL_5728850, EPI_ISL_5728853, EPI_ISL_5728854, EPI_ISL_5728861,
EPI_ISL_5728862, EPI_ISL_5728863, EPI_ISL_5728867, EPI_ISL_5728868, EPI_ISL_5728874, EPI_ISL_5728875, EPI_ISL_5728876, EPI_ISL_5728878, EPI_ISL_5728879, EPI_ISL_5728887, EPI_ISL_5728890, EPI_ISL_5728891, EPI_ISL_5728897, EPI_ISL_5728898, EPI_ISL_5728903, EPI_ISL_5728904, EPI_ISL_5728911, EPI_ISL_5728912, EPI_ISL_5728920, EPI_ISL_5728921, EPI_ISL_5728930, EPI_ISL_5728931,
EPI_ISL_5728932, EPI_ISL_5728942, EPI_ISL_5728943, EPI_ISL_5728944, EPI_ISL_5728945, EPI_ISL_5728946, EPI_ISL_5728947, EPI_ISL_5728948, EPI_ISL_5728951, EPI_ISL_5728952, EPI_ISL_5728953, EPI_ISL_5728954, EPI_ISL_5728959, EPI_ISL_5728960, EPI_ISL_5728965, EPI_ISL_5728966, EPI_ISL_5728967, EPI_ISL_5728970, EPI_ISL_5728976, EPI_ISL_5728977, EPI_ISL_5728986, EPI_ISL_5728987,
EPI_ISL_5728997, EPI_ISL_5728998, EPI_ISL_5728999, EPI_ISL_5729003, EPI_ISL_5729009, EPI_ISL_5729010, EPI_ISL_5729016, EPI_ISL_5729017, EPI_ISL_5729018, EPI_ISL_5729025, EPI_ISL_5729026, EPI_ISL_5729032, EPI_ISL_5729033, EPI_ISL_5729034, EPI_ISL_5729042, EPI_ISL_5729043, EPI_ISL_5729050, EPI_ISL_5729051, EPI_ISL_5729057, EPI_ISL_5729058, EPI_ISL_5729066, EPI_ISL_5729067,
EPI_ISL_5729073, EPI_ISL_5729074, EPI_ISL_5729075, EPI_ISL_5729085, EPI_ISL_5729086, EPI_ISL_5729094, EPI_ISL_5729095, EPI_ISL_5729104, EPI_ISL_5729105, EPI_ISL_5729108, EPI_ISL_5729109, EPI_ISL_5729110, EPI_ISL_5729119, EPI_ISL_5729120, EPI_ISL_5729126, EPI_ISL_5729127, EPI_ISL_5729136, EPI_ISL_5729137, EPI_ISL_5729138, EPI_ISL_5729142, EPI_ISL_5729143, EPI_ISL_5729149,
EPI_ISL_5729150, EPI_ISL_5729151, EPI_ISL_5729155, EPI_ISL_5729156, EPI_ISL_5729163, EPI_ISL_5729164, EPI_ISL_5729165, EPI_ISL_5729168, EPI_ISL_5729169, EPI_ISL_5729172, EPI_ISL_5729173, EPI_ISL_5729177, EPI_ISL_5729178, EPI_ISL_5729180, EPI_ISL_5729181, EPI_ISL_5729188, EPI_ISL_5729189, EPI_ISL_5729193, EPI_ISL_5729194, EPI_ISL_5729195, EPI_ISL_5729201, EPI_ISL_5729202,
EPI_ISL_5729204, EPI_ISL_5729205, EPI_ISL_5729206, EPI_ISL_5729211, EPI_ISL_5729212, EPI_ISL_5729218, EPI_ISL_5729219, EPI_ISL_5729222, EPI_ISL_5729223, EPI_ISL_5729224, EPI_ISL_5729230, EPI_ISL_5729231, EPI_ISL_5729238, EPI_ISL_5729239, EPI_ISL_5729245, EPI_ISL_5729246, EPI_ISL_5729247, EPI_ISL_5729255, EPI_ISL_5729256, EPI_ISL_5729262, EPI_ISL_5729263, EPI_ISL_5729264,
EPI_ISL_5729269, EPI_ISL_5729270, EPI_ISL_5729272, EPI_ISL_5729278, EPI_ISL_5729279, EPI_ISL_5729285, EPI_ISL_5729286, EPI_ISL_5729293, EPI_ISL_5729294, EPI_ISL_5729295, EPI_ISL_5729301, EPI_ISL_5729302, EPI_ISL_5729303, EPI_ISL_5729307, EPI_ISL_5729308, EPI_ISL_5729314, EPI_ISL_5729315, EPI_ISL_5729316, EPI_ISL_5729323, EPI_ISL_5729324, EPI_ISL_5729329, EPI_ISL_5729330,
EPI_ISL_5729331, EPI_ISL_5729336, EPI_ISL_5729337, EPI_ISL_5729338, EPI_ISL_5729345, EPI_ISL_5729346, EPI_ISL_5729351, EPI_ISL_5729352, EPI_ISL_5729353, EPI_ISL_5729358, EPI_ISL_5729359, EPI_ISL_5729365, EPI_ISL_5729366, EPI_ISL_5729367, EPI_ISL_5729374, EPI_ISL_5729375, EPI_ISL_5729378, EPI_ISL_5729379, EPI_ISL_5729385, EPI_ISL_5729386, EPI_ISL_5729387, EPI_ISL_5729394,
EPI_ISL_5729395, EPI_ISL_5729404, EPI_ISL_5729405, EPI_ISL_5729406, EPI_ISL_5729411, EPI_ISL_5729412, EPI_ISL_5729415, EPI_ISL_5729416, EPI_ISL_5729417, EPI_ISL_5729422, EPI_ISL_5729423, EPI_ISL_5729427, EPI_ISL_5729428, EPI_ISL_5729431, EPI_ISL_5729432, EPI_ISL_5729439, EPI_ISL_5729440, EPI_ISL_5729441, EPI_ISL_5729448, EPI_ISL_5729449, EPI_ISL_5729459, EPI_ISL_5729460,
EPI_ISL_5729461, EPI_ISL_5729468, EPI_ISL_5729469, EPI_ISL_5729471, EPI_ISL_5729472, EPI_ISL_5729473, EPI_ISL_5729478, EPI_ISL_5729479, EPI_ISL_5729480, EPI_ISL_5729486, EPI_ISL_5729487, EPI_ISL_5729496, EPI_ISL_5729497, EPI_ISL_5729498, EPI_ISL_5729502, EPI_ISL_5729503, EPI_ISL_5729511, EPI_ISL_5729512, EPI_ISL_5729513, EPI_ISL_5729520, EPI_ISL_5729521, EPI_ISL_5729527,
EPI_ISL_5729528, EPI_ISL_5729529, EPI_ISL_5729539, EPI_ISL_5729540, EPI_ISL_5729547, EPI_ISL_5729548, EPI_ISL_5729550, EPI_ISL_5729555, EPI_ISL_5729556, EPI_ISL_5729562, EPI_ISL_5729563, EPI_ISL_5729564, EPI_ISL_5729568, EPI_ISL_5729569, EPI_ISL_5729576, EPI_ISL_5729577, EPI_ISL_5729578, EPI_ISL_5729585, EPI_ISL_5729586, EPI_ISL_5729592, EPI_ISL_5729593, EPI_ISL_5729594,
EPI_ISL_5729597, EPI_ISL_5729598, EPI_ISL_5729603, EPI_ISL_5729604, EPI_ISL_5729605, EPI_ISL_5729608, EPI_ISL_5729613, EPI_ISL_5729616, EPI_ISL_5729617, EPI_ISL_5729618, EPI_ISL_5729623, EPI_ISL_5729624, EPI_ISL_5729625, EPI_ISL_5729631, EPI_ISL_5729636, EPI_ISL_5729637, EPI_ISL_5729638, EPI_ISL_5729644, EPI_ISL_5729645, EPI_ISL_5729649, EPI_ISL_5729650, EPI_ISL_5729651,
EPI_ISL_5729657, EPI_ISL_5729658, EPI_ISL_5729665, EPI_ISL_5729666, EPI_ISL_5729667, EPI_ISL_5729669, EPI_ISL_5729670, EPI_ISL_5729673, EPI_ISL_5729674, EPI_ISL_5729675, EPI_ISL_5729683, EPI_ISL_5729684, EPI_ISL_5729688, EPI_ISL_5729689, EPI_ISL_5729690, EPI_ISL_5729695, EPI_ISL_5729696, EPI_ISL_5729698, EPI_ISL_5729699, EPI_ISL_5729700, EPI_ISL_5729707, EPI_ISL_5729708,
EPI_ISL_5729709, EPI_ISL_5729714, EPI_ISL_5729715, EPI_ISL_5729722, EPI_ISL_5729723, EPI_ISL_5729724, EPI_ISL_5729728, EPI_ISL_5729729, EPI_ISL_5729732, EPI_ISL_5729733, EPI_ISL_5729734, EPI_ISL_5729741, EPI_ISL_5729742, EPI_ISL_5729748, EPI_ISL_5729749, EPI_ISL_5729750, EPI_ISL_5729754, EPI_ISL_5729755, EPI_ISL_5729764, EPI_ISL_5729765, EPI_ISL_5729766, EPI_ISL_5729774,
EPI_ISL_5729775, EPI_ISL_5729781, EPI_ISL_5729782, EPI_ISL_5729783, EPI_ISL_5729786, EPI_ISL_5729787, EPI_ISL_5729788, EPI_ISL_5729794, EPI_ISL_5729795, EPI_ISL_5729802, EPI_ISL_5729803, EPI_ISL_5729809, EPI_ISL_5729810, EPI_ISL_5729811, EPI_ISL_5729815, EPI_ISL_5729816, EPI_ISL_5729820, EPI_ISL_5729821, EPI_ISL_5729822, EPI_ISL_5729830, EPI_ISL_5729831, EPI_ISL_5729838,
EPI_ISL_5729839, EPI_ISL_5729840, EPI_ISL_5729845, EPI_ISL_5729846, EPI_ISL_5729852, EPI_ISL_5729853, EPI_ISL_5729856, EPI_ISL_5729857, EPI_ISL_5729863, EPI_ISL_5729864, EPI_ISL_5729865, EPI_ISL_5729873, EPI_ISL_5729874, EPI_ISL_5729883, EPI_ISL_5729884, EPI_ISL_5729885, EPI_ISL_5729891, EPI_ISL_5729892, EPI_ISL_5729900, EPI_ISL_5729901, EPI_ISL_5729902, EPI_ISL_5729911,
EPI_ISL_5729912, EPI_ISL_5729919, EPI_ISL_5729920, EPI_ISL_5729921, EPI_ISL_5729925, EPI_ISL_5729926, EPI_ISL_5729934, EPI_ISL_5729935, EPI_ISL_5729936, EPI_ISL_5729945, EPI_ISL_5729953, EPI_ISL_5729954, EPI_ISL_5729955, EPI_ISL_5729964, EPI_ISL_5729965, EPI_ISL_5729971, EPI_ISL_5729972, EPI_ISL_5729977, EPI_ISL_5729986, EPI_ISL_5729987, EPI_ISL_5729990, EPI_ISL_5729991,
EPI_ISL_5729992, EPI_ISL_5729997, EPI_ISL_5729998, EPI_ISL_5730000, EPI_ISL_5730001, EPI_ISL_5730002, EPI_ISL_5730010, EPI_ISL_5730011, EPI_ISL_5730019, EPI_ISL_5730020, EPI_ISL_5730021, EPI_ISL_5730029, EPI_ISL_5730033, EPI_ISL_5730034, EPI_ISL_5730035, EPI_ISL_5730037, EPI_ISL_5730038, EPI_ISL_5730048, EPI_ISL_5730049, EPI_ISL_5730053, EPI_ISL_5730054, EPI_ISL_5730059,
EPI_ISL_5730060, EPI_ISL_5730061, EPI_ISL_5730065, EPI_ISL_5730069, EPI_ISL_5730070, EPI_ISL_5730071, EPI_ISL_5730074, EPI_ISL_5730075, EPI_ISL_5730079, EPI_ISL_5730080, EPI_ISL_5730081, EPI_ISL_5730088, EPI_ISL_5730089, EPI_ISL_5730096, EPI_ISL_5730097, EPI_ISL_5730098, EPI_ISL_5730103, EPI_ISL_5730104, EPI_ISL_5730112, EPI_ISL_5730113, EPI_ISL_5730114, EPI_ISL_5730120,
EPI_ISL_5730121, EPI_ISL_5730123, EPI_ISL_5730130, EPI_ISL_5730137, EPI_ISL_5730138, EPI_ISL_5730142, EPI_ISL_5730143, EPI_ISL_5730149, EPI_ISL_5730150, EPI_ISL_5730151, EPI_ISL_5730160, EPI_ISL_5730169, EPI_ISL_5730170, EPI_ISL_5730171, EPI_ISL_5730178, EPI_ISL_5730179, EPI_ISL_5730184, EPI_ISL_5730185, EPI_ISL_5730186, EPI_ISL_5730194, EPI_ISL_5730195, EPI_ISL_5730198,
EPI_ISL_5730199, EPI_ISL_5730200, EPI_ISL_5730203, EPI_ISL_5730204, EPI_ISL_5730210, EPI_ISL_5730211, EPI_ISL_5730212, EPI_ISL_5730218, EPI_ISL_5730223, EPI_ISL_5730224, EPI_ISL_5730225, EPI_ISL_5730229, EPI_ISL_5730230, EPI_ISL_5730234, EPI_ISL_5730237, EPI_ISL_5730238, EPI_ISL_5730245, EPI_ISL_5730246, EPI_ISL_5730254, EPI_ISL_5730261, EPI_ISL_5730262, EPI_ISL_5730263,
EPI_ISL_5730272, EPI_ISL_5730278, EPI_ISL_5730279, EPI_ISL_5730280, EPI_ISL_5730287, EPI_ISL_5730288, EPI_ISL_5730294, EPI_ISL_5730295, EPI_ISL_5730296, EPI_ISL_5730299, EPI_ISL_5730300, EPI_ISL_5730301, EPI_ISL_5730303, EPI_ISL_5730305, EPI_ISL_5730313, EPI_ISL_5730314, EPI_ISL_5730320, EPI_ISL_5730321, EPI_ISL_5730322, EPI_ISL_5730328, EPI_ISL_5730329, EPI_ISL_5730337,
EPI_ISL_5730345, EPI_ISL_5730346, EPI_ISL_5730354, EPI_ISL_5730355, EPI_ISL_5730363, EPI_ISL_5730365, EPI_ISL_5730375, EPI_ISL_5730376, EPI_ISL_5730382, EPI_ISL_5730386, EPI_ISL_5730387, EPI_ISL_5730388, EPI_ISL_5730392, EPI_ISL_5730813, EPI_ISL_5730814, EPI_ISL_5730823, EPI_ISL_5730824, EPI_ISL_5730828, EPI_ISL_5730829, EPI_ISL_5730830, EPI_ISL_5730833, EPI_ISL_5730834,
EPI_ISL_5730842, EPI_ISL_5730843, EPI_ISL_5730850, EPI_ISL_5730851, EPI_ISL_5730852, EPI_ISL_5730858, EPI_ISL_5730859, EPI_ISL_5730866, EPI_ISL_5730867, EPI_ISL_5730875, EPI_ISL_5730876, EPI_ISL_5730877, EPI_ISL_5730886, EPI_ISL_5730887, EPI_ISL_5730893, EPI_ISL_5730894, EPI_ISL_5730895, EPI_ISL_5730902, EPI_ISL_5730903, EPI_ISL_5730909, EPI_ISL_5730910, EPI_ISL_5730914,
EPI_ISL_5730915, EPI_ISL_5730916, EPI_ISL_5730922, EPI_ISL_5730923, EPI_ISL_5730929, EPI_ISL_5730930, EPI_ISL_5730934, EPI_ISL_5730935, EPI_ISL_5730945, EPI_ISL_5730946, EPI_ISL_5730948, EPI_ISL_5730949, EPI_ISL_5730950, EPI_ISL_5730957, EPI_ISL_5730958, EPI_ISL_5730964, EPI_ISL_5730965, EPI_ISL_5730966, EPI_ISL_5730972, EPI_ISL_5730973, EPI_ISL_5730980, EPI_ISL_5730981,
EPI_ISL_5730982, EPI_ISL_5730988, EPI_ISL_5730989, EPI_ISL_5730994, EPI_ISL_5730995, EPI_ISL_5730996, EPI_ISL_5731001, EPI_ISL_5731002, EPI_ISL_5731008, EPI_ISL_5731009, EPI_ISL_5731010, EPI_ISL_5731016, EPI_ISL_5731017, EPI_ISL_5731025, EPI_ISL_5731026, EPI_ISL_5731035, EPI_ISL_5731036, EPI_ISL_5731037, EPI_ISL_5731044, EPI_ISL_5731045, EPI_ISL_5731049, EPI_ISL_5731050,
EPI_ISL_5731051, EPI_ISL_5731054, EPI_ISL_5731056, EPI_ISL_5731065, EPI_ISL_5731066, EPI_ISL_5731067, EPI_ISL_5731075, EPI_ISL_5731076, EPI_ISL_5731077, EPI_ISL_5731083, EPI_ISL_5731084, EPI_ISL_5731092, EPI_ISL_5731093, EPI_ISL_5731094, EPI_ISL_5731102, EPI_ISL_5731103, EPI_ISL_5731111, EPI_ISL_5731112, EPI_ISL_5731113, EPI_ISL_5731120, EPI_ISL_5731121, EPI_ISL_5731122,
EPI_ISL_5731130, EPI_ISL_5731131, EPI_ISL_5731137, EPI_ISL_5731138, EPI_ISL_5731143, EPI_ISL_5731144, EPI_ISL_5731145, EPI_ISL_5731151, EPI_ISL_5731152, EPI_ISL_5731158, EPI_ISL_5731159, EPI_ISL_5731160, EPI_ISL_5731164, EPI_ISL_5731165, EPI_ISL_5731167, EPI_ISL_5731171, EPI_ISL_5731172, EPI_ISL_5731180, EPI_ISL_5731181, EPI_ISL_5731186, EPI_ISL_5731187, EPI_ISL_5731188,
EPI_ISL_5731193, EPI_ISL_5731194, EPI_ISL_5731198, EPI_ISL_5731199, EPI_ISL_5731200, EPI_ISL_5731204, EPI_ISL_5731205, EPI_ISL_5731209, EPI_ISL_5731210, EPI_ISL_5731211, EPI_ISL_5731219, EPI_ISL_5731220, EPI_ISL_5731227, EPI_ISL_5731228, EPI_ISL_5731229, EPI_ISL_5731235, EPI_ISL_5731236, EPI_ISL_5731237, EPI_ISL_5731246, EPI_ISL_5731247, EPI_ISL_5731253, EPI_ISL_5731254,
EPI_ISL_5731255, EPI_ISL_5731262, EPI_ISL_5731263, EPI_ISL_5731269, EPI_ISL_5731270, EPI_ISL_5731271, EPI_ISL_5731275, EPI_ISL_5731276, EPI_ISL_5731279, EPI_ISL_5731280, EPI_ISL_5731281, EPI_ISL_5731289, EPI_ISL_5731290, EPI_ISL_5731295, EPI_ISL_5731296, EPI_ISL_5731300, EPI_ISL_5731301, EPI_ISL_5731309, EPI_ISL_5731310, EPI_ISL_5731311, EPI_ISL_5731316, EPI_ISL_5731317,
EPI_ISL_5731325, EPI_ISL_5731326, EPI_ISL_5731327, EPI_ISL_5731335, EPI_ISL_5731336, EPI_ISL_5731342, EPI_ISL_5731348, EPI_ISL_5731349, EPI_ISL_5731355, EPI_ISL_5731356, EPI_ISL_5731360, EPI_ISL_5731361, EPI_ISL_5731362, EPI_ISL_5731368, EPI_ISL_5731369, EPI_ISL_5731374, EPI_ISL_5731375, EPI_ISL_5731385, EPI_ISL_5731386, EPI_ISL_5731387, EPI_ISL_5731397, EPI_ISL_5731398,
EPI_ISL_5731407, EPI_ISL_5731408, EPI_ISL_5731412, EPI_ISL_5731413, EPI_ISL_5731414, EPI_ISL_5731420, EPI_ISL_5731421, EPI_ISL_5731425, EPI_ISL_5731426, EPI_ISL_5731427, EPI_ISL_5731431, EPI_ISL_5731432, EPI_ISL_5731437, EPI_ISL_5731438, EPI_ISL_5731445, EPI_ISL_5731446, EPI_ISL_5731447, EPI_ISL_5731456, EPI_ISL_5731457, EPI_ISL_5731464, EPI_ISL_5731465, EPI_ISL_5731470,
EPI_ISL_5731471, EPI_ISL_5731476, EPI_ISL_5731477, EPI_ISL_5731478, EPI_ISL_5731481, EPI_ISL_5731482, EPI_ISL_5731486, EPI_ISL_5731487, EPI_ISL_5731488, EPI_ISL_5731493, EPI_ISL_5731494, EPI_ISL_5731500, EPI_ISL_5731501, EPI_ISL_5731502, EPI_ISL_5731506, EPI_ISL_5731507, EPI_ISL_5731510, EPI_ISL_5731511, EPI_ISL_5731515, EPI_ISL_5731516, EPI_ISL_5731519, EPI_ISL_5731520,
EPI_ISL_5731521, EPI_ISL_5731528, EPI_ISL_5731529, EPI_ISL_5731534, EPI_ISL_5731535, EPI_ISL_5731536, EPI_ISL_5731542, EPI_ISL_5731543, EPI_ISL_5731546, EPI_ISL_5731550, EPI_ISL_5731551, EPI_ISL_5731557, EPI_ISL_5731558, EPI_ISL_5731559, EPI_ISL_5731564, EPI_ISL_5731565, EPI_ISL_5731570, EPI_ISL_5731571, EPI_ISL_5731572, EPI_ISL_5731580, EPI_ISL_5731581, EPI_ISL_5731586,
EPI_ISL_5731587, EPI_ISL_5731588, EPI_ISL_5731591, EPI_ISL_5731592, EPI_ISL_5731596, EPI_ISL_5731597, EPI_ISL_5731601, EPI_ISL_5731602, EPI_ISL_5731603, EPI_ISL_5731606, EPI_ISL_5731607, EPI_ISL_5731608, EPI_ISL_5731610, EPI_ISL_5731611, EPI_ISL_5731616, EPI_ISL_5731617, EPI_ISL_5731624, EPI_ISL_5731625, EPI_ISL_5731626, EPI_ISL_5731632, EPI_ISL_5731633, EPI_ISL_5731638,
EPI_ISL_5731639, EPI_ISL_5731640, EPI_ISL_5731646, EPI_ISL_5731648, EPI_ISL_5731654, EPI_ISL_5731663, EPI_ISL_5731664, EPI_ISL_5731670, EPI_ISL_5731678, EPI_ISL_5731679, EPI_ISL_5731680, EPI_ISL_5731686, EPI_ISL_5731687, EPI_ISL_5731694, EPI_ISL_5731695, EPI_ISL_5731704, EPI_ISL_5731705, EPI_ISL_5731706, EPI_ISL_5731714, EPI_ISL_5731715, EPI_ISL_5731723, EPI_ISL_5731724,
EPI_ISL_5731725, EPI_ISL_5731731, EPI_ISL_5731732, EPI_ISL_5731741, EPI_ISL_5731742, EPI_ISL_5731743, EPI_ISL_5731749, EPI_ISL_5731750, EPI_ISL_5731756, EPI_ISL_5731757, EPI_ISL_5731758, EPI_ISL_5731763, EPI_ISL_5731764, EPI_ISL_5731767, EPI_ISL_5731768, EPI_ISL_5731769, EPI_ISL_5731773, EPI_ISL_5731774, EPI_ISL_5731782, EPI_ISL_5731783, EPI_ISL_5731784, EPI_ISL_5731790,
EPI_ISL_5731791, EPI_ISL_5731793, EPI_ISL_5731794, EPI_ISL_5731795, EPI_ISL_5731801, EPI_ISL_5731802, EPI_ISL_5731808, EPI_ISL_5731809, EPI_ISL_5731810, EPI_ISL_5731815, EPI_ISL_5731816, EPI_ISL_5731825, EPI_ISL_5731826, EPI_ISL_5731827, EPI_ISL_5731832, EPI_ISL_5731833, EPI_ISL_5731840, EPI_ISL_5731841, EPI_ISL_5731842, EPI_ISL_5731848, EPI_ISL_5731849, EPI_ISL_5731850,
EPI_ISL_5731858, EPI_ISL_5731859, EPI_ISL_5731862, EPI_ISL_5731863, EPI_ISL_5731864, EPI_ISL_5731870, EPI_ISL_5731871, EPI_ISL_5731877, EPI_ISL_5731878, EPI_ISL_5731881, EPI_ISL_5731882, EPI_ISL_5731883, EPI_ISL_5731889, EPI_ISL_5731890, EPI_ISL_5731891, EPI_ISL_5731895, EPI_ISL_5731896, EPI_ISL_5731905, EPI_ISL_5731906, EPI_ISL_5731907, EPI_ISL_5731913, EPI_ISL_5731914,
EPI_ISL_5731921, EPI_ISL_5731922, EPI_ISL_5731923, EPI_ISL_5731931, EPI_ISL_5731932, EPI_ISL_5731937, EPI_ISL_5731938, EPI_ISL_5731948, EPI_ISL_5731949, EPI_ISL_5731950, EPI_ISL_5731957, EPI_ISL_5731958, EPI_ISL_5731959, EPI_ISL_5731966, EPI_ISL_5731967, EPI_ISL_5731972, EPI_ISL_5731973, EPI_ISL_5731982, EPI_ISL_5731983, EPI_ISL_5731984, EPI_ISL_5731988, EPI_ISL_5731989,
EPI_ISL_5731997, EPI_ISL_5731998, EPI_ISL_5731999, EPI_ISL_5732008, EPI_ISL_5732009, EPI_ISL_5732010, EPI_ISL_5732019, EPI_ISL_5732020, EPI_ISL_5732024, EPI_ISL_5732025, EPI_ISL_5732026, EPI_ISL_5732029, EPI_ISL_5732030, EPI_ISL_5732031, EPI_ISL_5732038, EPI_ISL_5732039, EPI_ISL_5732043, EPI_ISL_5732044, EPI_ISL_5732045, EPI_ISL_5732051, EPI_ISL_5732052, EPI_ISL_5732060,
EPI_ISL_5732061, EPI_ISL_5732062, EPI_ISL_5732068, EPI_ISL_5732069, EPI_ISL_5732072, EPI_ISL_5732073, EPI_ISL_5732074, EPI_ISL_5732077, EPI_ISL_5732078, EPI_ISL_5732079

| | | | |
|---|---|---|---|
| see above | Israel Central Virology laboratory | Israel Central Virology laboratory | Dana Bar-Ilan; Danit Sofer; Efrat Dahan Bucris; Efrat Glick-Saar; Ella Mendelson; Gideon Rechavi; Michal Mandelboim; Miranda Geva; Neta Zuckerman; Omer Asraf; Oran Erster; Orna Mor; Yaniv Lustig |

EPI_ISL_2182923, EPI_ISL_2183026, EPI_ISL_2183029, EPI_ISL_2183057, EPI_ISL_2183059, EPI_ISL_2183060, EPI_ISL_2183062, EPI_ISL_2183112, EPI_ISL_2183114, EPI_ISL_2183116, EPI_ISL_2183117, EPI_ISL_2183341, EPI_ISL_2183420, EPI_ISL_2183422, EPI_ISL_2183424, EPI_ISL_2183427, EPI_ISL_2183429, EPI_ISL_2183431, EPI_ISL_2183433, EPI_ISL_2183613, EPI_ISL_2183615, EPI_ISL_2183679,
EPI_ISL_2183691, EPI_ISL_2183692, EPI_ISL_2183694, EPI_ISL_2183702, EPI_ISL_2183704, EPI_ISL_2183706, EPI_ISL_2183721, EPI_ISL_2183727, EPI_ISL_2183736, EPI_ISL_2183749, EPI_ISL_2183777, EPI_ISL_2183779, EPI_ISL_2183802, EPI_ISL_2183805, EPI_ISL_2636161, EPI_ISL_2636166, EPI_ISL_2636189, EPI_ISL_2636204, EPI_ISL_2636241, EPI_ISL_2636298, EPI_ISL_2636309, EPI_ISL_2636326,
EPI_ISL_2636333, EPI_ISL_2636334, EPI_ISL_2636336, EPI_ISL_2636343, EPI_ISL_2636345, EPI_ISL_2636347, EPI_ISL_2636353, EPI_ISL_2636354, EPI_ISL_2636356, EPI_ISL_2636358, EPI_ISL_2636361, EPI_ISL_2636363, EPI_ISL_2636365, EPI_ISL_2636367, EPI_ISL_2636368, EPI_ISL_2636371, EPI_ISL_2636373, EPI_ISL_2636377, EPI_ISL_2636379, EPI_ISL_2636383, EPI_ISL_2636385, EPI_ISL_2636387,
EPI_ISL_2636394, EPI_ISL_2636396, EPI_ISL_2636399, EPI_ISL_3278304, EPI_ISL_3278305, EPI_ISL_3278306, EPI_ISL_3278308, EPI_ISL_3278310, EPI_ISL_3278311, EPI_ISL_3278312, EPI_ISL_3278313, EPI_ISL_3278314, EPI_ISL_3278315, EPI_ISL_3278316, EPI_ISL_3278317, EPI_ISL_3278318, EPI_ISL_3278319, EPI_ISL_3278320, EPI_ISL_3278321, EPI_ISL_3278322, EPI_ISL_3278324, EPI_ISL_3278325,
EPI_ISL_3278326, EPI_ISL_3278327, EPI_ISL_3278328, EPI_ISL_3278329, EPI_ISL_3278330, EPI_ISL_3278331, EPI_ISL_3278334, EPI_ISL_3278335, EPI_ISL_3278336, EPI_ISL_3278337, EPI_ISL_3278338, EPI_ISL_3278339, EPI_ISL_3278340, EPI_ISL_3278341, EPI_ISL_3278342, EPI_ISL_3278343, EPI_ISL_3278344, EPI_ISL_3278345, EPI_ISL_3278346, EPI_ISL_3278350, EPI_ISL_3278351, EPI_ISL_3278352,
EPI_ISL_3278353, EPI_ISL_3278354, EPI_ISL_3278355, EPI_ISL_3278356, EPI_ISL_3278357, EPI_ISL_3278358, EPI_ISL_3278359, EPI_ISL_3278361, EPI_ISL_3278362, EPI_ISL_3278363, EPI_ISL_3278364, EPI_ISL_3278365, EPI_ISL_3278366, EPI_ISL_3278367, EPI_ISL_3278369, EPI_ISL_3278370, EPI_ISL_3278372, EPI_ISL_3278373, EPI_ISL_3278374, EPI_ISL_3278375, EPI_ISL_3278376, EPI_ISL_3278377,
EPI_ISL_3278378, EPI_ISL_3278379, EPI_ISL_3278380, EPI_ISL_3278381, EPI_ISL_3278382, EPI_ISL_3278383, EPI_ISL_3278384, EPI_ISL_3278385, EPI_ISL_3278386, EPI_ISL_3278387, EPI_ISL_3278388, EPI_ISL_3278389, EPI_ISL_3278390, EPI_ISL_3278391, EPI_ISL_3278392, EPI_ISL_3278393, EPI_ISL_3278394, EPI_ISL_3278395, EPI_ISL_3278396, EPI_ISL_3278397, EPI_ISL_3278398, EPI_ISL_3278399,
EPI_ISL_3278400, EPI_ISL_3278401, EPI_ISL_3278402, EPI_ISL_3278403, EPI_ISL_3278404, EPI_ISL_3278405, EPI_ISL_3278406, EPI_ISL_3278407, EPI_ISL_3278408, EPI_ISL_3278409, EPI_ISL_3278410, EPI_ISL_3278412, EPI_ISL_3278414, EPI_ISL_3278415, EPI_ISL_3278416, EPI_ISL_3278417, EPI_ISL_3278418, EPI_ISL_3278419, EPI_ISL_3278420, EPI_ISL_3278421, EPI_ISL_3278423, EPI_ISL_3278424,
EPI_ISL_3278425, EPI_ISL_3278426, EPI_ISL_3278427, EPI_ISL_3278428, EPI_ISL_3278429, EPI_ISL_3278430, EPI_ISL_3278432, EPI_ISL_3278433, EPI_ISL_3278434, EPI_ISL_3278435, EPI_ISL_3278436, EPI_ISL_3278437, EPI_ISL_3278438, EPI_ISL_3278439, EPI_ISL_3278440, EPI_ISL_3278441, EPI_ISL_3278442, EPI_ISL_3278444, EPI_ISL_3278445, EPI_ISL_3278446, EPI_ISL_3278447, EPI_ISL_3278448,
EPI_ISL_3278449, EPI_ISL_3278451, EPI_ISL_3278452, EPI_ISL_3278453, EPI_ISL_3278454, EPI_ISL_3278455, EPI_ISL_3278456, EPI_ISL_3278458, EPI_ISL_3278459, EPI_ISL_3278461, EPI_ISL_3278462, EPI_ISL_3278463, EPI_ISL_3278464, EPI_ISL_3278465, EPI_ISL_3278466, EPI_ISL_3278467, EPI_ISL_3278468, EPI_ISL_3278469, EPI_ISL_3278470, EPI_ISL_3278471, EPI_ISL_3278472, EPI_ISL_3278473,
EPI_ISL_3278474, EPI_ISL_3278475, EPI_ISL_3278476, EPI_ISL_3278477, EPI_ISL_3278478, EPI_ISL_3278479, EPI_ISL_3278480, EPI_ISL_3278481, EPI_ISL_3278482, EPI_ISL_3278483, EPI_ISL_3278484, EPI_ISL_3278485, EPI_ISL_3278486, EPI_ISL_3278487, EPI_ISL_3278488, EPI_ISL_3278489, EPI_ISL_3278490, EPI_ISL_3278491, EPI_ISL_3278492, EPI_ISL_3278493, EPI_ISL_3278494, EPI_ISL_3278495,
EPI_ISL_3278496, EPI_ISL_3278497, EPI_ISL_3278498, EPI_ISL_3278499, EPI_ISL_3278500, EPI_ISL_3278501, EPI_ISL_3278502, EPI_ISL_3278503, EPI_ISL_3278504, EPI_ISL_3278505, EPI_ISL_3278506, EPI_ISL_3278507, EPI_ISL_3278509, EPI_ISL_3278510, EPI_ISL_3278511, EPI_ISL_3278512, EPI_ISL_3278513, EPI_ISL_3278514, EPI_ISL_3278515, EPI_ISL_3278516, EPI_ISL_3278517, EPI_ISL_3278518,
EPI_ISL_3278519, EPI_ISL_3278520, EPI_ISL_3278521, EPI_ISL_3278522, EPI_ISL_3278523, EPI_ISL_3278524, EPI_ISL_3278525, EPI_ISL_3278526, EPI_ISL_3278528, EPI_ISL_3278529, EPI_ISL_3278530, EPI_ISL_3278531, EPI_ISL_3278532, EPI_ISL_3278533, EPI_ISL_3278534, EPI_ISL_3278535, EPI_ISL_3278536, EPI_ISL_3278537, EPI_ISL_3278538, EPI_ISL_3278539, EPI_ISL_3278540, EPI_ISL_3278541,
EPI_ISL_3278542, EPI_ISL_3278543, EPI_ISL_3278544, EPI_ISL_3278545, EPI_ISL_3278546, EPI_ISL_3278547, EPI_ISL_3278548, EPI_ISL_3278549, EPI_ISL_3278550, EPI_ISL_3278551, EPI_ISL_3278554, EPI_ISL_3278555, EPI_ISL_3278556, EPI_ISL_3278557, EPI_ISL_3278558, EPI_ISL_3278559, EPI_ISL_3278560, EPI_ISL_3278562, EPI_ISL_3278563, EPI_ISL_3278565, EPI_ISL_3278566, EPI_ISL_3278567,
EPI_ISL_3278568, EPI_ISL_3278569, EPI_ISL_3278570, EPI_ISL_3278571, EPI_ISL_3278572, EPI_ISL_3278573, EPI_ISL_3278574, EPI_ISL_3278575, EPI_ISL_3278576, EPI_ISL_3278577, EPI_ISL_3278578, EPI_ISL_3278579, EPI_ISL_3278580, EPI_ISL_3278581, EPI_ISL_3278582, EPI_ISL_3278583, EPI_ISL_3278584, EPI_ISL_3278585, EPI_ISL_3278586, EPI_ISL_3278587, EPI_ISL_3278588, EPI_ISL_3278590,
EPI_ISL_3278591, EPI_ISL_3278593, EPI_ISL_3278594, EPI_ISL_3278595, EPI_ISL_3278597, EPI_ISL_3278598, EPI_ISL_3278599, EPI_ISL_3278600, EPI_ISL_3278601, EPI_ISL_3278603, EPI_ISL_3278604, EPI_ISL_3278606, EPI_ISL_3278607, EPI_ISL_3278608, EPI_ISL_3278609, EPI_ISL_3278610, EPI_ISL_3278611, EPI_ISL_3278612, EPI_ISL_3278614, EPI_ISL_3278615, EPI_ISL_3278616, EPI_ISL_3278617,
EPI_ISL_3278619, EPI_ISL_3278620, EPI_ISL_3278623, EPI_ISL_3278625, EPI_ISL_3278626, EPI_ISL_3278628, EPI_ISL_3278629, EPI_ISL_3278630, EPI_ISL_3278631, EPI_ISL_3278632, EPI_ISL_3278633, EPI_ISL_3278634, EPI_ISL_3278635, EPI_ISL_3278636, EPI_ISL_3278637, EPI_ISL_3278638, EPI_ISL_3278639, EPI_ISL_3278640, EPI_ISL_3278641, EPI_ISL_3278642, EPI_ISL_3278643, EPI_ISL_3278644,
EPI_ISL_3278645, EPI_ISL_3278646, EPI_ISL_3278647, EPI_ISL_3278648, EPI_ISL_3278649, EPI_ISL_3278652, EPI_ISL_3278653, EPI_ISL_3278654, EPI_ISL_3278655, EPI_ISL_3278656, EPI_ISL_3278658, EPI_ISL_3278659, EPI_ISL_3278660, EPI_ISL_3278661, EPI_ISL_3278662, EPI_ISL_3278663, EPI_ISL_3278664, EPI_ISL_3278665, EPI_ISL_3278666, EPI_ISL_3278667, EPI_ISL_3278668, EPI_ISL_3278669,
EPI_ISL_3278670, EPI_ISL_3278671, EPI_ISL_3278672, EPI_ISL_3278673, EPI_ISL_3278674, EPI_ISL_3278675, EPI_ISL_3278676, EPI_ISL_3278677, EPI_ISL_3278678, EPI_ISL_3278679, EPI_ISL_3278680, EPI_ISL_3278681, EPI_ISL_3278682, EPI_ISL_3278683, EPI_ISL_3278684, EPI_ISL_3278685, EPI_ISL_3278686, EPI_ISL_3278687, EPI_ISL_3278688, EPI_ISL_3278689, EPI_ISL_3278690, EPI_ISL_3278691,
EPI_ISL_3278692, EPI_ISL_3278693, EPI_ISL_3278694, EPI_ISL_3278695, EPI_ISL_3278696, EPI_ISL_3278697, EPI_ISL_3278698, EPI_ISL_3278699, EPI_ISL_3278700, EPI_ISL_3278701, EPI_ISL_3278702, EPI_ISL_3278703, EPI_ISL_3278704, EPI_ISL_3278705, EPI_ISL_3278706, EPI_ISL_3278707, EPI_ISL_3278708, EPI_ISL_3278709, EPI_ISL_3278710, EPI_ISL_3278711, EPI_ISL_3278712, EPI_ISL_3278713,
EPI_ISL_3278714, EPI_ISL_3278715, EPI_ISL_3278716, EPI_ISL_3278718, EPI_ISL_3278721, EPI_ISL_3278723, EPI_ISL_3278724, EPI_ISL_3278726, EPI_ISL_3278727, EPI_ISL_3278728, EPI_ISL_3278729, EPI_ISL_3278730, EPI_ISL_3278731, EPI_ISL_3278732, EPI_ISL_3278733, EPI_ISL_3278734, EPI_ISL_3278735, EPI_ISL_3278736, EPI_ISL_3278737, EPI_ISL_3278739, EPI_ISL_3278740, EPI_ISL_3278741,
EPI_ISL_3278743, EPI_ISL_3278744, EPI_ISL_3278745, EPI_ISL_3278746, EPI_ISL_3278747, EPI_ISL_3278748, EPI_ISL_3278749, EPI_ISL_3278750, EPI_ISL_3278752, EPI_ISL_3278753, EPI_ISL_3278754, EPI_ISL_3278755, EPI_ISL_3278756, EPI_ISL_3278757, EPI_ISL_3278758, EPI_ISL_3278759, EPI_ISL_3278760, EPI_ISL_3278761, EPI_ISL_3278762, EPI_ISL_3278763, EPI_ISL_3278764, EPI_ISL_3278765,
EPI_ISL_3278766, EPI_ISL_3278767, EPI_ISL_3278768, EPI_ISL_3278769, EPI_ISL_3278770, EPI_ISL_3278771, EPI_ISL_3278772, EPI_ISL_3278773, EPI_ISL_3278774, EPI_ISL_3278775, EPI_ISL_3278776, EPI_ISL_3278778, EPI_ISL_3278779, EPI_ISL_3278780, EPI_ISL_3278781, EPI_ISL_3278782, EPI_ISL_3278783, EPI_ISL_3278784, EPI_ISL_3278785, EPI_ISL_3278786, EPI_ISL_3278787, EPI_ISL_3278788,
EPI_ISL_3278789, EPI_ISL_3278790, EPI_ISL_3278791, EPI_ISL_3278793, EPI_ISL_3278794, EPI_ISL_3278795, EPI_ISL_3278796, EPI_ISL_3278797, EPI_ISL_3278798, EPI_ISL_3278799, EPI_ISL_3278800, EPI_ISL_3278801, EPI_ISL_3278802, EPI_ISL_3278803, EPI_ISL_3278804, EPI_ISL_3278805, EPI_ISL_3278806, EPI_ISL_3278807, EPI_ISL_3278808, EPI_ISL_3278809, EPI_ISL_3278810, EPI_ISL_3278811,
EPI_ISL_3278812, EPI_ISL_3278813, EPI_ISL_3278814, EPI_ISL_3278815, EPI_ISL_3278816, EPI_ISL_3278817, EPI_ISL_3278818, EPI_ISL_3278819, EPI_ISL_3278820, EPI_ISL_3278821, EPI_ISL_3278822, EPI_ISL_3278823, EPI_ISL_3278824, EPI_ISL_3278825, EPI_ISL_3278826, EPI_ISL_3278827, EPI_ISL_3278828, EPI_ISL_3278829, EPI_ISL_3278830, EPI_ISL_3278831, EPI_ISL_3278832, EPI_ISL_3278833,

EPI_ISL_3278834, EPI_ISL_3278835, EPI_ISL_3278836, EPI_ISL_3278837, EPI_ISL_3278838, EPI_ISL_3278839, EPI_ISL_3278841, EPI_ISL_3278842, EPI_ISL_3278844, EPI_ISL_3278845, EPI_ISL_3278846, EPI_ISL_3278847, EPI_ISL_3278849, EPI_ISL_3278850, EPI_ISL_3278851, EPI_ISL_3278852, EPI_ISL_3278853, EPI_ISL_3278854, EPI_ISL_3278855, EPI_ISL_3278856, EPI_ISL_3278857, EPI_ISL_3278858,
EPI_ISL_3278859, EPI_ISL_3278860, EPI_ISL_3278861, EPI_ISL_3278862, EPI_ISL_3278863, EPI_ISL_3278864, EPI_ISL_3278865, EPI_ISL_3278866, EPI_ISL_3278867, EPI_ISL_3278868, EPI_ISL_3278869, EPI_ISL_3278870, EPI_ISL_3278871, EPI_ISL_3278872, EPI_ISL_3278873, EPI_ISL_3278874, EPI_ISL_3278877, EPI_ISL_3278878, EPI_ISL_3278880, EPI_ISL_3278881, EPI_ISL_3278882, EPI_ISL_3278883,
EPI_ISL_3278884, EPI_ISL_3278885, EPI_ISL_3278886, EPI_ISL_3278887, EPI_ISL_3278888, EPI_ISL_3278889, EPI_ISL_3278890, EPI_ISL_3278891, EPI_ISL_3278892, EPI_ISL_3278893, EPI_ISL_3278894, EPI_ISL_3278895, EPI_ISL_3278896, EPI_ISL_3278897, EPI_ISL_3278899, EPI_ISL_3278900, EPI_ISL_3278901, EPI_ISL_3278902, EPI_ISL_3278904, EPI_ISL_3278905, EPI_ISL_3278906, EPI_ISL_3278907,
EPI_ISL_3278908, EPI_ISL_3278910, EPI_ISL_3278912, EPI_ISL_3278913, EPI_ISL_3278914, EPI_ISL_3278915, EPI_ISL_3278916, EPI_ISL_3278917, EPI_ISL_3278918, EPI_ISL_3278919, EPI_ISL_3278920, EPI_ISL_3278921, EPI_ISL_3278922, EPI_ISL_3278923, EPI_ISL_3278925, EPI_ISL_3278927, EPI_ISL_3278928, EPI_ISL_3278929, EPI_ISL_3278930, EPI_ISL_3278931, EPI_ISL_3278932, EPI_ISL_3278933,
EPI_ISL_3278934, EPI_ISL_3278936, EPI_ISL_3278937, EPI_ISL_3278938, EPI_ISL_3278939, EPI_ISL_3278940, EPI_ISL_3278942, EPI_ISL_3278943, EPI_ISL_3278944, EPI_ISL_3278945, EPI_ISL_3278946, EPI_ISL_3278947, EPI_ISL_3278948, EPI_ISL_3278949, EPI_ISL_3278950, EPI_ISL_3278951, EPI_ISL_3278952, EPI_ISL_3278953, EPI_ISL_3278954, EPI_ISL_3278955, EPI_ISL_3278956, EPI_ISL_3278957,
EPI_ISL_3278958, EPI_ISL_3278959, EPI_ISL_3278960, EPI_ISL_3278961, EPI_ISL_3278962, EPI_ISL_3278963, EPI_ISL_3278964, EPI_ISL_3278965, EPI_ISL_3278967, EPI_ISL_3278968, EPI_ISL_3278969, EPI_ISL_3278970, EPI_ISL_3278971, EPI_ISL_3278972, EPI_ISL_3278973, EPI_ISL_3278974, EPI_ISL_3278977, EPI_ISL_3278978, EPI_ISL_3278980, EPI_ISL_3278981, EPI_ISL_3278982, EPI_ISL_3278983,
EPI_ISL_3278984, EPI_ISL_3278985, EPI_ISL_3278986, EPI_ISL_3278987, EPI_ISL_3278988, EPI_ISL_3278989, EPI_ISL_3278990, EPI_ISL_3278991, EPI_ISL_3278992, EPI_ISL_3278993, EPI_ISL_3278994, EPI_ISL_3278995, EPI_ISL_3278996, EPI_ISL_3278997, EPI_ISL_3278998, EPI_ISL_3278999, EPI_ISL_3279000, EPI_ISL_3279001, EPI_ISL_3279002, EPI_ISL_3279003, EPI_ISL_3279004, EPI_ISL_3279005,
EPI_ISL_3279006, EPI_ISL_3279007, EPI_ISL_3279008, EPI_ISL_3279009, EPI_ISL_3279012, EPI_ISL_3279013, EPI_ISL_3279015, EPI_ISL_3279016, EPI_ISL_3279017, EPI_ISL_3279019, EPI_ISL_3279020, EPI_ISL_3279021, EPI_ISL_3279022, EPI_ISL_3279023, EPI_ISL_3279024, EPI_ISL_3279025, EPI_ISL_3279026, EPI_ISL_3279027, EPI_ISL_3279028, EPI_ISL_3279029, EPI_ISL_3279030, EPI_ISL_3279031,
EPI_ISL_3279032, EPI_ISL_3279033, EPI_ISL_3279034, EPI_ISL_3279035, EPI_ISL_3279036, EPI_ISL_3279037, EPI_ISL_3279038, EPI_ISL_3279039, EPI_ISL_3279040, EPI_ISL_3279041, EPI_ISL_3279042, EPI_ISL_3279043, EPI_ISL_3279044, EPI_ISL_3279045, EPI_ISL_3279046, EPI_ISL_3279047, EPI_ISL_3279048, EPI_ISL_3279049, EPI_ISL_3279050, EPI_ISL_3279051, EPI_ISL_3279053, EPI_ISL_3279054,
EPI_ISL_3279055, EPI_ISL_3279057, EPI_ISL_3279058, EPI_ISL_3279060, EPI_ISL_3279061, EPI_ISL_3279062, EPI_ISL_3279063, EPI_ISL_3279064, EPI_ISL_3279065, EPI_ISL_3279066, EPI_ISL_3279067, EPI_ISL_3279068, EPI_ISL_3279069, EPI_ISL_3279070, EPI_ISL_3279071, EPI_ISL_3279072, EPI_ISL_3279073, EPI_ISL_3279074, EPI_ISL_3279075, EPI_ISL_3279076, EPI_ISL_3279077, EPI_ISL_3279078,
EPI_ISL_3279079, EPI_ISL_3279080, EPI_ISL_3279081, EPI_ISL_3279082, EPI_ISL_3279083, EPI_ISL_3279084, EPI_ISL_3279085, EPI_ISL_3279086, EPI_ISL_3279087, EPI_ISL_3279088, EPI_ISL_3279089, EPI_ISL_3279090, EPI_ISL_3279091, EPI_ISL_3279092, EPI_ISL_3279093, EPI_ISL_3279094, EPI_ISL_3279095, EPI_ISL_3279096, EPI_ISL_3279098, EPI_ISL_3279099, EPI_ISL_3279101, EPI_ISL_3279102,
EPI_ISL_3279103, EPI_ISL_3279104, EPI_ISL_3279105, EPI_ISL_3279106, EPI_ISL_3279108, EPI_ISL_3279109, EPI_ISL_3279110, EPI_ISL_3279111, EPI_ISL_3279112, EPI_ISL_3279113, EPI_ISL_3279114, EPI_ISL_3279115, EPI_ISL_3279116, EPI_ISL_3279117, EPI_ISL_3279118, EPI_ISL_3279119, EPI_ISL_3279120, EPI_ISL_3279121, EPI_ISL_3279122, EPI_ISL_3279123, EPI_ISL_3279124, EPI_ISL_3279125,
EPI_ISL_3279126, EPI_ISL_3279127, EPI_ISL_3279128, EPI_ISL_3279129, EPI_ISL_3279130, EPI_ISL_3279131, EPI_ISL_3279132, EPI_ISL_3279133, EPI_ISL_3279134, EPI_ISL_3279135, EPI_ISL_3279136, EPI_ISL_3279137, EPI_ISL_3279138, EPI_ISL_3279139, EPI_ISL_3279140, EPI_ISL_3279141, EPI_ISL_3279142, EPI_ISL_3279145, EPI_ISL_3279146, EPI_ISL_3279148, EPI_ISL_3279149, EPI_ISL_3279150,
EPI_ISL_3279151, EPI_ISL_3279152, EPI_ISL_3279153, EPI_ISL_3279155, EPI_ISL_3279156, EPI_ISL_3279157, EPI_ISL_3279158, EPI_ISL_3279159, EPI_ISL_3279160, EPI_ISL_3279161, EPI_ISL_3279162, EPI_ISL_3279163, EPI_ISL_3279164, EPI_ISL_3279166, EPI_ISL_3279167, EPI_ISL_3279168, EPI_ISL_3279169, EPI_ISL_3279170, EPI_ISL_3279172, EPI_ISL_3279173, EPI_ISL_3279174, EPI_ISL_3279175,
EPI_ISL_3279176, EPI_ISL_3279177, EPI_ISL_3279178, EPI_ISL_3279179, EPI_ISL_3279180, EPI_ISL_3279181, EPI_ISL_3279182, EPI_ISL_3279183, EPI_ISL_3279184, EPI_ISL_3279185, EPI_ISL_3279186, EPI_ISL_3279187, EPI_ISL_3279188, EPI_ISL_3279189, EPI_ISL_3279190, EPI_ISL_3279191, EPI_ISL_3279192, EPI_ISL_3279193, EPI_ISL_3279194, EPI_ISL_3279195, EPI_ISL_3279196, EPI_ISL_3279197,
EPI_ISL_3279198, EPI_ISL_3279200, EPI_ISL_3279201, EPI_ISL_3279202, EPI_ISL_3279203, EPI_ISL_3279204, EPI_ISL_3279205, EPI_ISL_3279206, EPI_ISL_3279207, EPI_ISL_3279208, EPI_ISL_3279210, EPI_ISL_3279212, EPI_ISL_3279216, EPI_ISL_3279217, EPI_ISL_3279218, EPI_ISL_3279220, EPI_ISL_3279221, EPI_ISL_3279224, EPI_ISL_3279225, EPI_ISL_3279226, EPI_ISL_3279227, EPI_ISL_3279228,
EPI_ISL_3279229, EPI_ISL_3279230, EPI_ISL_3279232, EPI_ISL_3279233, EPI_ISL_3279234, EPI_ISL_3279235, EPI_ISL_3279236, EPI_ISL_3279237, EPI_ISL_3279238, EPI_ISL_3279239, EPI_ISL_3279241, EPI_ISL_3279242, EPI_ISL_3279243, EPI_ISL_3279244, EPI_ISL_3279245, EPI_ISL_3279246, EPI_ISL_3279248, EPI_ISL_3279249, EPI_ISL_3279250, EPI_ISL_3279251, EPI_ISL_3279252, EPI_ISL_3279253,
EPI_ISL_3279254, EPI_ISL_3279255, EPI_ISL_3279256, EPI_ISL_3279257, EPI_ISL_3279258, EPI_ISL_3279259, EPI_ISL_3279261, EPI_ISL_3279263, EPI_ISL_3279265, EPI_ISL_3279267, EPI_ISL_3279268, EPI_ISL_3279270, EPI_ISL_3279271, EPI_ISL_3279272, EPI_ISL_3279273, EPI_ISL_3279274, EPI_ISL_3279275, EPI_ISL_3279276, EPI_ISL_3279278, EPI_ISL_3279279, EPI_ISL_3279280, EPI_ISL_3279281,
EPI_ISL_3279282, EPI_ISL_3279283, EPI_ISL_3279284, EPI_ISL_3279285, EPI_ISL_3279286, EPI_ISL_3279287, EPI_ISL_3279288, EPI_ISL_3279289, EPI_ISL_3279290, EPI_ISL_3279291, EPI_ISL_3279292, EPI_ISL_3279293, EPI_ISL_3279294, EPI_ISL_3279295, EPI_ISL_3279296, EPI_ISL_3279297, EPI_ISL_3279298, EPI_ISL_3279300, EPI_ISL_3279301, EPI_ISL_3279302, EPI_ISL_3279303, EPI_ISL_3279305,
EPI_ISL_3279306, EPI_ISL_3279307, EPI_ISL_3279308, EPI_ISL_3279309, EPI_ISL_3279310, EPI_ISL_3279311, EPI_ISL_3279312, EPI_ISL_3279313, EPI_ISL_3279314, EPI_ISL_3279315, EPI_ISL_3279316, EPI_ISL_3279317, EPI_ISL_3279319, EPI_ISL_3279320, EPI_ISL_3279322, EPI_ISL_3279323, EPI_ISL_3279324, EPI_ISL_3279325, EPI_ISL_3279326, EPI_ISL_3279329, EPI_ISL_3279330, EPI_ISL_3279331,
EPI_ISL_3279334, EPI_ISL_3279335, EPI_ISL_3279336, EPI_ISL_3279337, EPI_ISL_3279339, EPI_ISL_3279340, EPI_ISL_3279341, EPI_ISL_3279342, EPI_ISL_3279343, EPI_ISL_3279344, EPI_ISL_3279347, EPI_ISL_3279348, EPI_ISL_3279350, EPI_ISL_3279352, EPI_ISL_3279356, EPI_ISL_3279357, EPI_ISL_3279361, EPI_ISL_3279362, EPI_ISL_3279363, EPI_ISL_3279364, EPI_ISL_3279365, EPI_ISL_3279367,
EPI_ISL_3279368, EPI_ISL_3279370, EPI_ISL_3279371, EPI_ISL_3279372, EPI_ISL_3279373, EPI_ISL_3279374, EPI_ISL_3279375, EPI_ISL_3279376, EPI_ISL_3279377, EPI_ISL_3279378, EPI_ISL_3279379, EPI_ISL_3279380, EPI_ISL_3279381, EPI_ISL_3279383, EPI_ISL_3279385, EPI_ISL_3279386, EPI_ISL_3279388, EPI_ISL_3279389, EPI_ISL_3279390, EPI_ISL_3279391, EPI_ISL_3279393, EPI_ISL_3279394,
EPI_ISL_3279395, EPI_ISL_3279396, EPI_ISL_3279397, EPI_ISL_3279398, EPI_ISL_3279399, EPI_ISL_3279401, EPI_ISL_3279402, EPI_ISL_3279403, EPI_ISL_3279404, EPI_ISL_3279405, EPI_ISL_3279406, EPI_ISL_3279407, EPI_ISL_3279408

| | | | |
|---|---|---|---|
| see above | Israel Central Virology laboratory | Israel National Consortium for SARS-CoV-2 sequencing | Dana Bar-Ilan; Danit Sofer; Efrat Dahan Bucris; Efrat Glick-Saar; Ella Mendelson; Gideon Rechavi; Michal Mandelboim; Miranda Geva; Neta Zuckerman; Netanel Abu; Omer Asraf; Omri Nayshool; Oran Erster; Orna Mor |
| EPI_ISL_4505006 | National Institute of Public Health | State Veterinary Institute Prague | Alexander Nagy; Helena Jirincova; Jaromira Vecerova; Lenka Cernikova; Martina Stara; Timotej Suri |
| EPI_ISL_3115140 | Platform BIS UZA/UAntwerpen | Labo Klinische Biologie, UZA | Basil Britto Xavier; Christine Lammens; Herman Goossens; Ines Verbesselt; Jasmine Coppens; Kathleen Holemans; Marie Le Mercier; Veerle Matheeussen |

EPI_ISL_2545226, EPI_ISL_2545227, EPI_ISL_2545234, EPI_ISL_2545236, EPI_ISL_2545240, EPI_ISL_2545243, EPI_ISL_2545246, EPI_ISL_2545247, EPI_ISL_2545251, EPI_ISL_2545254, EPI_ISL_2545257, EPI_ISL_2545259, EPI_ISL_2545262, EPI_ISL_2545264, EPI_ISL_2545265, EPI_ISL_2545267, EPI_ISL_2545270, EPI_ISL_2545276, EPI_ISL_2545277, EPI_ISL_2545290, EPI_ISL_2545292, EPI_ISL_2545295,
EPI_ISL_2545296, EPI_ISL_2545297, EPI_ISL_2545299, EPI_ISL_2545302, EPI_ISL_2545303, EPI_ISL_2566615, EPI_ISL_2566631, EPI_ISL_2566634, EPI_ISL_2566635, EPI_ISL_2566659, EPI_ISL_2566660, EPI_ISL_2566661, EPI_ISL_2567007, EPI_ISL_2567021, EPI_ISL_2567030, EPI_ISL_2567032, EPI_ISL_2567040, EPI_ISL_2567042, EPI_ISL_2567050, EPI_ISL_2567071, EPI_ISL_2811412, EPI_ISL_2811413,
EPI_ISL_2811414, EPI_ISL_2811415, EPI_ISL_2811416, EPI_ISL_2811417, EPI_ISL_2811418, EPI_ISL_2811419, EPI_ISL_2811420, EPI_ISL_2811421, EPI_ISL_2811422, EPI_ISL_2811423, EPI_ISL_2811424, EPI_ISL_2811425, EPI_ISL_2811426, EPI_ISL_2811427, EPI_ISL_2811428, EPI_ISL_2811429, EPI_ISL_2811430, EPI_ISL_2811431, EPI_ISL_2811432, EPI_ISL_2811433, EPI_ISL_2811434, EPI_ISL_2811435,
EPI_ISL_2811436, EPI_ISL_2811437, EPI_ISL_2811438, EPI_ISL_2811439, EPI_ISL_2811440, EPI_ISL_2811441, EPI_ISL_2811442, EPI_ISL_2811443, EPI_ISL_2811444, EPI_ISL_2811445, EPI_ISL_2811446, EPI_ISL_2811447, EPI_ISL_2811448, EPI_ISL_2811449, EPI_ISL_2811450, EPI_ISL_2811451, EPI_ISL_2811452, EPI_ISL_2811453, EPI_ISL_2811454, EPI_ISL_2811455, EPI_ISL_2811456, EPI_ISL_2811457,
EPI_ISL_2811458, EPI_ISL_2811459, EPI_ISL_2811460, EPI_ISL_2811461, EPI_ISL_2811462, EPI_ISL_2811463, EPI_ISL_2811464, EPI_ISL_2811465, EPI_ISL_2811466, EPI_ISL_2811467, EPI_ISL_2811468, EPI_ISL_2811469, EPI_ISL_2811470, EPI_ISL_2811471, EPI_ISL_2811472, EPI_ISL_2811473, EPI_ISL_2811474, EPI_ISL_2811475, EPI_ISL_2811476, EPI_ISL_2811477, EPI_ISL_2811478, EPI_ISL_2811479,
EPI_ISL_2811480, EPI_ISL_2811481, EPI_ISL_2811482, EPI_ISL_2811483, EPI_ISL_2811484, EPI_ISL_2811485, EPI_ISL_2811486, EPI_ISL_2811487, EPI_ISL_2811488, EPI_ISL_2811489, EPI_ISL_2811490, EPI_ISL_2811491, EPI_ISL_2811492, EPI_ISL_2811493, EPI_ISL_2811494, EPI_ISL_2811495, EPI_ISL_2811496, EPI_ISL_2811497, EPI_ISL_2811498, EPI_ISL_2811499, EPI_ISL_2811500, EPI_ISL_2811501,
EPI_ISL_2811502, EPI_ISL_2811503, EPI_ISL_2811504, EPI_ISL_2811505, EPI_ISL_2811506, EPI_ISL_2811507, EPI_ISL_2811508, EPI_ISL_2811510, EPI_ISL_2811511, EPI_ISL_2811512, EPI_ISL_2811513, EPI_ISL_2811514, EPI_ISL_2811515, EPI_ISL_2811516, EPI_ISL_2811517, EPI_ISL_2811518, EPI_ISL_2811519, EPI_ISL_2811520, EPI_ISL_2811521, EPI_ISL_2811522, EPI_ISL_2811523, EPI_ISL_2811524,
EPI_ISL_2811525, EPI_ISL_2811526, EPI_ISL_2811527, EPI_ISL_2811528, EPI_ISL_2811529, EPI_ISL_2811530, EPI_ISL_2811531, EPI_ISL_2811532, EPI_ISL_2811533, EPI_ISL_2811534, EPI_ISL_2811535, EPI_ISL_2811536, EPI_ISL_2811537, EPI_ISL_2811538, EPI_ISL_2811539, EPI_ISL_2811540, EPI_ISL_2811541, EPI_ISL_2811542, EPI_ISL_2811543, EPI_ISL_2811544, EPI_ISL_2811545, EPI_ISL_2811546,
EPI_ISL_2811547, EPI_ISL_2811548, EPI_ISL_2811549, EPI_ISL_2811550, EPI_ISL_2811551, EPI_ISL_2811552, EPI_ISL_2811553, EPI_ISL_2811554, EPI_ISL_2811555, EPI_ISL_2811556, EPI_ISL_2811558, EPI_ISL_2811559, EPI_ISL_2811560, EPI_ISL_2811561, EPI_ISL_2811562, EPI_ISL_2811563, EPI_ISL_2811564, EPI_ISL_2811565, EPI_ISL_2811566, EPI_ISL_2811567, EPI_ISL_2811568, EPI_ISL_2811569,
EPI_ISL_2811570, EPI_ISL_2811571, EPI_ISL_2811572, EPI_ISL_2811573, EPI_ISL_2811574, EPI_ISL_2811575, EPI_ISL_2811576, EPI_ISL_2811577, EPI_ISL_2811578, EPI_ISL_2811579, EPI_ISL_2811580, EPI_ISL_2811581, EPI_ISL_2811582, EPI_ISL_2811583, EPI_ISL_2811584, EPI_ISL_2811585, EPI_ISL_2811586, EPI_ISL_2811587, EPI_ISL_2811588, EPI_ISL_2811589, EPI_ISL_2811590, EPI_ISL_2811591,
EPI_ISL_2811592, EPI_ISL_2811593, EPI_ISL_2811594, EPI_ISL_2811595, EPI_ISL_2811596, EPI_ISL_2811597, EPI_ISL_2811598, EPI_ISL_2811599, EPI_ISL_2811600, EPI_ISL_2811601, EPI_ISL_2811602, EPI_ISL_2811603, EPI_ISL_2811604, EPI_ISL_2811605, EPI_ISL_2811606, EPI_ISL_2811607, EPI_ISL_2811608, EPI_ISL_2811609, EPI_ISL_2811610, EPI_ISL_2811611, EPI_ISL_2811612, EPI_ISL_2811613,
EPI_ISL_2811614, EPI_ISL_2811615, EPI_ISL_2811616, EPI_ISL_2811617, EPI_ISL_2811618, EPI_ISL_2811619, EPI_ISL_2811620, EPI_ISL_2811621, EPI_ISL_2811622, EPI_ISL_2811623, EPI_ISL_2811624, EPI_ISL_2811625, EPI_ISL_2811626, EPI_ISL_2811627, EPI_ISL_2811628, EPI_ISL_2811629, EPI_ISL_2811630, EPI_ISL_2811631, EPI_ISL_2811632, EPI_ISL_2811633, EPI_ISL_2811634, EPI_ISL_2811635,
EPI_ISL_2811636, EPI_ISL_2811637, EPI_ISL_2811638, EPI_ISL_2811639, EPI_ISL_2811640, EPI_ISL_2811641, EPI_ISL_2811642, EPI_ISL_2811643, EPI_ISL_2811644, EPI_ISL_2811645, EPI_ISL_2811646, EPI_ISL_2811647, EPI_ISL_2811648, EPI_ISL_2811649, EPI_ISL_2811650, EPI_ISL_2811651, EPI_ISL_2811652, EPI_ISL_2811653, EPI_ISL_2811654, EPI_ISL_2811655, EPI_ISL_2811656, EPI_ISL_2811657,
EPI_ISL_2811658, EPI_ISL_2811659, EPI_ISL_2811660, EPI_ISL_2811661, EPI_ISL_2811662, EPI_ISL_2811663, EPI_ISL_2811664, EPI_ISL_2811665, EPI_ISL_2811666, EPI_ISL_2811667, EPI_ISL_2811668, EPI_ISL_2811669, EPI_ISL_2811670, EPI_ISL_2811671, EPI_ISL_2811672, EPI_ISL_2811673, EPI_ISL_2811674, EPI_ISL_2811675, EPI_ISL_2811676, EPI_ISL_2811677, EPI_ISL_2811678, EPI_ISL_2811679,
EPI_ISL_2811680, EPI_ISL_2811681, EPI_ISL_2811682, EPI_ISL_2811683, EPI_ISL_2811684, EPI_ISL_2811685, EPI_ISL_2811686, EPI_ISL_2811687, EPI_ISL_2811688, EPI_ISL_2811689, EPI_ISL_2811690, EPI_ISL_2811691, EPI_ISL_2811692, EPI_ISL_2811693, EPI_ISL_2811694, EPI_ISL_2811695, EPI_ISL_2811696, EPI_ISL_2811697, EPI_ISL_2811698, EPI_ISL_2811699, EPI_ISL_2811700, EPI_ISL_2811701,
EPI_ISL_2811702, EPI_ISL_2811703, EPI_ISL_2811704, EPI_ISL_2811705, EPI_ISL_2811706, EPI_ISL_2811707, EPI_ISL_2811708, EPI_ISL_2811709, EPI_ISL_2811710, EPI_ISL_2811711, EPI_ISL_2811712, EPI_ISL_2811713, EPI_ISL_2811714, EPI_ISL_2811715, EPI_ISL_2834268, EPI_ISL_2834269, EPI_ISL_2834270, EPI_ISL_2834271, EPI_ISL_2834272, EPI_ISL_2834273, EPI_ISL_2834274, EPI_ISL_2834276,
EPI_ISL_2834277, EPI_ISL_2834278, EPI_ISL_2834279, EPI_ISL_2834280, EPI_ISL_2834281, EPI_ISL_2834282, EPI_ISL_2834283, EPI_ISL_2834284, EPI_ISL_2834285, EPI_ISL_2834286, EPI_ISL_2834287, EPI_ISL_2834288, EPI_ISL_2834289, EPI_ISL_2834290, EPI_ISL_2834291, EPI_ISL_2834292, EPI_ISL_2834293, EPI_ISL_2834294, EPI_ISL_2834295, EPI_ISL_2834296, EPI_ISL_2834297, EPI_ISL_2834298,
EPI_ISL_2834299, EPI_ISL_2834300, EPI_ISL_2834301, EPI_ISL_2834302, EPI_ISL_2834303, EPI_ISL_2834304, EPI_ISL_2834305, EPI_ISL_2834306, EPI_ISL_2834307, EPI_ISL_2834308, EPI_ISL_2834309, EPI_ISL_2834310, EPI_ISL_2834311, EPI_ISL_2834312, EPI_ISL_2834313, EPI_ISL_2834314, EPI_ISL_2834315, EPI_ISL_2834316, EPI_ISL_2834317, EPI_ISL_2834318, EPI_ISL_2834319, EPI_ISL_2834320,
EPI_ISL_2834321, EPI_ISL_2834322, EPI_ISL_2834323, EPI_ISL_2834324, EPI_ISL_2834325, EPI_ISL_2834326, EPI_ISL_2834328, EPI_ISL_2834329, EPI_ISL_2834330, EPI_ISL_2834331, EPI_ISL_2834332, EPI_ISL_2834333, EPI_ISL_2834334, EPI_ISL_2834335, EPI_ISL_2834336, EPI_ISL_2834337, EPI_ISL_2834338, EPI_ISL_2834339, EPI_ISL_2834340, EPI_ISL_2834341, EPI_ISL_2834342, EPI_ISL_2834343,
EPI_ISL_2834344, EPI_ISL_2834345, EPI_ISL_2834346, EPI_ISL_2834347, EPI_ISL_2834348, EPI_ISL_2834349, EPI_ISL_2834350, EPI_ISL_2834351, EPI_ISL_2834352, EPI_ISL_2834353, EPI_ISL_2834354, EPI_ISL_2834355, EPI_ISL_2834356, EPI_ISL_2834357, EPI_ISL_2834358, EPI_ISL_2834359, EPI_ISL_2834360, EPI_ISL_2834361, EPI_ISL_2834362, EPI_ISL_2834363, EPI_ISL_2834364, EPI_ISL_2834365,
EPI_ISL_2834366, EPI_ISL_2834367, EPI_ISL_2834368, EPI_ISL_2834369, EPI_ISL_2834371, EPI_ISL_2834372, EPI_ISL_2834373, EPI_ISL_2834374, EPI_ISL_2834375, EPI_ISL_2834376, EPI_ISL_2834377, EPI_ISL_2834378, EPI_ISL_2834379, EPI_ISL_2834380, EPI_ISL_2834381, EPI_ISL_2834382, EPI_ISL_2834383, EPI_ISL_2834384, EPI_ISL_2834385, EPI_ISL_2834386, EPI_ISL_2834387, EPI_ISL_2834389,
EPI_ISL_2834390, EPI_ISL_2834391, EPI_ISL_2834392, EPI_ISL_2834393, EPI_ISL_2834394, EPI_ISL_2834395, EPI_ISL_2834396, EPI_ISL_2834397, EPI_ISL_2834398, EPI_ISL_2834399, EPI_ISL_2834400, EPI_ISL_2834401, EPI_ISL_2834402, EPI_ISL_2834403, EPI_ISL_2834404, EPI_ISL_2834405, EPI_ISL_2834406, EPI_ISL_2834407, EPI_ISL_2834408, EPI_ISL_2834409, EPI_ISL_2834410, EPI_ISL_2834411,
EPI_ISL_2834412, EPI_ISL_2834413, EPI_ISL_2834414, EPI_ISL_2834415, EPI_ISL_2834416, EPI_ISL_2834417, EPI_ISL_2834418, EPI_ISL_2834419, EPI_ISL_2834420, EPI_ISL_2834421, EPI_ISL_2834422, EPI_ISL_2834423, EPI_ISL_2834424, EPI_ISL_2834425, EPI_ISL_2834426, EPI_ISL_2834427, EPI_ISL_2834428, EPI_ISL_2834430, EPI_ISL_2834431, EPI_ISL_2834432, EPI_ISL_2834433, EPI_ISL_2834434,
EPI_ISL_2834435, EPI_ISL_2834436, EPI_ISL_2834437, EPI_ISL_2834438, EPI_ISL_2834439, EPI_ISL_2834440, EPI_ISL_2834441, EPI_ISL_2834442, EPI_ISL_2834443, EPI_ISL_2834444, EPI_ISL_2834445, EPI_ISL_2834446, EPI_ISL_2834447, EPI_ISL_2834448, EPI_ISL_2834449, EPI_ISL_2834450, EPI_ISL_2834451, EPI_ISL_2834452, EPI_ISL_2834453, EPI_ISL_2834455, EPI_ISL_2834456, EPI_ISL_2834457,
EPI_ISL_2834458, EPI_ISL_2834459, EPI_ISL_2834460, EPI_ISL_2834461, EPI_ISL_2834462, EPI_ISL_2834463, EPI_ISL_2834464, EPI_ISL_2834465, EPI_ISL_2834466, EPI_ISL_2834467, EPI_ISL_2834468, EPI_ISL_2834469, EPI_ISL_2834470, EPI_ISL_2834471, EPI_ISL_2834472, EPI_ISL_2834473, EPI_ISL_2834474, EPI_ISL_2834475, EPI_ISL_2834476, EPI_ISL_2834477, EPI_ISL_2834478, EPI_ISL_2834479,
EPI_ISL_2834480, EPI_ISL_2834481, EPI_ISL_2834482, EPI_ISL_2834483, EPI_ISL_2834485, EPI_ISL_2834486, EPI_ISL_2834487, EPI_ISL_2834488, EPI_ISL_2834489, EPI_ISL_2834490, EPI_ISL_2834491, EPI_ISL_2834492, EPI_ISL_2834493, EPI_ISL_2834494, EPI_ISL_2834495, EPI_ISL_2834496, EPI_ISL_2834497, EPI_ISL_2834498, EPI_ISL_2834499, EPI_ISL_2834500, EPI_ISL_2834501, EPI_ISL_2834502,
EPI_ISL_2834503, EPI_ISL_2834504, EPI_ISL_2834506, EPI_ISL_2834507, EPI_ISL_2834508, EPI_ISL_2834509, EPI_ISL_2834510, EPI_ISL_2834511, EPI_ISL_2834512, EPI_ISL_2834513, EPI_ISL_2834514, EPI_ISL_2834515, EPI_ISL_2834516, EPI_ISL_2834517, EPI_ISL_2834518, EPI_ISL_2834519, EPI_ISL_2834520, EPI_ISL_2834522, EPI_ISL_2834523, EPI_ISL_2834524, EPI_ISL_2834525, EPI_ISL_2834526,
EPI_ISL_2834528, EPI_ISL_2834529, EPI_ISL_2834530, EPI_ISL_3019546, EPI_ISL_3019547, EPI_ISL_3019548, EPI_ISL_3019549, EPI_ISL_3019550, EPI_ISL_3019551, EPI_ISL_3019552, EPI_ISL_3019553, EPI_ISL_3019554, EPI_ISL_3019555, EPI_ISL_3019557, EPI_ISL_3019558, EPI_ISL_3019559, EPI_ISL_3019560, EPI_ISL_3019561, EPI_ISL_3019562, EPI_ISL_3019563, EPI_ISL_3019564, EPI_ISL_3019566,
EPI_ISL_3019567, EPI_ISL_3019568, EPI_ISL_3019569, EPI_ISL_3019570, EPI_ISL_3019571, EPI_ISL_3019572, EPI_ISL_3019573, EPI_ISL_3019574, EPI_ISL_3019575, EPI_ISL_3019576, EPI_ISL_3019577, EPI_ISL_3019578, EPI_ISL_3019579, EPI_ISL_3019580, EPI_ISL_3019581, EPI_ISL_3019582, EPI_ISL_3019583, EPI_ISL_3019584, EPI_ISL_3019585, EPI_ISL_3019586, EPI_ISL_3019587, EPI_ISL_3019588,
EPI_ISL_3019589, EPI_ISL_3019590, EPI_ISL_3019591, EPI_ISL_3019592, EPI_ISL_3019593, EPI_ISL_3019594, EPI_ISL_3019595, EPI_ISL_3019596, EPI_ISL_3019597, EPI_ISL_3019598, EPI_ISL_3019599, EPI_ISL_3019600, EPI_ISL_3019601, EPI_ISL_3019602, EPI_ISL_3019603, EPI_ISL_3019604, EPI_ISL_3019605, EPI_ISL_3019606, EPI_ISL_3019607, EPI_ISL_3019608, EPI_ISL_3019609, EPI_ISL_3019610,
EPI_ISL_3019611, EPI_ISL_3019612, EPI_ISL_3019613, EPI_ISL_3019614, EPI_ISL_3019615, EPI_ISL_3019616, EPI_ISL_3019617, EPI_ISL_3019618, EPI_ISL_3019619, EPI_ISL_3019620, EPI_ISL_3019621, EPI_ISL_3019622, EPI_ISL_3019623, EPI_ISL_3019624, EPI_ISL_3019625, EPI_ISL_3019626, EPI_ISL_3019627, EPI_ISL_3019628, EPI_ISL_3019629, EPI_ISL_3019630, EPI_ISL_3019632, EPI_ISL_3019633,
EPI_ISL_3019634, EPI_ISL_3019635, EPI_ISL_3019636, EPI_ISL_3019637, EPI_ISL_3019638, EPI_ISL_3019639, EPI_ISL_3019640, EPI_ISL_3019641, EPI_ISL_3019643, EPI_ISL_3019644, EPI_ISL_3019645, EPI_ISL_3019646, EPI_ISL_3019647, EPI_ISL_3019648, EPI_ISL_3019649, EPI_ISL_3019650, EPI_ISL_3019651, EPI_ISL_3019652, EPI_ISL_3019653, EPI_ISL_3019654, EPI_ISL_3019655, EPI_ISL_3019657,
EPI_ISL_3019658, EPI_ISL_3019659, EPI_ISL_3019660, EPI_ISL_3019661, EPI_ISL_3019662, EPI_ISL_3019663, EPI_ISL_3019664, EPI_ISL_3019665, EPI_ISL_3019666, EPI_ISL_3019667, EPI_ISL_3019668, EPI_ISL_3019669, EPI_ISL_3019670, EPI_ISL_3019671, EPI_ISL_3019672, EPI_ISL_3019673, EPI_ISL_3019674, EPI_ISL_3019675, EPI_ISL_3019676, EPI_ISL_3019677, EPI_ISL_3019678, EPI_ISL_3019679,
EPI_ISL_3019681, EPI_ISL_3019683, EPI_ISL_3019685, EPI_ISL_3019686, EPI_ISL_3019687, EPI_ISL_3019688, EPI_ISL_3019689, EPI_ISL_3019690, EPI_ISL_3019691, EPI_ISL_3019692, EPI_ISL_3019693, EPI_ISL_3019694, EPI_ISL_3019695, EPI_ISL_3019696, EPI_ISL_3019697, EPI_ISL_3019698, EPI_ISL_3019699, EPI_ISL_3019701, EPI_ISL_3019702, EPI_ISL_3019704, EPI_ISL_3019705, EPI_ISL_3019706,
EPI_ISL_3019707, EPI_ISL_3019708, EPI_ISL_3019709, EPI_ISL_3019710, EPI_ISL_3019711, EPI_ISL_3019712, EPI_ISL_3019713, EPI_ISL_3019714, EPI_ISL_3019715, EPI_ISL_3019716, EPI_ISL_3019717, EPI_ISL_3019720, EPI_ISL_3019721, EPI_ISL_3019722, EPI_ISL_3019723, EPI_ISL_3019724, EPI_ISL_3019725, EPI_ISL_3019726, EPI_ISL_3019727, EPI_ISL_3019728, EPI_ISL_3019729, EPI_ISL_3019731,
EPI_ISL_3019732, EPI_ISL_3019733, EPI_ISL_3019734, EPI_ISL_3019735, EPI_ISL_3019736, EPI_ISL_3019737, EPI_ISL_3019738, EPI_ISL_3019739, EPI_ISL_3019740, EPI_ISL_3019741, EPI_ISL_3019742, EPI_ISL_3019743, EPI_ISL_3019744, EPI_ISL_3019745, EPI_ISL_3019746, EPI_ISL_3019747, EPI_ISL_3019748, EPI_ISL_3019749, EPI_ISL_3019750, EPI_ISL_3019751, EPI_ISL_3019752, EPI_ISL_3019753,
EPI_ISL_3019754, EPI_ISL_3019755, EPI_ISL_3019756, EPI_ISL_3019757, EPI_ISL_3019758, EPI_ISL_3019759, EPI_ISL_3019760, EPI_ISL_3019761, EPI_ISL_3019762, EPI_ISL_3019764, EPI_ISL_3019765, EPI_ISL_3019766, EPI_ISL_3019767, EPI_ISL_3019768, EPI_ISL_3019769, EPI_ISL_3019770, EPI_ISL_3019771, EPI_ISL_3019772, EPI_ISL_3019773, EPI_ISL_3019774, EPI_ISL_3019775, EPI_ISL_3019776,
EPI_ISL_3019777, EPI_ISL_3019778, EPI_ISL_3019779, EPI_ISL_3019780, EPI_ISL_3019781, EPI_ISL_3019782, EPI_ISL_3019783, EPI_ISL_3019784, EPI_ISL_3019785, EPI_ISL_3019788, EPI_ISL_3019789, EPI_ISL_3019792, EPI_ISL_3019793, EPI_ISL_3019794, EPI_ISL_3019798, EPI_ISL_3019799, EPI_ISL_3019800, EPI_ISL_3019801, EPI_ISL_3019802, EPI_ISL_3019803, EPI_ISL_3019804, EPI_ISL_3019805,
EPI_ISL_3019806, EPI_ISL_3019807, EPI_ISL_3019808, EPI_ISL_3019809, EPI_ISL_3019810, EPI_ISL_3019812, EPI_ISL_3019813, EPI_ISL_3019814, EPI_ISL_3019815, EPI_ISL_3019816, EPI_ISL_3019817, EPI_ISL_3019818, EPI_ISL_3019819, EPI_ISL_3019820, EPI_ISL_3019821, EPI_ISL_3019822, EPI_ISL_3019823, EPI_ISL_3019824, EPI_ISL_3019825, EPI_ISL_3019826, EPI_ISL_3019827, EPI_ISL_3019828,
EPI_ISL_3019829, EPI_ISL_3019832, EPI_ISL_3019833, EPI_ISL_3019834, EPI_ISL_3019835, EPI_ISL_3019836, EPI_ISL_3019837, EPI_ISL_3019838, EPI_ISL_3019839, EPI_ISL_3019840, EPI_ISL_3019842, EPI_ISL_3019843, EPI_ISL_3019844, EPI_ISL_3019845, EPI_ISL_3019846, EPI_ISL_3019847, EPI_ISL_3019848, EPI_ISL_3019849, EPI_ISL_3019850, EPI_ISL_3019851, EPI_ISL_3019852, EPI_ISL_3019853,
EPI_ISL_3019854, EPI_ISL_3019856, EPI_ISL_3019857, EPI_ISL_3019858, EPI_ISL_3019859, EPI_ISL_3019860, EPI_ISL_3019861, EPI_ISL_3019862, EPI_ISL_3019863, EPI_ISL_3019864, EPI_ISL_3019865, EPI_ISL_3019867, EPI_ISL_3019868, EPI_ISL_3019869, EPI_ISL_3019870, EPI_ISL_3019871, EPI_ISL_3019872, EPI_ISL_3019873, EPI_ISL_3019874, EPI_ISL_3019876, EPI_ISL_3019877, EPI_ISL_3019878,
EPI_ISL_3019879, EPI_ISL_3019880, EPI_ISL_3019881, EPI_ISL_3019882, EPI_ISL_3019883, EPI_ISL_3019884, EPI_ISL_3019885, EPI_ISL_3019886, EPI_ISL_3019887, EPI_ISL_3019889, EPI_ISL_3019890, EPI_ISL_3019891, EPI_ISL_3019892, EPI_ISL_3019893, EPI_ISL_3019894, EPI_ISL_3019895, EPI_ISL_3019896, EPI_ISL_3019897, EPI_ISL_3019898, EPI_ISL_3019899, EPI_ISL_3019900, EPI_ISL_3019901,
EPI_ISL_3019902, EPI_ISL_3019904, EPI_ISL_3019905, EPI_ISL_3019906, EPI_ISL_3019907, EPI_ISL_3019908, EPI_ISL_3019909, EPI_ISL_3019910, EPI_ISL_3019911, EPI_ISL_3021549, EPI_ISL_3021550, EPI_ISL_3021551, EPI_ISL_3021552, EPI_ISL_3021553, EPI_ISL_3021554, EPI_ISL_3021555, EPI_ISL_3021556, EPI_ISL_3021557, EPI_ISL_3021559, EPI_ISL_3021560, EPI_ISL_3021561, EPI_ISL_3021562,
EPI_ISL_3021563, EPI_ISL_3021564, EPI_ISL_3021565, EPI_ISL_3021566, EPI_ISL_3021567, EPI_ISL_3021568, EPI_ISL_3021570, EPI_ISL_3021571, EPI_ISL_3021572, EPI_ISL_3021573, EPI_ISL_3021574, EPI_ISL_3021575, EPI_ISL_3021576, EPI_ISL_3021577, EPI_ISL_3021578, EPI_ISL_3021579, EPI_ISL_3021581, EPI_ISL_3021583, EPI_ISL_3021584, EPI_ISL_3021585, EPI_ISL_3021587, EPI_ISL_3021588,
EPI_ISL_3021589, EPI_ISL_3021590, EPI_ISL_3021591, EPI_ISL_3021592, EPI_ISL_3021593, EPI_ISL_3021594, EPI_ISL_3021595, EPI_ISL_3021597, EPI_ISL_3021599, EPI_ISL_3021600, EPI_ISL_3021601, EPI_ISL_3021602, EPI_ISL_3021603, EPI_ISL_3021604, EPI_ISL_3021605, EPI_ISL_3021606, EPI_ISL_3021607, EPI_ISL_3021608, EPI_ISL_3021609, EPI_ISL_3021610, EPI_ISL_3021611, EPI_ISL_3021612,
EPI_ISL_3021613, EPI_ISL_3021614, EPI_ISL_3021615, EPI_ISL_3021617, EPI_ISL_3021618, EPI_ISL_3021619, EPI_ISL_3021620, EPI_ISL_3021621, EPI_ISL_3021622, EPI_ISL_3021623, EPI_ISL_3021624, EPI_ISL_3021625, EPI_ISL_3021626, EPI_ISL_3021627, EPI_ISL_3021628, EPI_ISL_3021629, EPI_ISL_3021630, EPI_ISL_3021631, EPI_ISL_3021632, EPI_ISL_3021633, EPI_ISL_3021634, EPI_ISL_3021635,
EPI_ISL_3021636, EPI_ISL_3021637, EPI_ISL_3021638, EPI_ISL_3021641, EPI_ISL_3021644, EPI_ISL_3021645, EPI_ISL_3021647, EPI_ISL_3021648, EPI_ISL_3021649, EPI_ISL_3021650, EPI_ISL_3021651, EPI_ISL_3021652, EPI_ISL_3021653, EPI_ISL_3021654, EPI_ISL_3021657, EPI_ISL_3021659, EPI_ISL_3021661, EPI_ISL_3021662, EPI_ISL_3021663, EPI_ISL_3021664, EPI_ISL_3021667, EPI_ISL_3021668,
EPI_ISL_3021669, EPI_ISL_3021670, EPI_ISL_3021671, EPI_ISL_3021672, EPI_ISL_3021673, EPI_ISL_3021674, EPI_ISL_3021675, EPI_ISL_3021676, EPI_ISL_3021677, EPI_ISL_3021678, EPI_ISL_3021679, EPI_ISL_3021680, EPI_ISL_3021681, EPI_ISL_3021682, EPI_ISL_3021683, EPI_ISL_3021684, EPI_ISL_3021685, EPI_ISL_3021686, EPI_ISL_3021688, EPI_ISL_3021689, EPI_ISL_3021690, EPI_ISL_3021691,
EPI_ISL_3021692, EPI_ISL_3021693, EPI_ISL_3021694, EPI_ISL_3021695, EPI_ISL_3021696, EPI_ISL_3021697, EPI_ISL_3021698, EPI_ISL_3021699, EPI_ISL_3021700, EPI_ISL_3021701, EPI_ISL_3021703, EPI_ISL_3021704, EPI_ISL_3021705, EPI_ISL_3021708, EPI_ISL_3021709, EPI_ISL_3021710, EPI_ISL_3021711, EPI_ISL_3021712, EPI_ISL_3021713, EPI_ISL_3021714, EPI_ISL_3021716, EPI_ISL_3021717,
EPI_ISL_3021718, EPI_ISL_3021719, EPI_ISL_3021720, EPI_ISL_3021721, EPI_ISL_3021722, EPI_ISL_3021723, EPI_ISL_3021724, EPI_ISL_3021725, EPI_ISL_3021726, EPI_ISL_3021728, EPI_ISL_3021729, EPI_ISL_3021731, EPI_ISL_3021732, EPI_ISL_3021733, EPI_ISL_3021734, EPI_ISL_3021735, EPI_ISL_3021736, EPI_ISL_3021737, EPI_ISL_3021738, EPI_ISL_3021739, EPI_ISL_3021740, EPI_ISL_3021741,
EPI_ISL_3021742, EPI_ISL_3021744, EPI_ISL_3021745, EPI_ISL_3021746, EPI_ISL_3021747, EPI_ISL_3021748, EPI_ISL_3021749, EPI_ISL_3021750, EPI_ISL_3021751, EPI_ISL_3021752, EPI_ISL_3021753, EPI_ISL_3021754, EPI_ISL_3021755, EPI_ISL_3021756, EPI_ISL_3021757, EPI_ISL_3021758, EPI_ISL_3021759, EPI_ISL_3021760, EPI_ISL_3021761, EPI_ISL_3021762, EPI_ISL_3021763, EPI_ISL_3021764,
EPI_ISL_3021766, EPI_ISL_3021767, EPI_ISL_3021768, EPI_ISL_3021769, EPI_ISL_3021770, EPI_ISL_3021771, EPI_ISL_3021772, EPI_ISL_3021773, EPI_ISL_3021774, EPI_ISL_3021775, EPI_ISL_3021776, EPI_ISL_3021777, EPI_ISL_3021778, EPI_ISL_3021779, EPI_ISL_3021780, EPI_ISL_3021781, EPI_ISL_3021782, EPI_ISL_3021783, EPI_ISL_3021784, EPI_ISL_3021785, EPI_ISL_3021786, EPI_ISL_3021787,
EPI_ISL_3021788, EPI_ISL_3021789, EPI_ISL_3021790, EPI_ISL_3021792, EPI_ISL_3021793, EPI_ISL_3021794, EPI_ISL_3021795, EPI_ISL_3021796, EPI_ISL_3021797, EPI_ISL_3021798, EPI_ISL_3021799, EPI_ISL_3021800, EPI_ISL_3021801, EPI_ISL_3021802, EPI_ISL_3021803, EPI_ISL_3021804, EPI_ISL_3021805, EPI_ISL_3021806, EPI_ISL_3021807, EPI_ISL_3021808, EPI_ISL_3021809, EPI_ISL_3021811,
EPI_ISL_3021812, EPI_ISL_3021813, EPI_ISL_3021814, EPI_ISL_3021815, EPI_ISL_3021817, EPI_ISL_3021818, EPI_ISL_3021819, EPI_ISL_3021820, EPI_ISL_3021821, EPI_ISL_3021822, EPI_ISL_3021823, EPI_ISL_3021824, EPI_ISL_3021825, EPI_ISL_3021826, EPI_ISL_3021827, EPI_ISL_3021830, EPI_ISL_3021831, EPI_ISL_3021832, EPI_ISL_3021833, EPI_ISL_3021834, EPI_ISL_3021835, EPI_ISL_3021836,
EPI_ISL_3021837, EPI_ISL_3021838, EPI_ISL_3021839, EPI_ISL_3021840, EPI_ISL_3021841, EPI_ISL_3021842, EPI_ISL_3021843, EPI_ISL_3021844, EPI_ISL_3021845, EPI_ISL_3021846, EPI_ISL_3021847, EPI_ISL_3021848, EPI_ISL_3021849, EPI_ISL_3021851, EPI_ISL_3021852, EPI_ISL_3021853, EPI_ISL_3021854, EPI_ISL_3021855, EPI_ISL_3021856, EPI_ISL_3021858, EPI_ISL_3021859, EPI_ISL_3021860,
EPI_ISL_3021861, EPI_ISL_3021862, EPI_ISL_3021863, EPI_ISL_3021864, EPI_ISL_3021865, EPI_ISL_3021866, EPI_ISL_3021867, EPI_ISL_3021869, EPI_ISL_3021871, EPI_ISL_3021872, EPI_ISL_3021873, EPI_ISL_3021874, EPI_ISL_3021875, EPI_ISL_3021876, EPI_ISL_3021877, EPI_ISL_3021878, EPI_ISL_3021879, EPI_ISL_3021880, EPI_ISL_3021881, EPI_ISL_3021882, EPI_ISL_3021883, EPI_ISL_3022014,
EPI_ISL_3022015, EPI_ISL_3022016, EPI_ISL_3022018, EPI_ISL_3022019, EPI_ISL_3022020, EPI_ISL_3022021, EPI_ISL_3022022, EPI_ISL_3022023, EPI_ISL_3022024, EPI_ISL_3022025, EPI_ISL_3022026, EPI_ISL_3022027, EPI_ISL_3022028, EPI_ISL_3022029, EPI_ISL_3022030, EPI_ISL_3022031, EPI_ISL_3022032, EPI_ISL_3022033, EPI_ISL_3022034, EPI_ISL_3022035, EPI_ISL_3022037, EPI_ISL_3022038,
EPI_ISL_3022039, EPI_ISL_3022040, EPI_ISL_3022041, EPI_ISL_3022042, EPI_ISL_3022043, EPI_ISL_3022044, EPI_ISL_3022045, EPI_ISL_3022046, EPI_ISL_3022047, EPI_ISL_3022048, EPI_ISL_3022049, EPI_ISL_3022050, EPI_ISL_3022051, EPI_ISL_3022052, EPI_ISL_3022053, EPI_ISL_3022054, EPI_ISL_3022055, EPI_ISL_3022056, EPI_ISL_3022057, EPI_ISL_3022058, EPI_ISL_3022059, EPI_ISL_3022060,
EPI_ISL_3022061, EPI_ISL_3022062, EPI_ISL_3022063, EPI_ISL_3022064, EPI_ISL_3022065, EPI_ISL_3022066, EPI_ISL_3022067, EPI_ISL_3022068, EPI_ISL_3022069, EPI_ISL_3022070, EPI_ISL_3022071, EPI_ISL_3022072, EPI_ISL_3022073, EPI_ISL_3022074, EPI_ISL_3022075, EPI_ISL_3022076, EPI_ISL_3022077, EPI_ISL_3022078, EPI_ISL_3022079, EPI_ISL_3022080, EPI_ISL_3022081, EPI_ISL_3022082,
EPI_ISL_3022083, EPI_ISL_3022084, EPI_ISL_3022085, EPI_ISL_3022086, EPI_ISL_3022087, EPI_ISL_3022088, EPI_ISL_3022089, EPI_ISL_3022090, EPI_ISL_3022091, EPI_ISL_3022092, EPI_ISL_3022093, EPI_ISL_3022094, EPI_ISL_3022095, EPI_ISL_3022096, EPI_ISL_3022098, EPI_ISL_3022099, EPI_ISL_3022100, EPI_ISL_3022101, EPI_ISL_3022102, EPI_ISL_3022103, EPI_ISL_3022104, EPI_ISL_3022105,
EPI_ISL_3022106, EPI_ISL_3022107, EPI_ISL_3022108, EPI_ISL_3022109, EPI_ISL_3022110, EPI_ISL_3022111, EPI_ISL_3022112, EPI_ISL_3022113, EPI_ISL_3022114, EPI_ISL_3022115, EPI_ISL_3022116, EPI_ISL_3022117, EPI_ISL_3022118, EPI_ISL_3022120, EPI_ISL_3022121, EPI_ISL_3022122, EPI_ISL_3022123, EPI_ISL_3022124, EPI_ISL_3022125, EPI_ISL_3022126, EPI_ISL_3022127, EPI_ISL_3022128,
EPI_ISL_3022129, EPI_ISL_3022130, EPI_ISL_3022131, EPI_ISL_3022132, EPI_ISL_3022133, EPI_ISL_3022134, EPI_ISL_3022135, EPI_ISL_3022137, EPI_ISL_3022138, EPI_ISL_3022139, EPI_ISL_3022140, EPI_ISL_3022141, EPI_ISL_3022142, EPI_ISL_3022143, EPI_ISL_3022144, EPI_ISL_3022145, EPI_ISL_3022146, EPI_ISL_3022147, EPI_ISL_3022148, EPI_ISL_3022149, EPI_ISL_3022150, EPI_ISL_3022151,
EPI_ISL_3022152, EPI_ISL_3022153, EPI_ISL_3022155, EPI_ISL_3022156, EPI_ISL_3022158, EPI_ISL_3022160, EPI_ISL_3022161, EPI_ISL_3022162, EPI_ISL_3022163, EPI_ISL_3022164, EPI_ISL_3022165, EPI_ISL_3022166, EPI_ISL_3022168, EPI_ISL_3022169, EPI_ISL_3022171, EPI_ISL_3022172, EPI_ISL_3022173, EPI_ISL_3022174, EPI_ISL_3022175, EPI_ISL_3022176, EPI_ISL_3022177, EPI_ISL_3022178,
EPI_ISL_3022179, EPI_ISL_3022180, EPI_ISL_3022181, EPI_ISL_3022182, EPI_ISL_3022183, EPI_ISL_3022184, EPI_ISL_3022185, EPI_ISL_3022186, EPI_ISL_3022187, EPI_ISL_3022189, EPI_ISL_3022191, EPI_ISL_3022192, EPI_ISL_3022193, EPI_ISL_3022194, EPI_ISL_3022195, EPI_ISL_3022196, EPI_ISL_3022197, EPI_ISL_3022198, EPI_ISL_3022200, EPI_ISL_3022201, EPI_ISL_3022202, EPI_ISL_3022203,
EPI_ISL_3022205, EPI_ISL_3022206, EPI_ISL_3022208, EPI_ISL_3022209, EPI_ISL_3022210, EPI_ISL_3022211, EPI_ISL_3022212, EPI_ISL_3022213, EPI_ISL_3022214, EPI_ISL_3022216, EPI_ISL_3022217, EPI_ISL_3022218, EPI_ISL_3022219, EPI_ISL_3022220, EPI_ISL_3022221, EPI_ISL_3022223, EPI_ISL_3022224, EPI_ISL_3022225, EPI_ISL_3022226, EPI_ISL_3022227, EPI_ISL_3022228, EPI_ISL_3022229,
EPI_ISL_3022231, EPI_ISL_3022232, EPI_ISL_3022233, EPI_ISL_3022234, EPI_ISL_3022236, EPI_ISL_3022237, EPI_ISL_3022238, EPI_ISL_3022239, EPI_ISL_3022240, EPI_ISL_3022241, EPI_ISL_3022242, EPI_ISL_3022243, EPI_ISL_3022245, EPI_ISL_3022246, EPI_ISL_3022247, EPI_ISL_3022249, EPI_ISL_3022252, EPI_ISL_3022253, EPI_ISL_3022254, EPI_ISL_3022255, EPI_ISL_3022257, EPI_ISL_3022258,
EPI_ISL_3022259, EPI_ISL_3022260, EPI_ISL_3022261, EPI_ISL_3022262, EPI_ISL_3022263, EPI_ISL_3022264, EPI_ISL_3022266, EPI_ISL_3022267, EPI_ISL_3022268, EPI_ISL_3022269, EPI_ISL_3022271, EPI_ISL_3022273, EPI_ISL_3022274, EPI_ISL_3022275, EPI_ISL_3022276, EPI_ISL_3022277, EPI_ISL_3022279, EPI_ISL_3022280, EPI_ISL_3022281, EPI_ISL_3022282, EPI_ISL_3022283, EPI_ISL_3022284,
EPI_ISL_3022285, EPI_ISL_3022286, EPI_ISL_3022288, EPI_ISL_3022289, EPI_ISL_3022290, EPI_ISL_3022291, EPI_ISL_3022292, EPI_ISL_3022293, EPI_ISL_3022294, EPI_ISL_3022295, EPI_ISL_3022296, EPI_ISL_3022297, EPI_ISL_3022298, EPI_ISL_3022299, EPI_ISL_3022300, EPI_ISL_3022301, EPI_ISL_3022302, EPI_ISL_3022303, EPI_ISL_3022304, EPI_ISL_3022305, EPI_ISL_3022306, EPI_ISL_3022307,
EPI_ISL_3022308, EPI_ISL_3022309, EPI_ISL_3022310, EPI_ISL_3022311, EPI_ISL_3022312, EPI_ISL_3022313, EPI_ISL_3022314, EPI_ISL_3022315, EPI_ISL_3022316, EPI_ISL_3022317, EPI_ISL_3022318, EPI_ISL_3022319, EPI_ISL_3022321, EPI_ISL_3022322, EPI_ISL_3022323, EPI_ISL_3022324, EPI_ISL_3022325, EPI_ISL_3022326, EPI_ISL_3022327, EPI_ISL_3022329, EPI_ISL_3022330, EPI_ISL_3022331,
EPI_ISL_3022332, EPI_ISL_3022334, EPI_ISL_3022335, EPI_ISL_3022336, EPI_ISL_3022337, EPI_ISL_3022338, EPI_ISL_3022339, EPI_ISL_3022340, EPI_ISL_3022341, EPI_ISL_3022342, EPI_ISL_3022343, EPI_ISL_3022345, EPI_ISL_3022346, EPI_ISL_3022347, EPI_ISL_3022348, EPI_ISL_3022349, EPI_ISL_3022350, EPI_ISL_3022351, EPI_ISL_3022352, EPI_ISL_3022353, EPI_ISL_3022354, EPI_ISL_3022355,
EPI_ISL_3022356, EPI_ISL_3024135, EPI_ISL_3024136, EPI_ISL_3024137, EPI_ISL_3024138, EPI_ISL_3024139, EPI_ISL_3024140, EPI_ISL_3024141, EPI_ISL_3024142, EPI_ISL_3024143, EPI_ISL_3024144, EPI_ISL_3024146, EPI_ISL_3024147, EPI_ISL_3024148, EPI_ISL_3024149, EPI_ISL_3024150, EPI_ISL_3024151, EPI_ISL_3024152, EPI_ISL_3024153, EPI_ISL_3024154, EPI_ISL_3024155, EPI_ISL_3024156,
EPI_ISL_3024157, EPI_ISL_3024158, EPI_ISL_3024159, EPI_ISL_3024161, EPI_ISL_3024162, EPI_ISL_3024163, EPI_ISL_3024164, EPI_ISL_3024165, EPI_ISL_3024166, EPI_ISL_3024167, EPI_ISL_3024168, EPI_ISL_3024169, EPI_ISL_3024170, EPI_ISL_3024171, EPI_ISL_3024172, EPI_ISL_3024173, EPI_ISL_3024174, EPI_ISL_3024175, EPI_ISL_3024176, EPI_ISL_3024177, EPI_ISL_3024178, EPI_ISL_3024179,
EPI_ISL_3024180, EPI_ISL_3024181, EPI_ISL_3024182, EPI_ISL_3024183, EPI_ISL_3024184, EPI_ISL_3024185, EPI_ISL_3024186, EPI_ISL_3024187, EPI_ISL_3024188, EPI_ISL_3024189, EPI_ISL_3024190, EPI_ISL_3024191, EPI_ISL_3024193, EPI_ISL_3024195, EPI_ISL_3024197, EPI_ISL_3024198, EPI_ISL_3024199, EPI_ISL_3024200, EPI_ISL_3024201, EPI_ISL_3024202, EPI_ISL_3024203, EPI_ISL_3024204,
EPI_ISL_3024205, EPI_ISL_3024206, EPI_ISL_3024207, EPI_ISL_3024208, EPI_ISL_3024209, EPI_ISL_3024210, EPI_ISL_3024211, EPI_ISL_3024212, EPI_ISL_3024213, EPI_ISL_3024214, EPI_ISL_3024215, EPI_ISL_3024216, EPI_ISL_3024217, EPI_ISL_3024219, EPI_ISL_3024221, EPI_ISL_3024222, EPI_ISL_3024224, EPI_ISL_3024225, EPI_ISL_3024226, EPI_ISL_3024227, EPI_ISL_3024228, EPI_ISL_3024229,
EPI_ISL_3024231, EPI_ISL_3024232, EPI_ISL_3024233, EPI_ISL_3024234, EPI_ISL_3024235, EPI_ISL_3024236, EPI_ISL_3024237, EPI_ISL_3024238, EPI_ISL_3024239, EPI_ISL_3024243, EPI_ISL_3024244, EPI_ISL_3024245, EPI_ISL_3024246, EPI_ISL_3024247, EPI_ISL_3024248, EPI_ISL_3024249, EPI_ISL_3024250, EPI_ISL_3024251, EPI_ISL_3024252, EPI_ISL_3024253, EPI_ISL_3024254, EPI_ISL_3024255,
EPI_ISL_3024257, EPI_ISL_3024258, EPI_ISL_3024259, EPI_ISL_3024260, EPI_ISL_3024261, EPI_ISL_3024262, EPI_ISL_3024263, EPI_ISL_3024264, EPI_ISL_3024265, EPI_ISL_3024266, EPI_ISL_3024267, EPI_ISL_3024268, EPI_ISL_3024269, EPI_ISL_3024270, EPI_ISL_3024271, EPI_ISL_3024272, EPI_ISL_3024273, EPI_ISL_3024275, EPI_ISL_3024276, EPI_ISL_3024278, EPI_ISL_3024279, EPI_ISL_3024280,
EPI_ISL_3024281, EPI_ISL_3024282, EPI_ISL_3024283, EPI_ISL_3024284, EPI_ISL_3024285, EPI_ISL_3024286, EPI_ISL_3024287, EPI_ISL_3024288, EPI_ISL_3024289, EPI_ISL_3024290, EPI_ISL_3024291, EPI_ISL_3024292, EPI_ISL_3024293, EPI_ISL_3024294, EPI_ISL_3024295, EPI_ISL_3024296, EPI_ISL_3024297, EPI_ISL_3024298, EPI_ISL_3024299, EPI_ISL_3024300, EPI_ISL_3024301, EPI_ISL_3024302,
EPI_ISL_3024303, EPI_ISL_3024304, EPI_ISL_3024305, EPI_ISL_3024306, EPI_ISL_3024307, EPI_ISL_3024308, EPI_ISL_3024309, EPI_ISL_3024310, EPI_ISL_3024312, EPI_ISL_3024313, EPI_ISL_3024314, EPI_ISL_3024315, EPI_ISL_3024317, EPI_ISL_3024318, EPI_ISL_3024319, EPI_ISL_3024320, EPI_ISL_3024321, EPI_ISL_3024323, EPI_ISL_3024324, EPI_ISL_3024325, EPI_ISL_3024326, EPI_ISL_3024329,
EPI_ISL_3024330, EPI_ISL_3024331, EPI_ISL_3024332, EPI_ISL_3024333, EPI_ISL_3024334, EPI_ISL_3024335, EPI_ISL_3024336, EPI_ISL_3024337, EPI_ISL_3024338, EPI_ISL_3024339, EPI_ISL_3024340, EPI_ISL_3024341, EPI_ISL_3024342, EPI_ISL_3024343, EPI_ISL_3024344, EPI_ISL_3024345, EPI_ISL_3024346, EPI_ISL_3024347, EPI_ISL_3024349, EPI_ISL_3024350, EPI_ISL_3024351, EPI_ISL_3024352,
EPI_ISL_3024354, EPI_ISL_3024355, EPI_ISL_3024356, EPI_ISL_3024357, EPI_ISL_3024358, EPI_ISL_3024359, EPI_ISL_3024360, EPI_ISL_3024361, EPI_ISL_3024362, EPI_ISL_3024363, EPI_ISL_3024364, EPI_ISL_3024365, EPI_ISL_3024366, EPI_ISL_3024368, EPI_ISL_3024370, EPI_ISL_3024371, EPI_ISL_3024372, EPI_ISL_3024373, EPI_ISL_3024374, EPI_ISL_3024375, EPI_ISL_3024376, EPI_ISL_3024377,
EPI_ISL_3024378, EPI_ISL_3024381, EPI_ISL_3024382, EPI_ISL_3024383, EPI_ISL_3024384, EPI_ISL_3024385, EPI_ISL_3024386, EPI_ISL_3024387, EPI_ISL_3024388, EPI_ISL_3024389, EPI_ISL_3024390, EPI_ISL_3024391, EPI_ISL_3024392, EPI_ISL_3024393, EPI_ISL_3024394, EPI_ISL_3024395, EPI_ISL_3024396, EPI_ISL_3024397, EPI_ISL_3024398, EPI_ISL_3024399, EPI_ISL_3024400, EPI_ISL_3024401,
EPI_ISL_3024402, EPI_ISL_3024403, EPI_ISL_3024404, EPI_ISL_3024405, EPI_ISL_3024406, EPI_ISL_3024408, EPI_ISL_3024409, EPI_ISL_3024410, EPI_ISL_3024411, EPI_ISL_3024412, EPI_ISL_3024413, EPI_ISL_3024414, EPI_ISL_3024415, EPI_ISL_3024416, EPI_ISL_3024417, EPI_ISL_3024418, EPI_ISL_3024420, EPI_ISL_3024421, EPI_ISL_3024422, EPI_ISL_3024423, EPI_ISL_3024424, EPI_ISL_3024425,
EPI_ISL_3024426, EPI_ISL_3024427, EPI_ISL_3024428, EPI_ISL_3024429, EPI_ISL_3024430, EPI_ISL_3024431, EPI_ISL_3024432, EPI_ISL_3024433, EPI_ISL_3024434, EPI_ISL_3024435, EPI_ISL_3024436, EPI_ISL_3024437, EPI_ISL_3024438, EPI_ISL_3024439, EPI_ISL_3024440, EPI_ISL_3024441, EPI_ISL_3024442, EPI_ISL_3024443, EPI_ISL_3024444, EPI_ISL_3024445, EPI_ISL_3024446, EPI_ISL_3024449,

EPI_ISL_3024450, EPI_ISL_3024451, EPI_ISL_3024452, EPI_ISL_3024453, EPI_ISL_3024454, EPI_ISL_3024455, EPI_ISL_3024456, EPI_ISL_3024457, EPI_ISL_3024458, EPI_ISL_3024459, EPI_ISL_3024460, EPI_ISL_3024461, EPI_ISL_3024462, EPI_ISL_3024463, EPI_ISL_3024464, EPI_ISL_3024465, EPI_ISL_3024466, EPI_ISL_3024467, EPI_ISL_3024468, EPI_ISL_3024469, EPI_ISL_3024472, EPI_ISL_3024473, EPI_ISL_3024474, EPI_ISL_3024475, EPI_ISL_3024477, EPI_ISL_3024478, EPI_ISL_3024479, EPI_ISL_3024480, EPI_ISL_3024481, EPI_ISL_3024483, EPI_ISL_3024484, EPI_ISL_3024485, EPI_ISL_3024486, EPI_ISL_3024487, EPI_ISL_3024488, EPI_ISL_3024489, EPI_ISL_3024490, EPI_ISL_3024491, EPI_ISL_3024492, EPI_ISL_3024493, EPI_ISL_3024494, EPI_ISL_3024496, EPI_ISL_3024498, EPI_ISL_3667297, EPI_ISL_3667298, EPI_ISL_3667299, EPI_ISL_3667300, EPI_ISL_3667301, EPI_ISL_3667302, EPI_ISL_3667303, EPI_ISL_3667304, EPI_ISL_3667305, EPI_ISL_3667306, EPI_ISL_3667307, EPI_ISL_3667308, EPI_ISL_3667309, EPI_ISL_3667310, EPI_ISL_3667311, EPI_ISL_3667312, EPI_ISL_3667313, EPI_ISL_3667314, EPI_ISL_3667315, EPI_ISL_3667316, EPI_ISL_3667317, EPI_ISL_3667318, EPI_ISL_3667319, EPI_ISL_3667320, EPI_ISL_3667321, EPI_ISL_3667322, EPI_ISL_3667323, EPI_ISL_3667324, EPI_ISL_3667325, EPI_ISL_3667326, EPI_ISL_3667327, EPI_ISL_3667328, EPI_ISL_3667329, EPI_ISL_3667330, EPI_ISL_3667331, EPI_ISL_3667332, EPI_ISL_3667333, EPI_ISL_3667334, EPI_ISL_3667335, EPI_ISL_3667336, EPI_ISL_3667337, EPI_ISL_3667338, EPI_ISL_3667339, EPI_ISL_3667340, EPI_ISL_3667341, EPI_ISL_3667342, EPI_ISL_3667343, EPI_ISL_3667344, EPI_ISL_3667345, EPI_ISL_3667346, EPI_ISL_3667347, EPI_ISL_3667348, EPI_ISL_3667349, EPI_ISL_3667350, EPI_ISL_3667351, EPI_ISL_3667352, EPI_ISL_3667353, EPI_ISL_3667354, EPI_ISL_3667355, EPI_ISL_3667356, EPI_ISL_3667357, EPI_ISL_3667358, EPI_ISL_3667359, EPI_ISL_3667360, EPI_ISL_3667361, EPI_ISL_3667362, EPI_ISL_3667363, EPI_ISL_3667364, EPI_ISL_3667365, EPI_ISL_3667366, EPI_ISL_3667367, EPI_ISL_3667368, EPI_ISL_3667369, EPI_ISL_3667370, EPI_ISL_3667371, EPI_ISL_3667372, EPI_ISL_3667373, EPI_ISL_3667374, EPI_ISL_3667375, EPI_ISL_3667376, EPI_ISL_3667377, EPI_ISL_3667378, EPI_ISL_3667379, EPI_ISL_3667380, EPI_ISL_3667381, EPI_ISL_3667382, EPI_ISL_3667383, EPI_ISL_3667384, EPI_ISL_3667385, EPI_ISL_3667386, EPI_ISL_3667387, EPI_ISL_3667388, EPI_ISL_3667389, EPI_ISL_3667390, EPI_ISL_3667391, EPI_ISL_3667392, EPI_ISL_3667393, EPI_ISL_3667394, EPI_ISL_3667395, EPI_ISL_3667396, EPI_ISL_3667397, EPI_ISL_3667398, EPI_ISL_3667399, EPI_ISL_3667400, EPI_ISL_3667401, EPI_ISL_3667402, EPI_ISL_3667403, EPI_ISL_3667404, EPI_ISL_3667405, EPI_ISL_3667406, EPI_ISL_3667407, EPI_ISL_3667408, EPI_ISL_3667409, EPI_ISL_3667410, EPI_ISL_3667411, EPI_ISL_3667412, EPI_ISL_3667413, EPI_ISL_3667414, EPI_ISL_3667415, EPI_ISL_3667416, EPI_ISL_3667417, EPI_ISL_3667418, EPI_ISL_3667419, EPI_ISL_3667420, EPI_ISL_3667421, EPI_ISL_3667422, EPI_ISL_3667423, EPI_ISL_3667424, EPI_ISL_3667425, EPI_ISL_3667426, EPI_ISL_3667427, EPI_ISL_3667428, EPI_ISL_3667429, EPI_ISL_3667430, EPI_ISL_3667431, EPI_ISL_3667432, EPI_ISL_3667433, EPI_ISL_3667434, EPI_ISL_3667436, EPI_ISL_3667437, EPI_ISL_3667438, EPI_ISL_3667439, EPI_ISL_3667440, EPI_ISL_3667441, EPI_ISL_3667442, EPI_ISL_3667443, EPI_ISL_3667444, EPI_ISL_3667445, EPI_ISL_3667446, EPI_ISL_3667447, EPI_ISL_3667448, EPI_ISL_3667449, EPI_ISL_3667451, EPI_ISL_3667452, EPI_ISL_3667453, EPI_ISL_3667454, EPI_ISL_3667455, EPI_ISL_3667463, EPI_ISL_3667464, EPI_ISL_3667465, EPI_ISL_3667466, EPI_ISL_3667467, EPI_ISL_3667468, EPI_ISL_3667469, EPI_ISL_3667470, EPI_ISL_3667471, EPI_ISL_3667472, EPI_ISL_3667473, EPI_ISL_3667474, EPI_ISL_3667475, EPI_ISL_3667476, EPI_ISL_3667477, EPI_ISL_3667478, EPI_ISL_3667479, EPI_ISL_3667483, EPI_ISL_3667485, EPI_ISL_3667486, EPI_ISL_3667487, EPI_ISL_3667488, EPI_ISL_3667489, EPI_ISL_3667490, EPI_ISL_3667491, EPI_ISL_3667492, EPI_ISL_3667493, EPI_ISL_3667494, EPI_ISL_3667495, EPI_ISL_3667496, EPI_ISL_3667497, EPI_ISL_3667498, EPI_ISL_3667499, EPI_ISL_3667500, EPI_ISL_3667501, EPI_ISL_3667502, EPI_ISL_3667503, EPI_ISL_3667504, EPI_ISL_3667505, EPI_ISL_3667506, EPI_ISL_3667507, EPI_ISL_3667508, EPI_ISL_3667509, EPI_ISL_3667510, EPI_ISL_3667511, EPI_ISL_3667512, EPI_ISL_3667513, EPI_ISL_3667514, EPI_ISL_3667515, EPI_ISL_3667516, EPI_ISL_3667517, EPI_ISL_3667518, EPI_ISL_3667519, EPI_ISL_3667520, EPI_ISL_3667521, EPI_ISL_3667522, EPI_ISL_3667523, EPI_ISL_3667524, EPI_ISL_3667525, EPI_ISL_3667527, EPI_ISL_3667528, EPI_ISL_3667529, EPI_ISL_3667530, EPI_ISL_3667531, EPI_ISL_3667532, EPI_ISL_3667533, EPI_ISL_3667534, EPI_ISL_3667535, EPI_ISL_3667536, EPI_ISL_3667537, EPI_ISL_3667538, EPI_ISL_3667539, EPI_ISL_3667540, EPI_ISL_3667541, EPI_ISL_3667542, EPI_ISL_3667543, EPI_ISL_3667544, EPI_ISL_3667545, EPI_ISL_3667546, EPI_ISL_3667547, EPI_ISL_3667548, EPI_ISL_3667549, EPI_ISL_3667550, EPI_ISL_3667551, EPI_ISL_3667552, EPI_ISL_3667553, EPI_ISL_3667554, EPI_ISL_3667555, EPI_ISL_3667556, EPI_ISL_3667557, EPI_ISL_3667558, EPI_ISL_3667559, EPI_ISL_3667560, EPI_ISL_3667561, EPI_ISL_3667562, EPI_ISL_3667563, EPI_ISL_3667564, EPI_ISL_3667565, EPI_ISL_3667566, EPI_ISL_3667567, EPI_ISL_3667568, EPI_ISL_3667569, EPI_ISL_3667570, EPI_ISL_3667571, EPI_ISL_3667572, EPI_ISL_3667573, EPI_ISL_3667574, EPI_ISL_3667575, EPI_ISL_3667576, EPI_ISL_3667577, EPI_ISL_3667578, EPI_ISL_3667579, EPI_ISL_3667580, EPI_ISL_3667581, EPI_ISL_3667582, EPI_ISL_3667583, EPI_ISL_3667584, EPI_ISL_3667585, EPI_ISL_3667586, EPI_ISL_3667587, EPI_ISL_3667588, EPI_ISL_3667589, EPI_ISL_3667590, EPI_ISL_3667591, EPI_ISL_3667592, EPI_ISL_3667593, EPI_ISL_3667594, EPI_ISL_3667595, EPI_ISL_3667596, EPI_ISL_3667597, EPI_ISL_3667598, EPI_ISL_3667599, EPI_ISL_3667600, EPI_ISL_3667601, EPI_ISL_3667602, EPI_ISL_3667603, EPI_ISL_3667604, EPI_ISL_3667605, EPI_ISL_3667606, EPI_ISL_3667607, EPI_ISL_3667608, EPI_ISL_3667609, EPI_ISL_3667610, EPI_ISL_3667611, EPI_ISL_3667612, EPI_ISL_3667613, EPI_ISL_3667614, EPI_ISL_3667615, EPI_ISL_3667616, EPI_ISL_3667617, EPI_ISL_3667619, EPI_ISL_3667620, EPI_ISL_3667621, EPI_ISL_3667622, EPI_ISL_3667623, EPI_ISL_3667624, EPI_ISL_3667625, EPI_ISL_3667626, EPI_ISL_3667627, EPI_ISL_3667628, EPI_ISL_3667629, EPI_ISL_3667630, EPI_ISL_3667631, EPI_ISL_3667632, EPI_ISL_3667633, EPI_ISL_3667634, EPI_ISL_3667635, EPI_ISL_3667636, EPI_ISL_3667637, EPI_ISL_3667638, EPI_ISL_3667639, EPI_ISL_3667640, EPI_ISL_3667641, EPI_ISL_3667643, EPI_ISL_3667644, EPI_ISL_3834660, EPI_ISL_3834661, EPI_ISL_3834662, EPI_ISL_3834663, EPI_ISL_3834664, EPI_ISL_3834665, EPI_ISL_3834666, EPI_ISL_3834667, EPI_ISL_3834668, EPI_ISL_3834669, EPI_ISL_3834670, EPI_ISL_3834671, EPI_ISL_3834672, EPI_ISL_3834673, EPI_ISL_3834674, EPI_ISL_3834675, EPI_ISL_3834676, EPI_ISL_3834677, EPI_ISL_3834678, EPI_ISL_3834679, EPI_ISL_3834680, EPI_ISL_3834681, EPI_ISL_3834682, EPI_ISL_3834683, EPI_ISL_3834684, EPI_ISL_3834685, EPI_ISL_3834686, EPI_ISL_3834687, EPI_ISL_3834688, EPI_ISL_3834689, EPI_ISL_3834690, EPI_ISL_3834691, EPI_ISL_3834692, EPI_ISL_3834693, EPI_ISL_3834694, EPI_ISL_3834695, EPI_ISL_3834696, EPI_ISL_3834697, EPI_ISL_3834698, EPI_ISL_3834699, EPI_ISL_3834700, EPI_ISL_3834701, EPI_ISL_3834702, EPI_ISL_3834703, EPI_ISL_3834704, EPI_ISL_3834705, EPI_ISL_3834706, EPI_ISL_3834707, EPI_ISL_3834708, EPI_ISL_3834709, EPI_ISL_3834710, EPI_ISL_3834711, EPI_ISL_3834712, EPI_ISL_3834713, EPI_ISL_3834714, EPI_ISL_3834715, EPI_ISL_3834716, EPI_ISL_3834717, EPI_ISL_3834718, EPI_ISL_3834719, EPI_ISL_3834720, EPI_ISL_3834721, EPI_ISL_3834722, EPI_ISL_3834723, EPI_ISL_3834724, EPI_ISL_3834725, EPI_ISL_3834726, EPI_ISL_3834727, EPI_ISL_3834728, EPI_ISL_3834729, EPI_ISL_3834730, EPI_ISL_3834731, EPI_ISL_3834732, EPI_ISL_3834733, EPI_ISL_3834734, EPI_ISL_3834735, EPI_ISL_3834736, EPI_ISL_3834737, EPI_ISL_3834738, EPI_ISL_3834739, EPI_ISL_3834740, EPI_ISL_3834741, EPI_ISL_3834742, EPI_ISL_3834743, EPI_ISL_3834744, EPI_ISL_3834745, EPI_ISL_3834746, EPI_ISL_3834748, EPI_ISL_3834749, EPI_ISL_3834750, EPI_ISL_3834751, EPI_ISL_3834752, EPI_ISL_3834753, EPI_ISL_3834754, EPI_ISL_3834755, EPI_ISL_3834756, EPI_ISL_3834757, EPI_ISL_3834758, EPI_ISL_3834759, EPI_ISL_3834760, EPI_ISL_3834761, EPI_ISL_3834762, EPI_ISL_3834763, EPI_ISL_3834764, EPI_ISL_3834765, EPI_ISL_3834766, EPI_ISL_3834767, EPI_ISL_3834768, EPI_ISL_3834769, EPI_ISL_3834770, EPI_ISL_3834771, EPI_ISL_3834772, EPI_ISL_3834773, EPI_ISL_3834774, EPI_ISL_3834775, EPI_ISL_3834776, EPI_ISL_3834777, EPI_ISL_3834778, EPI_ISL_3834779, EPI_ISL_3834780, EPI_ISL_3834781, EPI_ISL_3834782, EPI_ISL_3834783, EPI_ISL_3834784, EPI_ISL_3834785, EPI_ISL_3834786, EPI_ISL_3834787, EPI_ISL_3834788, EPI_ISL_3834789, EPI_ISL_3834790, EPI_ISL_3834791, EPI_ISL_3834792, EPI_ISL_3834793, EPI_ISL_3834794, EPI_ISL_3834795, EPI_ISL_3834796, EPI_ISL_3834797, EPI_ISL_3834798, EPI_ISL_3834799, EPI_ISL_3834800, EPI_ISL_3834801, EPI_ISL_3834802, EPI_ISL_3834803, EPI_ISL_3834804, EPI_ISL_3834805, EPI_ISL_3834806, EPI_ISL_3834807, EPI_ISL_3834808, EPI_ISL_3834809, EPI_ISL_3834810, EPI_ISL_3834811, EPI_ISL_3834812, EPI_ISL_3834813, EPI_ISL_3834814, EPI_ISL_3834815, EPI_ISL_3834816, EPI_ISL_3834817, EPI_ISL_3834818, EPI_ISL_3834819, EPI_ISL_3834820, EPI_ISL_3834821, EPI_ISL_3834822, EPI_ISL_3834823, EPI_ISL_3834824, EPI_ISL_3834825, EPI_ISL_3834826, EPI_ISL_3834827, EPI_ISL_3834828, EPI_ISL_3834829, EPI_ISL_3834830, EPI_ISL_3834831, EPI_ISL_3834832, EPI_ISL_3834833, EPI_ISL_3834834, EPI_ISL_3834835, EPI_ISL_3834836, EPI_ISL_3834837, EPI_ISL_3834838, EPI_ISL_3834839, EPI_ISL_3834840, EPI_ISL_3834841, EPI_ISL_3834842, EPI_ISL_3834843, EPI_ISL_3834844, EPI_ISL_3834845, EPI_ISL_3834846, EPI_ISL_3834847, EPI_ISL_3834848, EPI_ISL_3834849, EPI_ISL_3834850, EPI_ISL_3834851, EPI_ISL_3834852, EPI_ISL_3834853, EPI_ISL_3834854, EPI_ISL_3834855, EPI_ISL_3834856, EPI_ISL_3834857, EPI_ISL_3834858, EPI_ISL_3834859, EPI_ISL_3834860, EPI_ISL_3834861, EPI_ISL_3834862, EPI_ISL_3834863, EPI_ISL_3834864, EPI_ISL_3834865, EPI_ISL_3834866, EPI_ISL_3834867, EPI_ISL_3834868, EPI_ISL_3834869, EPI_ISL_3834870, EPI_ISL_3834871, EPI_ISL_3834872, EPI_ISL_3834873, EPI_ISL_3834874, EPI_ISL_3834875, EPI_ISL_3834876, EPI_ISL_3834877, EPI_ISL_3834878, EPI_ISL_3834879, EPI_ISL_3834880, EPI_ISL_3834881, EPI_ISL_3834882, EPI_ISL_3834883, EPI_ISL_3834884, EPI_ISL_3834885, EPI_ISL_3834886, EPI_ISL_3834887, EPI_ISL_3834888, EPI_ISL_3834889, EPI_ISL_3834890, EPI_ISL_3834891, EPI_ISL_3834892, EPI_ISL_3834893, EPI_ISL_3834894, EPI_ISL_3834895, EPI_ISL_3834896, EPI_ISL_3834897, EPI_ISL_3834898, EPI_ISL_3834899, EPI_ISL_3834900, EPI_ISL_3834901, EPI_ISL_3834902, EPI_ISL_3834903, EPI_ISL_3834904, EPI_ISL_3834905, EPI_ISL_3834906, EPI_ISL_3834907, EPI_ISL_3834908, EPI_ISL_3834909, EPI_ISL_3834910, EPI_ISL_3834911, EPI_ISL_3834912, EPI_ISL_3834913, EPI_ISL_3834914, EPI_ISL_3834915, EPI_ISL_3834916, EPI_ISL_3834917, EPI_ISL_3834918, EPI_ISL_3834919, EPI_ISL_3834920, EPI_ISL_3834921, EPI_ISL_3834922, EPI_ISL_3834923, EPI_ISL_3834924, EPI_ISL_3834925, EPI_ISL_3834926, EPI_ISL_3834927, EPI_ISL_3834928, EPI_ISL_3834929, EPI_ISL_3834930, EPI_ISL_3834931, EPI_ISL_3834932, EPI_ISL_3834933, EPI_ISL_3834934, EPI_ISL_3834935, EPI_ISL_3834936, EPI_ISL_3834937, EPI_ISL_3834938, EPI_ISL_3834939, EPI_ISL_3834940, EPI_ISL_3834941, EPI_ISL_3834942, EPI_ISL_3834943, EPI_ISL_3834944, EPI_ISL_3834945, EPI_ISL_3834946, EPI_ISL_3834947, EPI_ISL_3834948, EPI_ISL_3834949, EPI_ISL_3834950, EPI_ISL_3834951, EPI_ISL_3834952, EPI_ISL_3834953, EPI_ISL_3834954, EPI_ISL_3834955, EPI_ISL_3834956, EPI_ISL_3834957, EPI_ISL_3834958, EPI_ISL_3834959, EPI_ISL_3834960, EPI_ISL_3834961, EPI_ISL_3834962, EPI_ISL_3834963, EPI_ISL_3834964, EPI_ISL_3834965, EPI_ISL_3834966, EPI_ISL_3834967, EPI_ISL_3834968, EPI_ISL_3834969, EPI_ISL_3834970, EPI_ISL_3834971, EPI_ISL_3834972, EPI_ISL_3834973, EPI_ISL_3834974, EPI_ISL_3834975, EPI_ISL_3834976, EPI_ISL_3834977, EPI_ISL_3834978, EPI_ISL_3834979, EPI_ISL_3834980, EPI_ISL_3834981, EPI_ISL_3834982, EPI_ISL_3834983, EPI_ISL_3834984, EPI_ISL_3834985, EPI_ISL_3834986, EPI_ISL_3834987, EPI_ISL_3834988, EPI_ISL_3834989, EPI_ISL_3834990, EPI_ISL_3834991, EPI_ISL_3834992, EPI_ISL_3834993, EPI_ISL_3834994, EPI_ISL_3834995, EPI_ISL_3834996, EPI_ISL_3834997, EPI_ISL_3834998, EPI_ISL_3834999, EPI_ISL_3835000, EPI_ISL_3835001, EPI_ISL_3835002, EPI_ISL_3835003, EPI_ISL_3835004, EPI_ISL_3835005, EPI_ISL_3835007

| | | | |
|---|---|---|---|
| see above | Shamir Medical Center (Asaf Harofe) | Shamir Medical Center (Asaf Harofe) | Abu Hamad Ramzia; Adina Bar Chaim; Anna Vishnevsky; Chen Weiner; Netta Zuckerman; Nir Rainy; Patricia Benveniste-Lekovitz; Reut Sorek Abramovich; Yevgeni Yegorov |
