## Supplementary material for "δ subvariants of SARS-COV-2 in Israel, Qatar and Bahrain: Optimal vaccination as an effective strategy to block viral evolution and control the pandemic": Acknowledgement table S2 for the GISAID SARS-COV-2 genomes used in this study

We gratefully acknowledge the following Authors from the Originating laboratories responsible for obtaining the specimens, as well as the Submitting laboratories where the genome data were generated and shared via GISAID, on which this research is based.

All Submitters of data may be contacted directly via www.gisaid.org

Authors are sorted alphabetically.

| Accession ID | Originating Laboratory | Submitting Laboratory | Authors |
|---|---|---|---|
| EPI_ISL_1713365, EPI_ISL_1713369, EPI_ISL_1713375, EPI_ISL_1713378, EPI_ISL_1713379, EPI_ISL_1713380, EPI_ISL_1713381, EPI_ISL_1713382, EPI_ISL_1713383, EPI_ISL_1713384, EPI_ISL_1713385, EPI_ISL_1713386, EPI_ISL_1713387, EPI_ISL_1713388, EPI_ISL_1713389, EPI_ISL_1713390, EPI_ISL_1713391, EPI_ISL_1713392, EPI_ISL_1713393, EPI_ISL_1713394, EPI_ISL_1713395, EPI_ISL_1713396, EPI_ISL_1713397, EPI_ISL_1713398, EPI_ISL_1713399, EPI_ISL_1713400, EPI_ISL_1713401, EPI_ISL_1713402, EPI_ISL_1713403, EPI_ISL_1713404, EPI_ISL_1713405, EPI_ISL_1713406, EPI_ISL_1713407, EPI_ISL_1713408, EPI_ISL_1713409, EPI_ISL_1713410, EPI_ISL_1713411, EPI_ISL_1713412, EPI_ISL_1713413, EPI_ISL_1713414, EPI_ISL_1713415, EPI_ISL_1713416, EPI_ISL_1713417, EPI_ISL_1713418, EPI_ISL_1713419, EPI_ISL_1713421, EPI_ISL_1713422, EPI_ISL_1713423, EPI_ISL_1713424, EPI_ISL_1713425, EPI_ISL_1713426, EPI_ISL_1713427, EPI_ISL_1713428, EPI_ISL_1713429, EPI_ISL_1713430, EPI_ISL_1713440, EPI_ISL_1713441, EPI_ISL_1713442, EPI_ISL_1713448, EPI_ISL_1713449, EPI_ISL_1713450, EPI_ISL_1713451, EPI_ISL_1713452, EPI_ISL_1713453, EPI_ISL_1713454, EPI_ISL_1713455, EPI_ISL_1713456, EPI_ISL_1713457, EPI_ISL_1713458, EPI_ISL_1713459, EPI_ISL_1713460, EPI_ISL_1713461, EPI_ISL_1713462, EPI_ISL_1713463, EPI_ISL_1713464, EPI_ISL_1713465, EPI_ISL_1713466, EPI_ISL_1713467, EPI_ISL_1713484, EPI_ISL_1713485, EPI_ISL_1713486, EPI_ISL_1713487, EPI_ISL_1713488, EPI_ISL_1713489, EPI_ISL_1713490, EPI_ISL_1713491, EPI_ISL_1713492, EPI_ISL_1713493, EPI_ISL_1713494, EPI_ISL_1713495, EPI_ISL_1713496, EPI_ISL_1713497, EPI_ISL_1713498, EPI_ISL_1713499, EPI_ISL_1713500, EPI_ISL_1713501, EPI_ISL_1713502, EPI_ISL_1713503, EPI_ISL_1713504, EPI_ISL_1713505, EPI_ISL_1713506, EPI_ISL_1713507, EPI_ISL_1713508, EPI_ISL_1713509, EPI_ISL_1713510, EPI_ISL_1713511, EPI_ISL_1713512, EPI_ISL_1713513, EPI_ISL_1713514, EPI_ISL_1713515, EPI_ISL_1713516, EPI_ISL_1713517, EPI_ISL_1713518, EPI_ISL_1713519, EPI_ISL_1713520, EPI_ISL_1713521, EPI_ISL_1713524, EPI_ISL_1713525, EPI_ISL_1713526, EPI_ISL_1713527, EPI_ISL_1713528, EPI_ISL_1713529, EPI_ISL_1713530, EPI_ISL_1713531, EPI_ISL_1713532, EPI_ISL_1713533, EPI_ISL_1713534, EPI_ISL_1713535, EPI_ISL_1713536, EPI_ISL_1713537, EPI_ISL_1713538, EPI_ISL_1713539, EPI_ISL_1713541, EPI_ISL_1713542, EPI_ISL_1713543, EPI_ISL_1713544, EPI_ISL_1713545, EPI_ISL_1713546, EPI_ISL_1713547, EPI_ISL_1713548, EPI_ISL_1713549, EPI_ISL_1713551, EPI_ISL_1713552, EPI_ISL_1713553, EPI_ISL_1713554, EPI_ISL_1713555, EPI_ISL_1713557, EPI_ISL_1713558, EPI_ISL_1713559, EPI_ISL_1713560, EPI_ISL_1713561, EPI_ISL_1713562, EPI_ISL_1713563, EPI_ISL_1713564, EPI_ISL_1713565, EPI_ISL_1713566, EPI_ISL_1713567, EPI_ISL_1713568, EPI_ISL_1713569, EPI_ISL_1713570, EPI_ISL_1713571, EPI_ISL_1713572, EPI_ISL_1713573, EPI_ISL_1713574, EPI_ISL_1713575, EPI_ISL_1713576, EPI_ISL_1713577, EPI_ISL_1713578, EPI_ISL_1713579, EPI_ISL_1713580, EPI_ISL_1713581, EPI_ISL_1713582, EPI_ISL_1713583, EPI_ISL_1713584, EPI_ISL_1713585, EPI_ISL_1713586, EPI_ISL_1713587, EPI_ISL_1713588, EPI_ISL_1713589, EPI_ISL_1713590, EPI_ISL_1713591, EPI_ISL_1713592, EPI_ISL_1713593, EPI_ISL_1713594, EPI_ISL_1713595, EPI_ISL_1713596, EPI_ISL_1713597, EPI_ISL_1713598, EPI_ISL_1713599, EPI_ISL_1713600, EPI_ISL_1713601, EPI_ISL_1713602, EPI_ISL_1713603, EPI_ISL_1713604, EPI_ISL_1713605, EPI_ISL_1713606, EPI_ISL_1713607, EPI_ISL_1713608, EPI_ISL_1713609, EPI_ISL_1713610, EPI_ISL_1713611, EPI_ISL_1713612, EPI_ISL_1713613, EPI_ISL_1713614, EPI_ISL_1713615, EPI_ISL_1713616, EPI_ISL_1713617, EPI_ISL_1713618, EPI_ISL_1713619, EPI_ISL_1713620, EPI_ISL_1713621, EPI_ISL_1713622, EPI_ISL_1713623, EPI_ISL_1713624, EPI_ISL_1713625, EPI_ISL_1713626, EPI_ISL_1713627, EPI_ISL_1713628, EPI_ISL_1713629, EPI_ISL_1713630, EPI_ISL_1713631, EPI_ISL_1713632, EPI_ISL_1713633, EPI_ISL_1713634, EPI_ISL_1713635, EPI_ISL_1713636, EPI_ISL_1713637, EPI_ISL_1713638, EPI_ISL_1713639, EPI_ISL_1713640, EPI_ISL_1713641, EPI_ISL_1713642, EPI_ISL_1713643, EPI_ISL_1713644, EPI_ISL_1713645, EPI_ISL_1713646, EPI_ISL_1713647, EPI_ISL_1713648, EPI_ISL_1713649, EPI_ISL_1713650, EPI_ISL_1713651, EPI_ISL_1713652, EPI_ISL_1713653, EPI_ISL_1713654, EPI_ISL_1713655, EPI_ISL_1713656, EPI_ISL_1713657, EPI_ISL_1713658, EPI_ISL_1713659, EPI_ISL_1713660, EPI_ISL_1713661, EPI_ISL_1713662, EPI_ISL_1713663, EPI_ISL_1713664, EPI_ISL_1713665, EPI_ISL_1713666, EPI_ISL_1713667, EPI_ISL_1713668, EPI_ISL_1713669, EPI_ISL_1713670, EPI_ISL_1713671, EPI_ISL_1713672, EPI_ISL_1713673, EPI_ISL_1713674, EPI_ISL_1713675, EPI_ISL_1713676, EPI_ISL_1713677, EPI_ISL_1713678, EPI_ISL_1713679, EPI_ISL_1713680, EPI_ISL_1713681, EPI_ISL_1713682, EPI_ISL_1713683, EPI_ISL_1713684, EPI_ISL_1713685, EPI_ISL_1713686, EPI_ISL_1713687, EPI_ISL_1713688, EPI_ISL_1713689, EPI_ISL_1713690, EPI_ISL_1713691, EPI_ISL_1713692, EPI_ISL_1713693, EPI_ISL_1713694, EPI_ISL_1713695, EPI_ISL_1713696, EPI_ISL_1713697, EPI_ISL_1713698, EPI_ISL_1713699, EPI_ISL_1713700, EPI_ISL_1713701, EPI_ISL_1713702, EPI_ISL_1713703, EPI_ISL_1713704, EPI_ISL_1713705, EPI_ISL_1713706, EPI_ISL_1713707, EPI_ISL_1713708, EPI_ISL_1713709, EPI_ISL_1713710, EPI_ISL_1713711, EPI_ISL_1713712, EPI_ISL_1713713, EPI_ISL_1713714, EPI_ISL_1713715, EPI_ISL_1713716, EPI_ISL_1713717, EPI_ISL_1713718, EPI_ISL_1713719, EPI_ISL_1713720, EPI_ISL_1713721, EPI_ISL_1713722, EPI_ISL_1713723, EPI_ISL_1713724, EPI_ISL_1713725, EPI_ISL_1713726, EPI_ISL_1713727, EPI_ISL_1713728, EPI_ISL_1713729, EPI_ISL_1713730, EPI_ISL_1713731, EPI_ISL_1713732, EPI_ISL_1713733, EPI_ISL_1713734, EPI_ISL_1713735, EPI_ISL_1713736, EPI_ISL_1713737, EPI_ISL_1713738, EPI_ISL_1713739, EPI_ISL_1713740, EPI_ISL_1713741, EPI_ISL_1713742, EPI_ISL_1713743, EPI_ISL_1713744, EPI_ISL_1713745, EPI_ISL_1713746, EPI_ISL_1713747, EPI_ISL_1713748, EPI_ISL_1713749, EPI_ISL_1713750, EPI_ISL_1713751, EPI_ISL_1713752, EPI_ISL_1713753, EPI_ISL_1713754, EPI_ISL_1713755, EPI_ISL_1713756, EPI_ISL_1713757, EPI_ISL_1713758, EPI_ISL_1713759, EPI_ISL_1713760, EPI_ISL_1713761, EPI_ISL_1713762, EPI_ISL_1713763, EPI_ISL_1713764, EPI_ISL_1713765, EPI_ISL_1713766, EPI_ISL_1713767, EPI_ISL_1713768, EPI_ISL_1713769, EPI_ISL_1713770, EPI_ISL_1713771, EPI_ISL_1713772, EPI_ISL_1713773, EPI_ISL_1713774, EPI_ISL_1713775, EPI_ISL_1713776, EPI_ISL_1713777, EPI_ISL_1713778, EPI_ISL_1713779, EPI_ISL_1713780, EPI_ISL_1713781, EPI_ISL_1713782, EPI_ISL_1713783, EPI_ISL_1713784, EPI_ISL_1713785, EPI_ISL_1713786, EPI_ISL_1713787, EPI_ISL_1713788, EPI_ISL_1713789, EPI_ISL_1713790, EPI_ISL_1713791, EPI_ISL_1713792, EPI_ISL_1713793, EPI_ISL_1713794, EPI_ISL_1713795, EPI_ISL_1713796, EPI_ISL_1713797, EPI_ISL_1713798, EPI_ISL_1713799, EPI_ISL_1713800, EPI_ISL_1713801, EPI_ISL_1713802, EPI_ISL_1713803, EPI_ISL_1713804, EPI_ISL_1713805, EPI_ISL_1713806, EPI_ISL_1713807, EPI_ISL_1713808, EPI_ISL_1713809, EPI_ISL_1713810, EPI_ISL_1713811, EPI_ISL_1713812, EPI_ISL_1713813, EPI_ISL_1713814, EPI_ISL_1713815, EPI_ISL_1713816, EPI_ISL_1713817, EPI_ISL_1713818, EPI_ISL_1713819, EPI_ISL_1713820, EPI_ISL_1713821, EPI_ISL_1713822, EPI_ISL_1713823, EPI_ISL_1713824, EPI_ISL_1713825, EPI_ISL_1713826, EPI_ISL_1713827, EPI_ISL_1713828, EPI_ISL_1713829, EPI_ISL_1713830, EPI_ISL_1713831, EPI_ISL_1713832, EPI_ISL_1713833, EPI_ISL_1713834, EPI_ISL_1713835, EPI_ISL_1713836, EPI_ISL_1713837, EPI_ISL_1713838, EPI_ISL_1713839, EPI_ISL_1713840, EPI_ISL_1713841, EPI_ISL_1713842, EPI_ISL_1713843, EPI_ISL_1713844, EPI_ISL_1713845, EPI_ISL_1713846, EPI_ISL_1713847, EPI_ISL_1713848, EPI_ISL_1713849, EPI_ISL_1713850, EPI_ISL_1713851, EPI_ISL_1713852, EPI_ISL_1713853, EPI_ISL_1713854, EPI_ISL_1713855, EPI_ISL_1713856, EPI_ISL_1713857, EPI_ISL_1713858, EPI_ISL_1713859, EPI_ISL_1713860, EPI_ISL_1713861, EPI_ISL_1713862, EPI_ISL_1713863, EPI_ISL_1713864, EPI_ISL_1713865, EPI_ISL_1713866, EPI_ISL_1713867, EPI_ISL_1713868, EPI_ISL_1713869, EPI_ISL_1713870, EPI_ISL_1713871, EPI_ISL_1713872, EPI_ISL_1713873, EPI_ISL_1713874, EPI_ISL_1713875, EPI_ISL_1713876, EPI_ISL_1713877, EPI_ISL_1713878, EPI_ISL_1713879, EPI_ISL_1713880, EPI_ISL_1713881, EPI_ISL_1713882, EPI_ISL_1713883, EPI_ISL_1713884, EPI_ISL_1713885, EPI_ISL_1713886, EPI_ISL_1713887, EPI_ISL_1713888, EPI_ISL_1713889, EPI_ISL_1713890, EPI_ISL_1713891, EPI_ISL_1713892, EPI_ISL_1713893, EPI_ISL_1713894, EPI_ISL_1713895, EPI_ISL_1713896, EPI_ISL_1713897, EPI_ISL_1713898, EPI_ISL_1713899, EPI_ISL_1713900, EPI_ISL_1713901, EPI_ISL_1713902, EPI_ISL_1713903, EPI_ISL_1713904, EPI_ISL_1713905, EPI_ISL_1713906, EPI_ISL_1713907, EPI_ISL_1713908, EPI_ISL_1713909, EPI_ISL_1713910, EPI_ISL_1713911, EPI_ISL_1713912, EPI_ISL_1713913, EPI_ISL_1713914, EPI_ISL_1713915, EPI_ISL_1713916, EPI_ISL_1713917, EPI_ISL_1713918, EPI_ISL_1713919, EPI_ISL_1713920, EPI_ISL_1713921, EPI_ISL_1713922, EPI_ISL_1713923, EPI_ISL_1713924, EPI_ISL_1713925, EPI_ISL_1713926, EPI_ISL_1713927, EPI_ISL_1713928, EPI_ISL_1713929, EPI_ISL_1713931, EPI_ISL_1713932, EPI_ISL_1713933, EPI_ISL_1713934, EPI_ISL_1713935, EPI_ISL_1713936, EPI_ISL_1713937, EPI_ISL_1713938, EPI_ISL_1713940, EPI_ISL_1713941, EPI_ISL_1713942, EPI_ISL_1713944, EPI_ISL_1713945, EPI_ISL_1713946, EPI_ISL_2274251, EPI_ISL_2274252, EPI_ISL_2274253, EPI_ISL_2274254, EPI_ISL_2274255, EPI_ISL_2274256, EPI_ISL_2274257, EPI_ISL_2274258, EPI_ISL_2274259, EPI_ISL_2274260, EPI_ISL_2274261, EPI_ISL_2274262, EPI_ISL_2274263, EPI_ISL_2274265, EPI_ISL_2274266, EPI_ISL_2274267, EPI_ISL_2274268, EPI_ISL_2274269, EPI_ISL_2274270, EPI_ISL_2274271, EPI_ISL_2274272, EPI_ISL_2274273, EPI_ISL_2274274, EPI_ISL_2274275, EPI_ISL_2274276, EPI_ISL_2274277, EPI_ISL_2274278, EPI_ISL_2274279, EPI_ISL_2274280, EPI_ISL_2274281, EPI_ISL_2274282, EPI_ISL_2274283, EPI_ISL_2274284, EPI_ISL_2274285, EPI_ISL_2274286, EPI_ISL_2274287, EPI_ISL_2274288, EPI_ISL_2274289, EPI_ISL_2274290, EPI_ISL_2274291, EPI_ISL_2274292, EPI_ISL_2274294, EPI_ISL_2274295, EPI_ISL_2274296, EPI_ISL_2274297, EPI_ISL_2274298, EPI_ISL_2274299, EPI_ISL_2274300, EPI_ISL_2274301, EPI_ISL_2274302, EPI_ISL_2274303, EPI_ISL_2274304, EPI_ISL_2274305, EPI_ISL_2274306, EPI_ISL_2274307, EPI_ISL_2274308, EPI_ISL_2274309, EPI_ISL_2274310, EPI_ISL_2274311, EPI_ISL_2274312, EPI_ISL_2274313, EPI_ISL_2274314, EPI_ISL_2274315, EPI_ISL_2274316, EPI_ISL_2274317, EPI_ISL_2274318, EPI_ISL_2274319, EPI_ISL_2274320, EPI_ISL_2274321, EPI_ISL_2274322, EPI_ISL_2274323, EPI_ISL_2274324, EPI_ISL_2274325, EPI_ISL_2274326, EPI_ISL_2274327, EPI_ISL_2274328, EPI_ISL_2274329, EPI_ISL_2274330, EPI_ISL_2274331, EPI_ISL_2274332, EPI_ISL_2274333, EPI_ISL_2274334, EPI_ISL_2274335, EPI_ISL_2274336, EPI_ISL_2274337, EPI_ISL_2274338, EPI_ISL_2274339, EPI_ISL_2274340, EPI_ISL_2274341, EPI_ISL_2274342, EPI_ISL_2274343, EPI_ISL_2274344, EPI_ISL_2274345, EPI_ISL_2274346, EPI_ISL_2274347, EPI_ISL_2274348, EPI_ISL_2274349, EPI_ISL_2274350, EPI_ISL_2274351, EPI_ISL_2274352, EPI_ISL_2274353, EPI_ISL_2274354, EPI_ISL_2274355, EPI_ISL_2274356, EPI_ISL_2274357, EPI_ISL_2274358, EPI_ISL_2274359, EPI_ISL_2274360, EPI_ISL_2274361, EPI_ISL_2274362, EPI_ISL_2274363, EPI_ISL_2274364, EPI_ISL_2274365, EPI_ISL_2274366, EPI_ISL_2274367, EPI_ISL_2274368, EPI_ISL_2274369, EPI_ISL_2274370, EPI_ISL_2274371, EPI_ISL_2274372, EPI_ISL_2274373, EPI_ISL_2274374, EPI_ISL_2274375, EPI_ISL_2274376, EPI_ISL_2274377, EPI_ISL_2274378, EPI_ISL_2274379, EPI_ISL_2274380, EPI_ISL_2274381, EPI_ISL_2274382, EPI_ISL_2274383, EPI_ISL_2274384, EPI_ISL_2274385, EPI_ISL_2274386, EPI_ISL_2274387, EPI_ISL_2274388, EPI_ISL_2274389, EPI_ISL_2274390, EPI_ISL_2274391, EPI_ISL_2274392, EPI_ISL_2274393, EPI_ISL_2274394, EPI_ISL_2274395, EPI_ISL_2274396, EPI_ISL_2274397, EPI_ISL_2274398, EPI_ISL_2274399, EPI_ISL_2274400, EPI_ISL_2274401, EPI_ISL_2274402, EPI_ISL_2274403, EPI_ISL_2274404, EPI_ISL_2274405, EPI_ISL_2274406, EPI_ISL_2274407, EPI_ISL_2274408, EPI_ISL_2274409, EPI_ISL_2274410, EPI_ISL_2274411, EPI_ISL_2274412, EPI_ISL_2274413, EPI_ISL_2274414, EPI_ISL_2274415, EPI_ISL_2274416, EPI_ISL_2274417, EPI_ISL_2274418, EPI_ISL_2274419, EPI_ISL_2274420, EPI_ISL_2274421, EPI_ISL_2274422, EPI_ISL_2274423, EPI_ISL_2274424, EPI_ISL_2274425, EPI_ISL_2274426, EPI_ISL_2274427, EPI_ISL_2274428, EPI_ISL_2274429, EPI_ISL_2274430, EPI_ISL_2274431, EPI_ISL_2274432, EPI_ISL_2274433, EPI_ISL_2274434, EPI_ISL_2274435, EPI_ISL_2408232, EPI_ISL_2408233, EPI_ISL_2408234, EPI_ISL_2408235, EPI_ISL_2408236, EPI_ISL_2408237, EPI_ISL_2408238, EPI_ISL_2408239, EPI_ISL_2408240, EPI_ISL_2408241, EPI_ISL_2408242, EPI_ISL_2408243, EPI_ISL_2408244, EPI_ISL_2408245, EPI_ISL_2408246, EPI_ISL_2408247, EPI_ISL_2408248, EPI_ISL_2408249, EPI_ISL_2408250, EPI_ISL_2408251, EPI_ISL_2408252, EPI_ISL_2408253, EPI_ISL_2408254, EPI_ISL_2408255, EPI_ISL_2408256, EPI_ISL_2408257, EPI_ISL_2408258, EPI_ISL_2408259, EPI_ISL_2408260, EPI_ISL_2408261, EPI_ISL_2408262, EPI_ISL_2408263, EPI_ISL_2408264, EPI_ISL_2408265, EPI_ISL_2408266, EPI_ISL_2408267, EPI_ISL_2408268, EPI_ISL_2408269, EPI_ISL_2408270, EPI_ISL_2408271, EPI_ISL_2408272, EPI_ISL_2408273, EPI_ISL_2408274, EPI_ISL_2408275, EPI_ISL_2408276, EPI_ISL_2408277, EPI_ISL_2408278, EPI_ISL_2408279, EPI_ISL_2408280, EPI_ISL_2408281, EPI_ISL_2408282, EPI_ISL_2408283, EPI_ISL_2408284, EPI_ISL_2408285, EPI_ISL_2408286, EPI_ISL_2408287, EPI_ISL_2408288, EPI_ISL_2408289, EPI_ISL_2408290, EPI_ISL_2408291, EPI_ISL_2408292, EPI_ISL_2408293, EPI_ISL_2408294, EPI_ISL_2408295, EPI_ISL_2408296, EPI_ISL_2408297, EPI_ISL_2408298, EPI_ISL_2408299, EPI_ISL_2408300, EPI_ISL_2408301, EPI_ISL_2408302, EPI_ISL_2408303, EPI_ISL_2408304, EPI_ISL_2408305, EPI_ISL_2408306, EPI_ISL_2408307, EPI_ISL_2408308, EPI_ISL_2408309, EPI_ISL_2408310, EPI_ISL_2408311, EPI_ISL_2408312, EPI_ISL_2408313, EPI_ISL_2408314, EPI_ISL_2408315, EPI_ISL_2408316, EPI_ISL_2408317, EPI_ISL_2408318, EPI_ISL_2408319, EPI_ISL_2408320, EPI_ISL_2408321, EPI_ISL_2408322, EPI_ISL_2408323, EPI_ISL_2408324, EPI_ISL_2408325, EPI_ISL_2408326, EPI_ISL_2408327, EPI_ISL_2408328, EPI_ISL_2408329, EPI_ISL_2408330, EPI_ISL_2408331, EPI_ISL_2408332, EPI_ISL_2408333, EPI_ISL_2408334, EPI_ISL_2408335, EPI_ISL_2408336, EPI_ISL_2408337, EPI_ISL_2408338, EPI_ISL_2408339, EPI_ISL_2408340, EPI_ISL_2408341, EPI_ISL_2408342, EPI_ISL_2408343, EPI_ISL_2408344, EPI_ISL_2408345, EPI_ISL_2408346, EPI_ISL_2408347, EPI_ISL_2408348, EPI_ISL_2408349, EPI_ISL_2408350, EPI_ISL_2408351, EPI_ISL_2408352, EPI_ISL_2408353, EPI_ISL_2408354, EPI_ISL_2408355, EPI_ISL_2408356, EPI_ISL_2408357, EPI_ISL_2408358, EPI_ISL_2408359, EPI_ISL_2408360, EPI_ISL_2408361, EPI_ISL_2408362, EPI_ISL_2408363, EPI_ISL_2408364, EPI_ISL_2408365, EPI_ISL_2408366, EPI_ISL_2408367, EPI_ISL_2408368, EPI_ISL_2408369, EPI_ISL_2408370, EPI_ISL_2408371, EPI_ISL_2408372, EPI_ISL_2408373, EPI_ISL_2408374, EPI_ISL_2408375, EPI_ISL_2408376, EPI_ISL_2408377, EPI_ISL_2408378, EPI_ISL_2408379, EPI_ISL_2408380, EPI_ISL_2408381, EPI_ISL_2408382, EPI_ISL_2408383, EPI_ISL_2408384, EPI_ISL_2408385, EPI_ISL_2408386, EPI_ISL_2408387, EPI_ISL_2408388, EPI_ISL_2408389, EPI_ISL_2408390, EPI_ISL_2408391, EPI_ISL_2408392, EPI_ISL_2408393, EPI_ISL_2408394, EPI_ISL_2408395, EPI_ISL_2408396, EPI_ISL_2408397, EPI_ISL_2408398, EPI_ISL_2408399, EPI_ISL_2408400, EPI_ISL_2408401, EPI_ISL_2408402, EPI_ISL_2408403, EPI_ISL_2408404, EPI_ISL_2408405, EPI_ISL_2408406, EPI_ISL_2408407, EPI_ISL_2408408, EPI_ISL_2408409, EPI_ISL_2408410, EPI_ISL_2408411, EPI_ISL_2408412, EPI_ISL_2408413, EPI_ISL_2408414, EPI_ISL_2408415, EPI_ISL_2408416, EPI_ISL_2408417, EPI_ISL_2408418, EPI_ISL_2408419, EPI_ISL_2408420, EPI_ISL_2408421, EPI_ISL_2842847, EPI_ISL_2842848, EPI_ISL_2842849, EPI_ISL_2842850, EPI_ISL_2842851, EPI_ISL_2842852, EPI_ISL_2842853, EPI_ISL_2842854, EPI_ISL_2842855, EPI_ISL_2842856, EPI_ISL_2842857, EPI_ISL_2842858, EPI_ISL_2842859, EPI_ISL_2842860, EPI_ISL_2842861, EPI_ISL_2842862, EPI_ISL_2842863, EPI_ISL_2842864, EPI_ISL_2842865, EPI_ISL_2842866, EPI_ISL_2842867, EPI_ISL_2842868, EPI_ISL_2842869, EPI_ISL_2842870, EPI_ISL_2842871, EPI_ISL_2842872, EPI_ISL_2842873, EPI_ISL_2842874, EPI_ISL_2842875, EPI_ISL_2842876, EPI_ISL_2842877, EPI_ISL_2842878, EPI_ISL_2842879, EPI_ISL_2842880, EPI_ISL_2842881, EPI_ISL_2842882, EPI_ISL_2842883, EPI_ISL_2842884, EPI_ISL_2842885, EPI_ISL_2842886, EPI_ISL_2842887, EPI_ISL_2842888, EPI_ISL_2842889, EPI_ISL_2842890, EPI_ISL_2842891, EPI_ISL_2842892, EPI_ISL_2842893, EPI_ISL_2842894, EPI_ISL_2842895, EPI_ISL_2842896, EPI_ISL_2842897, EPI_ISL_2842898, EPI_ISL_2842899, EPI_ISL_2842900, EPI_ISL_2842901, EPI_ISL_2842902, EPI_ISL_2842903, EPI_ISL_2842904, EPI_ISL_2842905, EPI_ISL_2842906, EPI_ISL_2842907, EPI_ISL_2842908, EPI_ISL_2842909, EPI_ISL_2842910, EPI_ISL_2842911, EPI_ISL_2842912, EPI_ISL_2842913, EPI_ISL_2842914, EPI_ISL_2842915, EPI_ISL_2842916, EPI_ISL_2842917, EPI_ISL_2842918, EPI_ISL_2842919, EPI_ISL_2842920, EPI_ISL_2842921, EPI_ISL_2842922, EPI_ISL_2842923, EPI_ISL_2842924, EPI_ISL_2842925, EPI_ISL_2842926, EPI_ISL_2842927, EPI_ISL_2842928, EPI_ISL_2842929, EPI_ISL_2842930, EPI_ISL_2842931, EPI_ISL_2842932, EPI_ISL_2842933, EPI_ISL_2842934, EPI_ISL_2842935, EPI_ISL_2842936, EPI_ISL_2842937, EPI_ISL_2842938, EPI_ISL_2842939, EPI_ISL_2842940, EPI_ISL_2842941, EPI_ISL_2842942, EPI_ISL_2842943, EPI_ISL_2842944, EPI_ISL_2842945, EPI_ISL_2842946, EPI_ISL_2842947, EPI_ISL_2842948, EPI_ISL_2842949, EPI_ISL_2842950, EPI_ISL_2842951, EPI_ISL_2842952, EPI_ISL_2842953, EPI_ISL_2842954, EPI_ISL_2842955, EPI_ISL_2842956, EPI_ISL_2842957, EPI_ISL_2842958, EPI_ISL_2842959, EPI_ISL_2842960, EPI_ISL_2842961, EPI_ISL_2842962, EPI_ISL_2842963, EPI_ISL_2842964, EPI_ISL_2842965, EPI_ISL_2842966, EPI_ISL_2842967, EPI_ISL_2842968, EPI_ISL_2842969, EPI_ISL_2842970, EPI_ISL_2842971, EPI_ISL_2842972, EPI_ISL_2842973, EPI_ISL_2842974, EPI_ISL_2842975, EPI_ISL_2842976, EPI_ISL_2842977, EPI_ISL_2842978, EPI_ISL_2842979, EPI_ISL_2842980, EPI_ISL_2842981, EPI_ISL_2842982, EPI_ISL_2842983, EPI_ISL_2842984, EPI_ISL_2842985, EPI_ISL_2842986, EPI_ISL_2842987, EPI_ISL_2842988, EPI_ISL_2842989, EPI_ISL_2842990, EPI_ISL_2842991, EPI_ISL_2842992, EPI_ISL_2842993, EPI_ISL_2842994, EPI_ISL_2842995, EPI_ISL_2842996, EPI_ISL_2842997, EPI_ISL_2842998, EPI_ISL_2842999, EPI_ISL_2843000, EPI_ISL_2843001, EPI_ISL_2843002, EPI_ISL_2843003, EPI_ISL_2843004, EPI_ISL_2843005, EPI_ISL_2843006, EPI_ISL_2843007, EPI_ISL_2843008, EPI_ISL_2843009, EPI_ISL_2843010, EPI_ISL_2843011, EPI_ISL_2843012, EPI_ISL_2843013, EPI_ISL_2843014, EPI_ISL_2843015, EPI_ISL_2843016, EPI_ISL_2843017, EPI_ISL_2843018, EPI_ISL_2843019, EPI_ISL_2843020, EPI_ISL_2843021, EPI_ISL_2843022, EPI_ISL_2843023, EPI_ISL_2843024, EPI_ISL_2843025, EPI_ISL_2843026, EPI_ISL_2843027, EPI_ISL_2843028, EPI_ISL_2843029, EPI_ISL_2843030, EPI_ISL_2843031, EPI_ISL_2843032, EPI_ISL_2843033, EPI_ISL_2843034, EPI_ISL_2843035, EPI_ISL_2843036, EPI_ISL_2843037, EPI_ISL_2843038, EPI_ISL_2843039, EPI_ISL_2843040, EPI_ISL_2843041, EPI_ISL_2843042, EPI_ISL_2843043, EPI_ISL_2843044, EPI_ISL_2843045, EPI_ISL_2843046, EPI_ISL_2843047, EPI_ISL_2843048, EPI_ISL_2843049, EPI_ISL_2843050, EPI_ISL_2843051, EPI_ISL_2843052, EPI_ISL_2843053, EPI_ISL_2843054, EPI_ISL_2843055, EPI_ISL_2843056, EPI_ISL_2843057, EPI_ISL_2843058, EPI_ISL_2843059, EPI_ISL_2843060, EPI_ISL_2843061, EPI_ISL_2843062, EPI_ISL_2843063, EPI_ISL_2843064, EPI_ISL_2843065, EPI_ISL_2843066, EPI_ISL_2843067, EPI_ISL_2843068, EPI_ISL_2843069, EPI_ISL_2843070, EPI_ISL_2843071, EPI_ISL_2843072, EPI_ISL_2843073, EPI_ISL_2843074, EPI_ISL_2843075, EPI_ISL_2843076, EPI_ISL_2843077, EPI_ISL_2843078, EPI_ISL_2843079, EPI_ISL_2843080, EPI_ISL_2843081, EPI_ISL_2843082, EPI_ISL_2843083, EPI_ISL_2843084, EPI_ISL_2843085, EPI_ISL_2843086, EPI_ISL_2843087, EPI_ISL_2843088, EPI_ISL_2843089, EPI_ISL_2843090, EPI_ISL_2843091, EPI_ISL_2843092, EPI_ISL_2843093, EPI_ISL_2843094, EPI_ISL_2843095, EPI_ISL_2843096, EPI_ISL_2843097, EPI_ISL_2843098, EPI_ISL_2843099, EPI_ISL_2843100, EPI_ISL_2843101, EPI_ISL_2843102, EPI_ISL_2843103, EPI_ISL_2843104, EPI_ISL_2843105, EPI_ISL_2843106, EPI_ISL_2843107, EPI_ISL_2843108, EPI_ISL_2843109, EPI_ISL_2843110, EPI_ISL_2843111, EPI_ISL_2843112, EPI_ISL_2843113, EPI_ISL_2843114, EPI_ISL_2843115, EPI_ISL_2843116, EPI_ISL_2843117, EPI_ISL_2843118, EPI_ISL_2843119, EPI_ISL_2843120, EPI_ISL_2843121, EPI_ISL_2843122, EPI_ISL_2843123, EPI_ISL_2843124, EPI_ISL_2843125, EPI_ISL_2843126, EPI_ISL_2843127, EPI_ISL_2843128, EPI_ISL_2843129, EPI_ISL_2843130, EPI_ISL_2843131, EPI_ISL_2843132, EPI_ISL_2843133, EPI_ISL_2843134, EPI_ISL_2843135, EPI_ISL_2843136, EPI_ISL_2843137, EPI_ISL_2843138, EPI_ISL_2843139, EPI_ISL_2843140, EPI_ISL_2843141, EPI_ISL_2843142, EPI_ISL_2843143, EPI_ISL_2843144, EPI_ISL_2843145, EPI_ISL_2843146, EPI_ISL_2843147, EPI_ISL_2843148, EPI_ISL_2843149, EPI_ISL_2843150, EPI_ISL_2843151, EPI_ISL_2843152, EPI_ISL_2843153, EPI_ISL_2843154, EPI_ISL_2843155, EPI_ISL_2843156, EPI_ISL_2843157, EPI_ISL_2843158, EPI_ISL_2843159, EPI_ISL_2843160, EPI_ISL_2843161, EPI_ISL_2843162, EPI_ISL_2843163, EPI_ISL_2843164, EPI_ISL_2843165, EPI_ISL_2843166, EPI_ISL_2843167, EPI_ISL_2843168, EPI_ISL_2843169, EPI_ISL_2843170, EPI_ISL_2843171, EPI_ISL_2843172, EPI_ISL_2843173, EPI_ISL_2843174, EPI_ISL_2843175, EPI_ISL_2843176, EPI_ISL_2843177, EPI_ISL_2843178, EPI_ISL_2843179, EPI_ISL_2843180, EPI_ISL_2843181, EPI_ISL_2843182, EPI_ISL_2843183, EPI_ISL_2843184, EPI_ISL_2843185, EPI_ISL_2843186, EPI_ISL_2843187, EPI_ISL_2843188, EPI_ISL_2843189, EPI_ISL_2843190, EPI_ISL_2843191, EPI_ISL_2843192, EPI_ISL_2843193, EPI_ISL_2843194, EPI_ISL_2843195, EPI_ISL_2843196, EPI_ISL_2843197, EPI_ISL_2843198, EPI_ISL_2843199, EPI_ISL_2843200, EPI_ISL_2843201, EPI_ISL_2843202, EPI_ISL_2843203, EPI_ISL_2843204, EPI_ISL_2843205, EPI_ISL_2843206, EPI_ISL_2843207, EPI_ISL_2843208, EPI_ISL_2843209, EPI_ISL_2843210, EPI_ISL_2843211, EPI_ISL_2843212, EPI_ISL_2843213, EPI_ISL_2843214, EPI_ISL_2843215, EPI_ISL_2843216, EPI_ISL_2843217, EPI_ISL_2843218, EPI_ISL_2843219, EPI_ISL_2843220, EPI_ISL_2843221, EPI_ISL_2843222, EPI_ISL_2843223, EPI_ISL_2843224, EPI_ISL_2843225, EPI_ISL_2843226, EPI_ISL_2843227, EPI_ISL_2843228, EPI_ISL_2843229, EPI_ISL_2843230, EPI_ISL_2843231, EPI_ISL_2843232, EPI_ISL_2843233, EPI_ISL_2843234, EPI_ISL_2843235, EPI_ISL_2843236, EPI_ISL_2843237, EPI_ISL_2843238, EPI_ISL_2843239, EPI_ISL_2843240, EPI_ISL_2843241, EPI_ISL_2843242, EPI_ISL_2843243, EPI_ISL_2843244, EPI_ISL_2843245, EPI_ISL_2843246, EPI_ISL_2843247, EPI_ISL_2843248, EPI_ISL_2843249, EPI_ISL_2843250, EPI_ISL_2843251, EPI_ISL_2843252, EPI_ISL_2843253, EPI_ISL_2843254, EPI_ISL_2843255, EPI_ISL_2843256, EPI_ISL_2843257, EPI_ISL_2843258, EPI_ISL_2843259, EPI_ISL_2843260, EPI_ISL_2843261, EPI_ISL_2843262, EPI_ISL_2843263, EPI_ISL_2843264, EPI_ISL_2843265, EPI_ISL_2843266, EPI_ISL_2843267, EPI_ISL_2843268, EPI_ISL_2843269, EPI_ISL_2843270, EPI_ISL_2843271, EPI_ISL_2843272, EPI_ISL_2843273, EPI_ISL_2843274, EPI_ISL_2843275, EPI_ISL_2843276, EPI_ISL_2843277, EPI_ISL_2843278, EPI_ISL_2843279, EPI_ISL_2843280, EPI_ISL_2843281, EPI_ISL_2843282, EPI_ISL_2843283, EPI_ISL_2843284, EPI_ISL_2843285, EPI_ISL_2843286, EPI_ISL_2843287, EPI_ISL_2843288, EPI_ISL_2843289, EPI_ISL_2843290, EPI_ISL_2843291, EPI_ISL_2843292, EPI_ISL_2843293, EPI_ISL_2843294, EPI_ISL_2843295, EPI_ISL_2843296, EPI_ISL_2843297, EPI_ISL_2843298, EPI_ISL_2843299, EPI_ISL_2843300, EPI_ISL_2843301, EPI_ISL_2843302, EPI_ISL_2843303, EPI_ISL_2843304 | | | |
| see above | Ministry of Public Health / Hamad Medical Corporation | Biomedical Research Center (BRC), Qatar University / Qatar Genome Project (QGP) | Asmaa A. Al-Thani. MOPH and HMC: Abdullatif Al-Khal; Asmaa A. Al-Thani. MOPH and HMC: Abdullatif Al-Khal; BRC: Fatiha M. Benslimane; Chadi Saad; Dana Al-Batesh; Dina Elgakhlab QGP: Fatima H. Al-Kuwari; Dina Elgakhlab QGP: Fatima H. Al-Kuwari; Einas A. E. Al-Kuwari; Hadi M. Yassine; Hamad E. Al-Romaihi; Hamda Alromaihi; Heba A. Al-Khatib; Mashael A. Al-Bader; Mohammed Al-Thani; Muna A. S. Al-Maslamani; Oal Al-Jamal; Peter V. Coyle; Reham A. El-Kahlout. QBB: Tasneem Al-Hamad; Reham A. El-Kahlout. QBB: Tasneem Al-Hamad; Roberto Bertollini; Salih Al-Marri |
| EPI_ISL_1714275, EPI_ISL_1714276, EPI_ISL_1714277, EPI_ISL_1714278, EPI_ISL_1714279, EPI_ISL_1714280, EPI_ISL_1714281, EPI_ISL_1714282, EPI_ISL_1714283, EPI_ISL_1714284, EPI_ISL_1714285, EPI_ISL_1714286, EPI_ISL_1714287, EPI_ISL_1714288, EPI_ISL_1714289, EPI_ISL_1714290, EPI_ISL_1714291, EPI_ISL_1714292, EPI_ISL_1714293, EPI_ISL_1714294, EPI_ISL_1714295, EPI_ISL_1714296, EPI_ISL_1714297, EPI_ISL_1714298, EPI_ISL_1714299, EPI_ISL_1714300, EPI_ISL_1714301, EPI_ISL_1714302, EPI_ISL_1714303, EPI_ISL_1714304, EPI_ISL_1714305, EPI_ISL_1714306, EPI_ISL_1714307, EPI_ISL_1714308, EPI_ISL_1714309, EPI_ISL_1714310, EPI_ISL_1714311, EPI_ISL_1714312, EPI_ISL_1714313, EPI_ISL_1714314, EPI_ISL_1714315, EPI_ISL_1714316, EPI_ISL_1714317, EPI_ISL_1714318, EPI_ISL_1714319, EPI_ISL_1714320, EPI_ISL_1714321, EPI_ISL_1714322, EPI_ISL_1714323, EPI_ISL_1714324, EPI_ISL_1714325, EPI_ISL_1714326, EPI_ISL_1714327, EPI_ISL_1714328, EPI_ISL_1714329, EPI_ISL_1714330, EPI_ISL_1714331, EPI_ISL_1714332, EPI_ISL_1714333, EPI_ISL_1714334, EPI_ISL_1714335, EPI_ISL_1714336, EPI_ISL_1714337, EPI_ISL_1714338, EPI_ISL_1714339, EPI_ISL_1714340, EPI_ISL_1714341, EPI_ISL_1714342, EPI_ISL_1714343, EPI_ISL_1714344, EPI_ISL_1714345, EPI_ISL_1714346, EPI_ISL_1714347, EPI_ISL_1714348, EPI_ISL_1714349, EPI_ISL_1714350, EPI_ISL_1714351, EPI_ISL_1714352, EPI_ISL_1714353 | | | |
| see above | Ministry of Public Health / Hamad Medical Corporation | Weill Cornell Medical College - Qatar (WCM-Q), Genomics Core Laboratory / Qatar Genome Project (QGP) | Chadi Saad MOPH and HMC: Abdullatif Al-Khal; Dina Elgakhlab; Einas A. E. Al-Kuwari; Hamad E. Al-Romaihi; Hamda Alromaihi; Joel A Malek. QGP: Fatima H. Al-Kuwari; Laith Abu-Raddad; Mashael A. Al-Bader; Meryem Bensaad; Mohammed Al-Thani; Muna A. S. Al-Maslamani; Peter V. Coyle; Reham A. El-Kahlout. QBB: Tasneem Al-Hamad; Roberto Bertollini; Salih Al-Marri; Shameem Younuskunju; WCMQ: Ayeda A. Ahmed; Yasmin Mohamoud |
| EPI_ISL_3417507, EPI_ISL_3417508, EPI_ISL_3417509, EPI_ISL_3417510, EPI_ISL_3417511, EPI_ISL_3417512 | Pathology, Sidra Medicine | Pathology, Sidra Medicine | Hasan; M.R. |
| EPI_ISL_2661491, EPI_ISL_2661492, EPI_ISL_2661493, EPI_ISL_2661494, EPI_ISL_2661495, EPI_ISL_2661496, EPI_ISL_2661497, EPI_ISL_3048164, EPI_ISL_3048165, EPI_ISL_3048166, EPI_ISL_3941764, EPI_ISL_4295235, EPI_ISL_4295237, EPI_ISL_4295239 | | | |
| see above | US Air Force School of Aerospace Medicine | US Air Force School of Aerospace Medicine | Amanda Javorina; Anthony Fries; Carol Garrett; Clarise Starr; Cole Anderson; Deanna Muehleman; Elizabeth Macias; Fritz Castillo; Jennifer Black; Jennifer Meyer; Kelsey Lanter; Sarah Purves; William Gruner |
