## Supplementary material for "δ subvariants of SARS-COV-2 in Israel, Qatar and Bahrain: Optimal vaccination as an effective strategy to block viral evolution and control the pandemic": Acknowledgement table S3 for the GISAID SARS-COV-2 genomes used in this study

We gratefully acknowledge the following Authors from the Originating laboratories responsible for obtaining the specimens, as well as the Submitting laboratories where the genome data were generated and shared via GISAID, on which this research is based.

All Submitters of data may be contacted directly via www.gisaid.org

Authors are sorted alphabetically.

| Accession ID | Originating Laboratory | Submitting Laboratory | Authors |
|---|---|---|---|
| EPI_ISL_4105092, EPI_ISL_4105093, EPI_ISL_4105094, EPI_ISL_4105095, EPI_ISL_4105096, EPI_ISL_4105097, EPI_ISL_4105098, EPI_ISL_4105099, EPI_ISL_4105100, EPI_ISL_4105101, EPI_ISL_4105102, EPI_ISL_4105103, EPI_ISL_4105104, EPI_ISL_4105105, EPI_ISL_4105106, EPI_ISL_4105107, EPI_ISL_4105108, EPI_ISL_4105109, EPI_ISL_4105110, EPI_ISL_4105111, EPI_ISL_4105112, EPI_ISL_4105113, EPI_ISL_4105114, EPI_ISL_4105115, EPI_ISL_4105116, EPI_ISL_4105117, EPI_ISL_4105118, EPI_ISL_4105119, EPI_ISL_4105120, EPI_ISL_4105121, EPI_ISL_4105122, EPI_ISL_4105123, EPI_ISL_4105124, EPI_ISL_4105125, EPI_ISL_4105126, EPI_ISL_4105127, EPI_ISL_4105128, EPI_ISL_4105129, EPI_ISL_4105130, EPI_ISL_4105131, EPI_ISL_4105132, EPI_ISL_4105133, EPI_ISL_4105134, EPI_ISL_4105135, EPI_ISL_4105136, EPI_ISL_4105137, EPI_ISL_4105138, EPI_ISL_4105139, EPI_ISL_4105140, EPI_ISL_4105141, EPI_ISL_4105142, EPI_ISL_4105143, EPI_ISL_4105144, EPI_ISL_4105145, EPI_ISL_4105146, EPI_ISL_4105147, EPI_ISL_4105148, EPI_ISL_4105149, EPI_ISL_4105150, EPI_ISL_4105151, EPI_ISL_4105152, EPI_ISL_4105153, EPI_ISL_4105154, EPI_ISL_4105155, EPI_ISL_4105156, EPI_ISL_4105157, EPI_ISL_4105158, EPI_ISL_4105159, EPI_ISL_4105160, EPI_ISL_4105161, EPI_ISL_4105162, EPI_ISL_4105163, EPI_ISL_4105164, EPI_ISL_4105165, EPI_ISL_4105166, EPI_ISL_4105167, EPI_ISL_4105168, EPI_ISL_4105169, EPI_ISL_4105170, EPI_ISL_4105171, EPI_ISL_4105172, EPI_ISL_4105173, EPI_ISL_4105174, EPI_ISL_4105175, EPI_ISL_4105176, EPI_ISL_4105177, EPI_ISL_4105178, EPI_ISL_4105179, EPI_ISL_4105180, EPI_ISL_4105181, EPI_ISL_4105182, EPI_ISL_4105183, EPI_ISL_4105184, EPI_ISL_4105185, EPI_ISL_4105186, EPI_ISL_4105187, EPI_ISL_4105188, EPI_ISL_4105189, EPI_ISL_4105190, EPI_ISL_4105191, EPI_ISL_4105192, EPI_ISL_4105193, EPI_ISL_4105194, EPI_ISL_4105195, EPI_ISL_4105196, EPI_ISL_4105197, EPI_ISL_4105198, EPI_ISL_4105199, EPI_ISL_4105200, EPI_ISL_4105201, EPI_ISL_4105202, EPI_ISL_4105203, EPI_ISL_4105204, EPI_ISL_4105205, EPI_ISL_4105206, EPI_ISL_4105207, EPI_ISL_4105208, EPI_ISL_4105209, EPI_ISL_4105210, EPI_ISL_4105211, EPI_ISL_4105212, EPI_ISL_4105213, EPI_ISL_4105214, EPI_ISL_4105215, EPI_ISL_4105216, EPI_ISL_4105217, EPI_ISL_4105218, EPI_ISL_4105219, EPI_ISL_4105220, EPI_ISL_4105221, EPI_ISL_4105222, EPI_ISL_4105223, EPI_ISL_4105224, EPI_ISL_4105225, EPI_ISL_4105226, EPI_ISL_4105227, EPI_ISL_4105228, EPI_ISL_4105229, EPI_ISL_4105230, EPI_ISL_4105231, EPI_ISL_4105232, EPI_ISL_4105233, EPI_ISL_4105234, EPI_ISL_4105235, EPI_ISL_4105236, EPI_ISL_4105237, EPI_ISL_4105238, EPI_ISL_4105239, EPI_ISL_4105240, EPI_ISL_4105241, EPI_ISL_4105242, EPI_ISL_4105243, EPI_ISL_4105244, EPI_ISL_4105245, EPI_ISL_4105246, EPI_ISL_4105247, EPI_ISL_4105248, EPI_ISL_4105249, EPI_ISL_4105250, EPI_ISL_4105251, EPI_ISL_4105252, EPI_ISL_4105253, EPI_ISL_4105254, EPI_ISL_4105255, EPI_ISL_4105256, EPI_ISL_4105257, EPI_ISL_4105258, EPI_ISL_4105259, EPI_ISL_4105260, EPI_ISL_4105261, EPI_ISL_4105262, EPI_ISL_4105263, EPI_ISL_4105264, EPI_ISL_4105265, EPI_ISL_4105266, EPI_ISL_4105267, EPI_ISL_4105268, EPI_ISL_4105269, EPI_ISL_4105270, EPI_ISL_4105271, EPI_ISL_4105272, EPI_ISL_4105273, EPI_ISL_4105274, EPI_ISL_4105275, EPI_ISL_4105276, EPI_ISL_4105277, EPI_ISL_4105278, EPI_ISL_4105279, EPI_ISL_4105280, EPI_ISL_4105281, EPI_ISL_4105282, EPI_ISL_4105283, EPI_ISL_4105284, EPI_ISL_4105285, EPI_ISL_4105286, EPI_ISL_4105287, EPI_ISL_4105288, EPI_ISL_4105289, EPI_ISL_4105290, EPI_ISL_4105291, EPI_ISL_4105292, EPI_ISL_4105293, EPI_ISL_4105294, EPI_ISL_4105295, EPI_ISL_4105296, EPI_ISL_4105297, EPI_ISL_4105298, EPI_ISL_4105299, EPI_ISL_4105300, EPI_ISL_4105301, EPI_ISL_4105302, EPI_ISL_4105303, EPI_ISL_4105304, EPI_ISL_4105305, EPI_ISL_4105306, EPI_ISL_4105307, EPI_ISL_4105308, EPI_ISL_4105309, EPI_ISL_4105310, EPI_ISL_4105311, EPI_ISL_4105312, EPI_ISL_4105313, EPI_ISL_4105314, EPI_ISL_4105315, EPI_ISL_4105316, EPI_ISL_4105317, EPI_ISL_4105318, EPI_ISL_5032792, EPI_ISL_5032793, EPI_ISL_5032794, EPI_ISL_5032795, EPI_ISL_5032796, EPI_ISL_5032797, EPI_ISL_5032798, EPI_ISL_5032799, EPI_ISL_5032800, EPI_ISL_5032801, EPI_ISL_5032802, EPI_ISL_5032803, EPI_ISL_5032804, EPI_ISL_5032805, EPI_ISL_5032806, EPI_ISL_5032807, EPI_ISL_5032808, EPI_ISL_5032809, EPI_ISL_5032810, EPI_ISL_5032811, EPI_ISL_5032812, EPI_ISL_5032813, EPI_ISL_5032814, EPI_ISL_5032815, EPI_ISL_5032816, EPI_ISL_5032817, EPI_ISL_5032818, EPI_ISL_5032819, EPI_ISL_5032820, EPI_ISL_5032821, EPI_ISL_5032822, EPI_ISL_5032823, EPI_ISL_5032824, EPI_ISL_5032825, EPI_ISL_5032826, EPI_ISL_5032827, EPI_ISL_5032828, EPI_ISL_5032829, EPI_ISL_5032830, EPI_ISL_5032831, EPI_ISL_5032832, EPI_ISL_5032833, EPI_ISL_5032834, EPI_ISL_5032835, EPI_ISL_5032836, EPI_ISL_5032837, EPI_ISL_5032838, EPI_ISL_5032839, EPI_ISL_5032840, EPI_ISL_5032841, EPI_ISL_5032842, EPI_ISL_5032843, EPI_ISL_5032844, EPI_ISL_5032845, EPI_ISL_5032846, EPI_ISL_5032847, EPI_ISL_5032848, EPI_ISL_5032849, EPI_ISL_5032850, EPI_ISL_5032851, EPI_ISL_5032852, EPI_ISL_5032853, EPI_ISL_5032854, EPI_ISL_5032855, EPI_ISL_5032856, EPI_ISL_5032857, EPI_ISL_5032858, EPI_ISL_5032859, EPI_ISL_5032860, EPI_ISL_5032861, EPI_ISL_5032862, EPI_ISL_5032863, EPI_ISL_5032864, EPI_ISL_5032865, EPI_ISL_5032866, EPI_ISL_5032867, EPI_ISL_5032868, EPI_ISL_5032869, EPI_ISL_5032870, EPI_ISL_5032871, EPI_ISL_5032872, EPI_ISL_5032873, EPI_ISL_5032874, EPI_ISL_5032875, EPI_ISL_5032876, EPI_ISL_5032877, EPI_ISL_5032878, EPI_ISL_5032879, EPI_ISL_5032880, EPI_ISL_5032884, EPI_ISL_5032885, EPI_ISL_5032886, EPI_ISL_5032887, EPI_ISL_5032888 | | | |
| see above | Communicable Disease Laboratory, Public Health Directorate | Communicable Disease Laboratory, Public Health Directorate | AlAbbas, Z.; AlHujairi, Z.; Almoamen, G.; Altaif, Z.; Alwasti, H.; Marhoon, A.; Touq, M. |
